# GLP-1 Receptor Agonists vs Alternatives for Alcohol Use Disorder: A Multi-Target Trial Emulation

**DOI:** 10.1101/2025.06.07.25329184

**Authors:** Patricia J Rodriguez, Jay B Lusk, Hemalkumar B. Mehta, Joseph F. Levy, Andreas Kalogeropoulos, Samir Soneji, Emily Webber, Ty J. Gluckman, Nicholas L Stucky

## Abstract

**Background:** Glucagon-like peptide 1 receptor agonists (GLP-1 RAs) have shown promise for alcohol use disorder (AUD).

**Objective:** To evaluate the association between use of newer GLP-1 RAs (semaglutide, tirzepatide) and alcohol-related hospitalizations among adults with AUD and either type 2 diabetes (T2D) or obesity.

**Methods:** This retrospective target trial emulation study used electronic health record data from Truveta to identify adults with AUD and either T2D or obesity, who initiated a newer GLP-1 RA (semaglutide, tirzepatide) or relevant active comparator between 2018 and 2024. Four target trials were constructed to reflect clinically distinct populations and comparators: (1) ADM trial (patients with T2D and comparators of other anti-diabetic medications [ADM]), (2) AOM trial (patients with obesity but not T2D and comparators of other anti-obesity medications [AOM]), (3) MAUD-T2D trial (patients with T2D and markers of more severe AUD and comparators of medications for alcohol use disorder [MAUD]), (4) MAUD-obesity trial (patients with obesity, no T2D, and markers of more severe AUD and comparators of MAUD). The primary endpoint was time to alcohol-related hospitalization. Non-alcohol-related hospitalization served as a negative control outcome. Propensity score-based methods (weighting and matching) were used to control for confounding. Cox proportional hazards models were used to estimate the treatment effect of newer GLP-1 RA in four target trials.

**Results:** A total of 40,260 patients were identified, including 18,515 in the ADM trial, 9,256 in the AOM trial, 9,975 in the MAUD and T2D trial, and 11,039 in the MAUD and obesity trial. GLP-1 RAs were associated with a lower hazard of alcohol-related hospitalization in the ADM (HR [95% CI]: 0.70 [0.59 - 0.83] vs. sulfonylureas; 0.73 [0.62 - 0.86] vs. other ADMs), AOM (HR: 0.59 [0.48 - 0.74]), MAUD-T2D (HR: 0.36 [0.29 - 0.46]), and MAUD-obesity (HR: 0.32 [0.23 - 0.43]) trials. No significant differences were observed for non-alcohol-related hospitalizations for ADM and MAUD-T2D trials.

**Conclusion:** Newer GLP-1 RAs were associated with reduced risk of alcohol-related hospitalization across clinically distinct populations.

## Introduction

Excessive alcohol use is a leading cause of preventable mortality in the US, with more than 178,000 attributable deaths annually and an estimated economic burden exceeding $200 billion.^1–4^ Alcohol use disorder (AUD) occurs in 10% of US adults,^5^ yet only 2% of adults with AUD receive medication assisted treatment.^6–8^ While efficacious,^9^ current FDA-approved medications for AUD (MAUD) - acamprosate, disulfiram, and naltrexone-have tolerability and adherence challenges that limit their real-world effectiveness.^10–12^

Glucagon-like peptide 1 receptor agonists (GLP-1 RAs) are used primarily for the treatment of type 2 diabetes mellitus (T2D) and obesity, however, a growing body of evidence from both randomized and observational studies suggest that GLP-1 RAs may also reduce alcohol consumption.^13,14^ In one small randomized clinical trial (RCT), a significant reduction in alcohol consumption was found with semaglutide.^15^ Similarly, observational studies of varying quality have shown reductions in negative AUD-related clinical outcomes, including AUD-related hospitalizations and recurrent AUD diagnoses, however populations may lack generalizability and few studies include tirzepatide.^13,14,16,17^

Importantly, gaps exist in understanding the potential impact of GLP-1 RAs on AUD. These include a lack of high-quality observational studies across rigorous and consistently defined populations.^13^ In addition, given its recent approval, few studies have included tirzepatide, which has shown superior effectiveness on T2D-and weight-related endpoints compared to semaglutide.^18–20^ Accordingly, we employed a target trial emulation framework to estimate the effect of newer GLP-1 RAs (semaglutide and tirzepatide) on alcohol-related hospitalizations for adults with AUD, using four target trials involving clinically distinct populations.

## Methods

### Data

This study used a subset of Truveta Data.^21^ Truveta provides access to daily updated and linked electronic health record (EHR) data from a collective of 30 US health care systems. This includes data related to demographics, encounters, diagnoses, vital signs (e.g., weight, body mass index, blood pressure), medication requests (prescriptions), and laboratory tests and results (e.g., hemoglobin A1c, blood alcohol concentration). In addition to EHR data, medication dispensing data (via e-prescribing) includes fills for prescriptions written both within and outside Truveta constituent health care systems, resulting in greater insights into patients’ medication history. Medication dispense histories are updated at the time of the encounter, and include fill dates, NDC or RxNorm codes, quantity dispensed, and days of medication supplied.

Truveta Data are normalized into a common data model through syntactic and semantic normalization.^21^ Truveta Data are then de-identified by expert determination under the Health Insurance Portability and Accountability Act Privacy Rule. Once de-identified, data are available for analysis in R or Python using Truveta Studio. Data for this study were accessed on April 1, 2025. This study used only de-identified patient records and therefore did not require Institutional Review Board approval.

### Study Design

A target trial emulation framework was used to compare on-treatment alcohol-related hospitalizations for adults with AUD who newly initiated a GLP-1 RA (semaglutide or tirzepatide) or an active comparator medication. Newer GLP-1 RAs were compared to alternatives using four separate target trials (Table 1), defined by a disease cohort (T2D vs obesity without T2D) and a treatment-related reason (related to T2D/obesity vs AUD). This yielded four trials: (1) ADM trial, (2) AOM trial, (3) MAUD-T2D trial, (4) MAUD-obesity trial. Each trial included a clinically distinct population with expected differences on both measured and unmeasured variables related to underlying health status, severity of AUD, and primary reason for seeking treatment.

**Table 1:** Target trials and arms. Target trials are defined by the disease cohort (T2D or obesity without T2D) and (presumed) reason for seeking treatment (non-AUD or AUD). Abbreviations: ADM = anti-diabetic medication, AOM = anti-obesity medication, AUD = alcohol use disorder, DPP4 = dipeptidyl peptidase 4 inhibitor, GLP-1 RA = glucagon-like peptide-1 receptor agonist, MAUD = medications for alcohol use disorder (includes acamprosate, disulfiram, and naltrexone), SGLT2i = sodium/glucose cotransporter-2 inhibitor.

|  |  | Cohort |  |
| --- | --- | --- | --- |
|  |  | T2D | Obesity (without T2D) |
| Treatment seeking reason | Non-AUD<br>(T2D or Obesity) | (a) ADM Trial: Patients with T2D seeking a second-line ADM classified as:<br>1. A newer GLP-1 RA (semaglutide, tirzepatide)<br>2. An older GLP-1 RA (liraglutide, dulaglutide, exenatide)<br>3. A sulfonylurea<br>4. Another 2<sup>nd</sup> line ADM (DPP4, SGLT2i) | (b) AOM Trial: Patients with obesity but without T2D seeking an AOM classified as:<br>1. A newer GLP-1 RA (semaglutide, tirzepatide)<br>2. An older GLP-1 RA (liraglutide, dulaglutide, exenatide)<br>3. Another AOM (phentermine and/or topiramate, orlistat) |
|  | AUD | (c) MAUD-T2D Trial: Patients with T2D seeking MAUD treatment classified as:<br>1. A newer GLP-1 RA (semaglutide, tirzepatide)<br>2. Approved MAUD (naltrexone, disulfiram, acamprosate) | (d) MAUD-Obesity Trial: Patients with obesity but without T2D seeking MAUD treatment classified as:<br>1. A newer GLP-1 RA (semaglutide, tirzepatide)<br>2. Approved MAUD (naltrexone, disulfiram, acamprosate) |

This retrospective observational cohort study follows the RECORD-PE reporting guidelines.^22^

### Study population

Each trial included a new-user cohort of adults with AUD and a cohort-specific condition of interest (T2D or obesity, respectively) between 2018 and 2024.

Initiation of a trial-specific medication, defined by pharmacy dispensing, served as the study index event. New use was defined by a prior 2-year negative history of dispensing and administration of trial-specific medications.

T2D was defined by the presence of T2D diagnostic codes within the previous 2 years. For inclusion in the T2D cohorts, a baseline hemoglobin A1c (A1c) was also required (up to 1 year before the index event), though no restrictions were made on the A1c value. Obesity without T2D was defined by a body mass index (BMI) ≥30 kg/m^2^ at baseline, using the most recent BMI in the 12 months prior to the index date. Patients with T2D were excluded from the obesity cohort. To improve observability of historical and follow-up information for this study, the population was restricted to patients with at least two outpatient office visits in the prior 2 years. All code lists are provided in the supplement.

For the MAUD trials, additional restrictions were applied to emulate patients likely to seek treatment for AUD. History of AUD was defined more narrowly, excluding broader and less severe alcohol-related diagnostic codes (ICD10 F10). In addition, a qualifying AUD diagnosis was required within the year prior to the index date, rather than within two years as used in other trials.

Comorbidities, previous medication use, and utilization were assessed using a two-year lookback window. Study design timelines are depicted in supplementary figures S1-S4.

### Outcomes

Patients were followed for up to one year to identify alcohol-related emergency department (ED) visits or hospitalizations, defined as emergency department or inpatient encounters with either alcohol-related diagnosis (including AUD, alcohol withdrawal, and/or other diagnoses related to acute alcohol use; codes in supplement) or testing for blood alcohol, ethyl glucuronide, or ethyl sulfate levels. Among those with AUD, alcohol-related testing in acute care settings was presumed to be suggestive of the potential involvement of alcohol in their visit. Test values themselves were not considered, as values are highly sensitive to time elapsed relative to alcohol consumption.^23^ Unlike claims data, diagnosis positions are not consistently captured in the EHR, particularly for outpatient encounters, including ED visits. As such, some visits captured as outcomes in this study likely represent visits *with* alcohol-related reasons rather than visits specifically *for* alcohol-related reasons.

To test specificity to alcohol-related hospitalizations, non-alcohol-related hospitalizations were also compared as a negative control outcome. Non-alcohol-related hospitalizations were defined as all ED and inpatient encounters other than those meeting the above criteria for an alcohol-related hospitalization. Additional details on negative control outcome selection are provided in the supplement.

Patients were censored at medication discontinuation, initiation of a GLP-1 RA (or a newer GLP-1 RA for those starting on an older GLP-1 RA), the last encounter before April 1, 2025, or 1 year from the index event, whichever occurred first. Medication discontinuation was defined as 60 days without medication on hand, based on fill dates and days’ supply per fill.

## Statistical Analysis

### Treatment selection

Population balancing methods were used to address non-random treatment selection. Different balancing approaches were used for the ADM and AOM trials, compared to the MAUD trials, due to differences in the target population and estimand.

For the ADM and AOM trials, the estimand of interest was the on-treatment average treatment effect (ATE), such that estimates would generalize to the full trial population. The propensity to initiate a newer GLP-1 RA, relative to other trial medications, was estimated using multinomial regression, inclusive of a variety of factors plausibly related to treatment selection, including demographics, clinical factors, comorbidities, and utilization (complete list provided in supplementary methods). We calculated stabilized inverse probability of treatment weights (IPTW), truncated at the 95^th^ percentile to reduce the impact of extreme weights.^24,25^

For MAUD trials, the target population included patients actively seeking treatment for AUD. Therefore, the estimand of interest was the on-treatment, average treatment effect among those treated (ATT) with MAUD, with the expectation that effects generalize to the cohort of patients who would otherwise receive treatment with approved MAUD. Propensity scores were estimated as the likelihood of initiating MAUD, relative to new GLP-1 RAs, using logistic regression. Additional covariates were included to adjust for differences in AUD severity and recency (complete list in supplement). Because weighting approaches yielded poor balance between groups (likely due to the requirement that all patients receive some weight), 1:1 nearest neighbor propensity score matching was applied (with a caliper of 0.05),^26,27^ pairing a patient treated with MAUD to a similar patient treated with a newer GLP-1 RA. Unmatched patients were not included in the analysis.

### Informative censoring

Inverse probability of censoring weights (IPCW) were applied to account for informative censoring, where patients remaining on treatment differed from those who were censored.^28^ The probability of artificial censoring (due to medication discontinuation, switching, or loss to follow-up [last encounter]) before 365 days was first estimated using logistic regression. The model considered demographic, clinical, and utilization factors, as well as the exposure group and the index year.

Stabilized and truncated IPCW were calculated as the inverse probability of artificial censoring.

For ADM and AOM trials, combined weights were calculated as the product of IPCW and IPTW. For MAUD trials, IPCW was applied to the propensity score matched population.

### Outcomes models

The probability of on-treatment hospitalization by 365 days was extracted from weighted Kaplan Meier curves. Unweighted curves were also plotted for comparison (supplement, Figure S6). Cox proportional hazards models with robust standard errors were used to estimate the on-treatment hazard of alcohol-related hospitalization between treatment groups. The same approach was used to estimate the hazard of non-alcohol-related hospitalizations between groups. E-values were calculated to estimate the magnitude of unmeasured confounding required to negate the observed treatment effects.^29^

## Results

In total, 40,260 patients met the criteria for at least one trial. This included 18,515 in the ADM trial (newer GLP-1 RA: 4,034 [22%]; older GLP-1 RA: 2,190 [12%]; sulfonylurea: 5,217 [28%]; other ADM: 7074 [38%]), 9,256 in the AOM trial (newer GLP-1 RA: 3,511 [38%]; older GLP-1 RA: 609 [7%]; other AOM: 5136 [55%]), 8,875 in the MAUD-T2D trial (newer GLP-1 RA: 5,574 [63%]; MAUD: 3301 [37%]) and 11,039 in the MAUD-Obesity trial (newer GLP-1 RA: 2,003 [18%]; MAUD: 9,036 [82%]) (Table 2).

**Table 2:** Characteristics of patients at baseline before balancing. Quantitative variables are expressed as mean (standard deviation). Categorical variables are expressed as number (percentage). Utilization counts are expressed as median (IQR). Durations refers to the time (years) since first evidence. Other race includes Asian, American Indian or Alaska Native, Native Hawaiian or Other Pacific Islander, other race, unknown, or declined to answer. Abbreviations: AUD = alcohol use disorder, AST = aspartate aminotransferase, ALT = alanine aminotransferase, A1c = glycated hemoglobin, BMI = body mass index, DPP4 = dipeptidyl peptidase 4 inhibitor, MAUD = medications for alcohol use disorder (includes acamprosate, disulfiram, and naltrexone), SD = standard deviation, SGLT2i = sodium/glucose cotransporter-2 inhibitor, SSRI = selective serotonin reuptake inhibitor, T2D = type 2 diabetes.

|  | ADM trial<br>N = 18,515 |  |  |  | AOM trial<br>N = 9,256 |  |  | MAUD-T2D trial<br>N = 8,875 |  | MAUD-Obesity trial<br>N = 11,039 |  |
| --- | --- | --- | --- | --- | --- | --- | --- | --- | --- | --- | --- |
| Characteristic | Newer<br>GLP-1<br>RA<br>N =<br>4,034 | Older<br>GLP-1<br>RA<br>N =<br>2,190 | Sulfonyl<br>urea<br>N =<br>5,217 | Other<br>ADM<br>N =<br>7,074 | Newer<br>GLP-1<br>RA<br>N =<br>3,511 | Older<br>GLP-1<br>RA<br>N =<br>609 | Other<br>AOM<br>N =<br>5,136 | Newer<br>GLP-1<br>RA<br>N =<br>5,574 | MAUD<br>N =<br>3,301 | Newer<br>GLP-1<br>RA<br>N =<br>2,003 | MAUD<br>N =<br>9,036 |
| Age | 57.9<br>(11.5) | 56.8<br>(11.6) | 60.3<br>(12.0) | 61.2<br>(11.9) | 53.0<br>(12.7) | 49.6<br>(12.5) | 47.5<br>(13.1) | 59.5<br>(11.2) | 55.1<br>(11.8) | 52.9<br>(12.6) | 48.1<br>(12.9) |
| Female | 1,476<br>(37%) | 798<br>(36%) | 1,179<br>(23%) | 1,579<br>(22%) | 1,863<br>(53%) | 380<br>(62%) | 3,406<br>(66%) | 1,687<br>(30%) | 966<br>(29%) | 1,090<br>(54%) | 3,874<br>(43%) |
| Race |  |  |  |  |  |  |  |  |  |  |  |
| Black | 393<br>(9.7%) | 303<br>(14%) | 510<br>(9.8%) | 940<br>(13%) | 265<br>(7.5%) | 71<br>(12%) | 446<br>(8.7%) | 584<br>(10%) | 389<br>(12%) | 172<br>(8.6%) | 542<br>(6.0%) |
| White | 3,082<br>(76%) | 1,491<br>(68%) | 3,518<br>(67%) | 4,702<br>(66%) | 2,785<br>(79%) | 456<br>(75%) | 3,854<br>(75%) | 4,101<br>(74%) | 2,399<br>(73%) | 1,535<br>(77%) | 7,346<br>(81%) |
| Other or unknown | 559<br>(14%) | 396<br>(18%) | 1,189<br>(23%) | 1,432<br>(20%) | 461<br>(13%) | 82<br>(13%) | 836<br>(16%) | 889<br>(16%) | 513<br>(16%) | 296<br>(15%) | 1,148<br>(13%) |
| Ethnicity |  |  |  |  |  |  |  |  |  |  |  |
| Hispanic or Latino | 465<br>(12%) | 237<br>(11%) | 632<br>(12%) | 757<br>(11%) | 269<br>(7.7%) | 55<br>(9.0%) | 511<br>(9.9%) | 629<br>(11%) | 347<br>(11%) | 154<br>(7.7%) | 706<br>(7.8%) |
| Not Hispanic or Latino | 3,439<br>(85%) | 1,856<br>(85%) | 4,223<br>(81%) | 5,870<br>(83%) | 3,102<br>(88%) | 530<br>(87%) | 4,453<br>(87%) | 4,721<br>(85%) | 2,759<br>(84%) | 1,762<br>(88%) | 7,923<br>(88%) |
| Unknown | 130<br>(3.2%) | 97<br>(4.4%) | 362<br>(6.9%) | 447<br>(6.3%) | 140<br>(4.0%) | 24<br>(3.9%) | 172<br>(3.3%) | 224<br>(4.0%) | 195<br>(5.9%) | 87<br>(4.3%) | 407<br>(4.5%) |
| Any college on record | 1,592<br>(39%) | 736<br>(34%) | 1,221<br>(23%) | 1,758<br>(25%) | 1,912<br>(54%) | 326<br>(54%) | 2,956<br>(58%) | 1,883<br>(34%) | 1,201<br>(36%) | 1,056<br>(53%) | 4,842<br>(54%) |
| Baseline health |  |  |  |  |  |  |  |  |  |  |  |
| Baseline A1c | 7.4<br>(1.8) | 8.2<br>(2.1) | 8.4<br>(2.1) | 7.9<br>(2.0) | - | - | - | 7.8 (1.9) | 7.1 (1.9) |  |  |
| Baseline BMI | 36.8<br>(7.4) | 35.7<br>(8.1) | 31.6<br>(7.1) | 31.8<br>(7.1) | 38.3<br>(6.7) | 39.7<br>(7.3) | 37.1<br>(6.2) | 35.8<br>(7.5) | 31.2<br>(7.2) | 38.6<br>(6.9) | 35.4<br>(5.2) |
| Unknown | 751 | 478 | 1,060 | 1,510 | - | - | - | 908 | 485 | - | - |
| Baseline ALT | 36.0<br>(26.0) | 35.5<br>(24.7) | 36.6<br>(27.3) | 33.2<br>(24.1) | 35.3<br>(25.4) | 33.2<br>(22.5) | 33.4<br>(25.1) | 35.4<br>(26.6) | 44.6<br>(34.5) | 35.0<br>(25.7) | 47.8<br>(36.3) |
| Unknown | 329 | 213 | 468 | 597 | 701 | 153 | 1,174 | 452 | 170 | 352 | 1,547 |
| Baseline AST | 31.0<br>(24.0) | 30.1<br>(22.5) | 32.7<br>(26.0) | 30.7<br>(23.6) | 29.8<br>(22.3) | 29.2<br>(23.4) | 29.8<br>(24.6) | 29.8<br>(23.5) | 50.2<br>(45.2) | 29.9<br>(24.3) | 49.2<br>(45.0) |
| Unknown | 306 | 218 | 478 | 586 | 692 | 150 | 1,164 | 449 | 156 | 350 | 1,516 |
| Comorbidities |  |  |  |  |  |  |  |  |  |  |  |
| Elixhauser comorbidity score | 9.5<br>(10.4) | 10.6<br>(10.9) | 10.9<br>(11.3) | 13.3<br>(11.8) | 5.2<br>(10.1) | 4.8<br>(10.9) | 3.4 (9.9) | 10.8<br>(11.1) | 10.2<br>(11.1) | 5.0<br>(10.3) | 2.5<br>(10.3) |
| Cirrhosis | 383<br>(9.5%) | 240<br>(11%) | 660<br>(13%) | 907<br>(13%) | 192<br>(5.5%) | 46<br>(7.6%) | 237<br>(4.6%) | 617<br>(11%) | 520<br>(16%) | 124<br>(6.2%) | 712<br>(7.9%) |
| Chronic kidney disease | 1,002<br>(25%) | 711<br>(32%) | 1,617<br>(31%) | 2,805<br>(40%) | 388<br>(11%) | 78<br>(13%) | 512<br>(10.0%) | 1,778<br>(32%) | 858<br>(26%) | 238<br>(12%) | 804<br>(8.9%) |
| Chronic liver disease | 556<br>(14%) | 324<br>(15%) | 926<br>(18%) | 1,233<br>(17%) | 294<br>(8.4%) | 69<br>(11%) | 412<br>(8.0%) | 864<br>(16%) | 771<br>(23%) | 180<br>(9.0%) | 1,145<br>(13%) |
| Cardiovascular disease | 1,277<br>(32%) | 756<br>(35%) | 1,746<br>(33%) | 2,955<br>(42%) | 373<br>(11%) | 76<br>(12%) | 418<br>(8.1%) | 2,064<br>(37%) | 814<br>(25%) | 206<br>(10%) | 660<br>(7.3%) |
| Obesity | 3,643<br>(90%) | 1,910<br>(87%) | 4,063<br>(78%) | 5,703<br>(81%) | 3,511<br>(100%) | 609<br>(100%) | 5,136<br>(100%) | 4,961<br>(89%) | 2,568<br>(78%) | 2,003<br>(100%) | 9,036<br>(100%) |
| Obstructive sleep apnea | 1,823<br>(45%) | 866<br>(40%) | 1,326<br>(25%) | 2,313<br>(33%) | 1,339<br>(38%) | 203<br>(33%) | 1,363<br>(27%) | 2,506<br>(45%) | 946<br>(29%) | 763<br>(38%) | 1,988<br>(22%) |
| Hypertension | 3,489<br>(86%) | 1,902<br>(87%) | 4,554<br>(87%) | 6,350<br>(90%) | 2,361<br>(67%) | 382<br>(63%) | 2,727<br>(53%) | 4,958<br>(89%) | 2,794<br>(85%) | 1,312<br>(66%) | 5,533<br>(61%) |
| Hyperlipidemia | 3,361<br>(83%) | 1,811<br>(83%) | 4,179<br>(80%) | 5,936<br>(84%) | 2,059<br>(59%) | 263<br>(43%) | 2,107<br>(41%) | 4,829<br>(87%) | 2,462<br>(75%) | 1,134<br>(57%) | 3,681<br>(41%) |
| Major depressive disorder | 1,340<br>(33%) | 887<br>(41%) | 1,444<br>(28%) | 1,994<br>(28%) | 1,141<br>(32%) | 224<br>(37%) | 2,337<br>(46%) | 1,836<br>(33%) | 1,429<br>(43%) | 686<br>(34%) | 3,883<br>(43%) |
| Opioid use disorder | 242<br>(6.0%) | 182<br>(8.3%) | 331<br>(6.3%) | 393<br>(5.6%) | 196<br>(5.6%) | 54<br>(8.9%) | 457<br>(8.9%) | 381<br>(6.8%) | 266<br>(8.1%) | 121<br>(6.0%) | 627<br>(6.9%) |
| Other substance use disorder | 343<br>(8.5%) | 318<br>(15%) | 528<br>(10%) | 808<br>(11%) | 306<br>(8.7%) | 96<br>(16%) | 841<br>(16%) | 548<br>(9.8%) | 518<br>(16%) | 206<br>(10%) | 1,348<br>(15%) |
| Previous or concurrent medications |  |  |  |  |  |  |  |  |  |  |  |
| History of metformin | 2,690<br>(67%) | 1,525<br>(70%) | 3,843<br>(74%) | 4,701<br>(66%) | 321<br>(9.1%) | 102<br>(17%) | 229<br>(4.5%) | 3,854<br>(69%) | 1,867<br>(57%) | 212<br>(11%) | 203<br>(2.2%) |
| History of MAUD | 349<br>(8.7%) | 157<br>(7.2%) | 230<br>(4.4%) | 347<br>(4.9%) | 570<br>(16%) | 106<br>(17%) | 933<br>(18%) | 0 (0%) | 0 (0%) | 0 (0%) | 0 (0%) |
| History of opioid | 1,736<br>(43%) | 1,091<br>(50%) | 2,432<br>(47%) | 3,601<br>(51%) | 1,271<br>(36%) | 268<br>(44%) | 2,309<br>(45%) | 2,589<br>(46%) | 1,709<br>(52%) | 763<br>(38%) | 3,444<br>(38%) |
| History of opioid antagonist | 694<br>(17%) | 354<br>(16%) | 777<br>(15%) | 1,171<br>(17%) | 699<br>(20%) | 143<br>(23%) | 1,127<br>(22%) | 810<br>(15%) | 1,989<br>(60%) | 266<br>(13%) | 5,263<br>(58%) |
| History of SSRI | 1,394<br>(35%) | 886<br>(40%) | 1,520<br>(29%) | 2,066<br>(29%) | 1,524<br>(43%) | 296<br>(49%) | 2,547<br>(50%) | 1,853<br>(33%) | 1,596<br>(48%) | 871<br>(43%) | 4,763<br>(53%) |
| AUD severity |  |  |  |  |  |  |  |  |  |  |  |
| AUD duration (years) | 3.1<br>(3.0) | 3.0<br>(3.1) | 2.7<br>(2.7) | 3.0<br>(3.0) | 2.9 (2.8) | 3.1<br>(3.1) | 2.6 (2.6) | 3.6 (3.4) | 3.0 (3.0) | 3.4 (3.3) | 2.3 (2.7) |
| Dates with AUD diagnosis, previous 2 years (n) | 3 (1-9) | 4 (2-11) | 3 (1-9) | 4 (1-10) | 3 (1-8) | 3 (1-9) | 3 (2-9) | 4 (2-11) | 6 (3-13) | 4 (2-9) | 4 (2-8) |
| Most recent AUD diagnosis (years before index) | 0.4<br>(0.5) | 0.4<br>(0.5) | 0.4<br>(0.5) | 0.4<br>(0.5) | 0.5 (0.5) | 0.5<br>(0.5) | 0.4 (0.5) | 0.2 (0.3) | 0.1 (0.2) | 0.3 (0.3) | 0.1 (0.2) |
| Met narrow AUD definition (other than F10) | 3,341<br>(83%) | 1,939<br>(89%) | 4,662<br>(89%) | 6,210<br>(88%) | 2,705<br>(77%) | 544<br>(89%) | 4,424<br>(86%) | 5,574<br>(100%) | 3,301<br>(100%) | 2,003<br>(100%) | 9,036<br>(100%) |
| Had any alcohol-related hospitalization in prior 2 years | 940<br>(23%) | 684<br>(31%) | 1,675<br>(32%) | 2,540<br>(36%) | 740<br>(21%) | 191<br>(31%) | 1,620<br>(32%) | 1,441<br>(26%) | 1,754<br>(53%) | 326<br>(16%) | 3,336<br>(37%) |
| Years in Truveta (since first encounter) | 12.6<br>(11.9) | 12.0<br>(11.7) | 11.2<br>(10.8) | 11.4<br>(11.0) | 12.8<br>(11.8) | 14.0<br>(12.1) | 12.7<br>(11.7) | 13.1<br>(12.4) | 13.1<br>(12.2) | 12.8<br>(12.0) | 13.7<br>(12.7) |
| Outpatient visits in previous 0-12 months (n) | 9 (5-18) | 11 (5-23) | 8 (4-17) | 10 (5-21) | 7 (4-14) | 9 (4-19) | 9 (4-18) | 10 (5-21) | 9 (4-18) | 7 (4-15) | 7 (3-14) |
| Outpatient visits in previous 13-24 months (n) | 7 (3-16) | 9 (3-21) | 6 (2-14) | 7 (2-16) | 5 (2-12) | 6 (2-14) | 6 (2-15) | 7 (3-17) | 6 (2-14) | 5 (1-11) | 4 (1-10) |

Patient characteristics differed across trials (Table 2). Patients with T2D were older and less likely to be female (ADM trial: mean age 59.7; 27% female, MAUD-T2D trial age 57.9; 30% female), compared to those in the obesity cohorts (AOM trial: age 49.7; 61% female, MAUD-obesity trial age 49.0; 45% female). Patients in the MAUD trials had markers of more recent and more severe AUD, as indicated by the number of visits with AUD diagnoses and the proportion of patients with previous alcohol-related hospitalizations. Across all trials, the distribution of initiation time differed across exposure groups, with users of the newer GLP-1 RAs having started their treatment more recently (supplemental Figure S5).

### ADM trial

Before balancing, patients initiating a newer GLP-1 RA were more likely to be White and female, with markers of better health overall (as indicated by a lower Elixhauser comorbidity score,^30^ lower prevalence of most comorbidities, and a lower baseline A1c). IPTW achieved good balance, with all standardized mean differences < 0.1 (supplement, Table S1).

After weighting, alcohol-related hospitalization within one year of treatment occurred in 10.1% (8.6% - 11.5%) of patients on a newer GLP-1 RA, 9.9% (8.2% - 11.6%) of patients on an older GLP-1 RA,14.4% (13.7% - 14.9%) of patients on a sulfonylurea, and 13.9% (12.9% - 15.0%) of patients on other ADMs (DPP4i or SGLT2i) (Figure 1A). Newer GLP-1 RAs were associated with a lower hazard of alcohol-related hospitalization compared to sulfonylureas (HR: 0.70 [0.59 - 0.83]; e-value: 2.2) and other ADMs (HR: 0.73 [0.62 - 0.86]; e-value: 2.1) (Figure 2A), but not when compared to older GLP-1 RAs (HR: 1.04 [0.84 - 1.30]).

**Figure 1:**
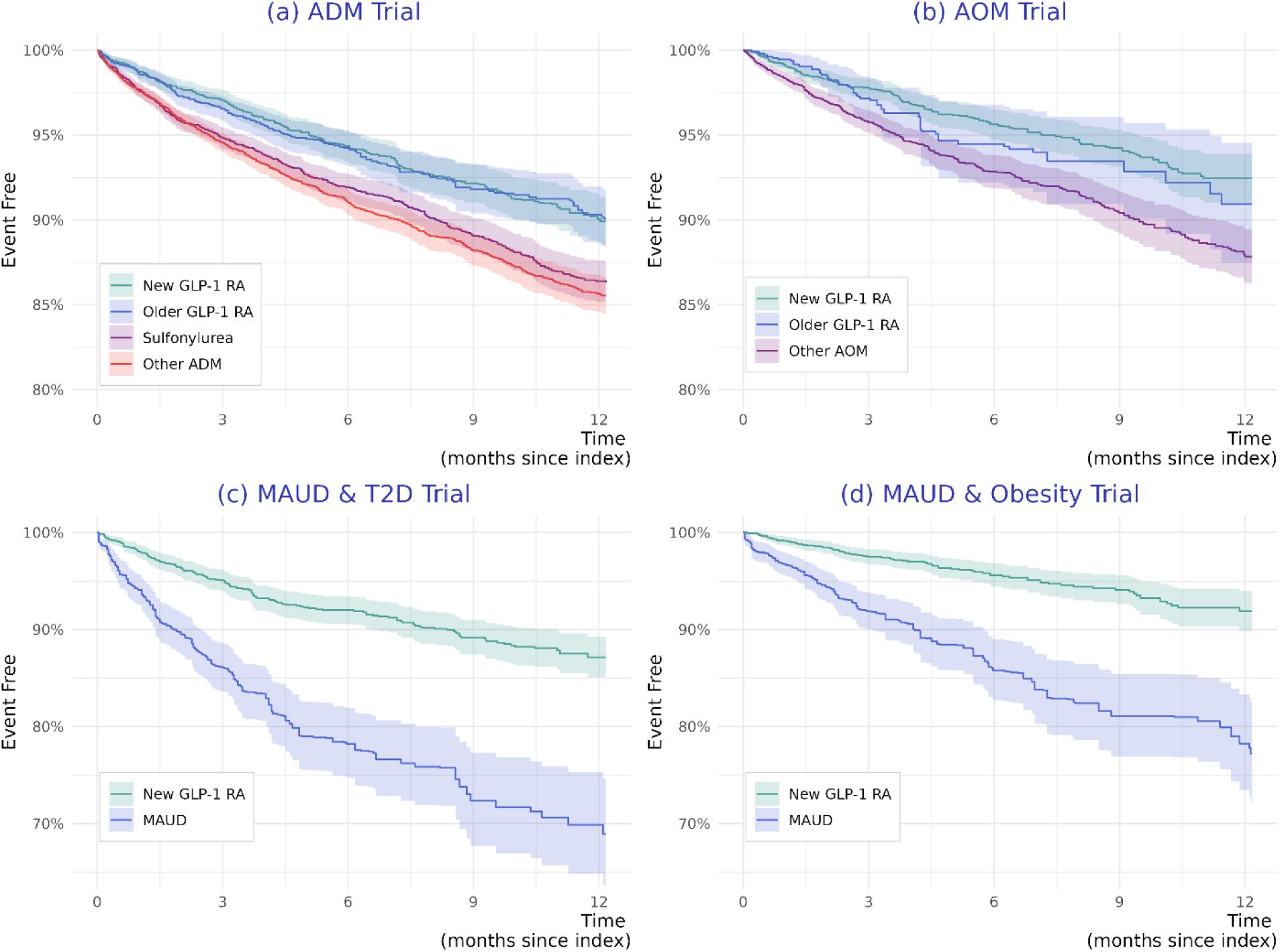
Time to alcohol-related hospitalization. Panel A: ADM trial, Panel B: AOM trial, Panel C: MAUD (T2D) trial, Panel D MAUD (Obesity) trial. Y-axis represents the probability of being event free (1-survival), x-axis represents time since initiation. Patients were censored at discontinuation, switching to a GLP-1 RA, or last encounter. Panels (a) and (b) use IPCW*ICTW weights. Panels (c) and (d) use IPCW weights on a matched population.

**Figure 2:**
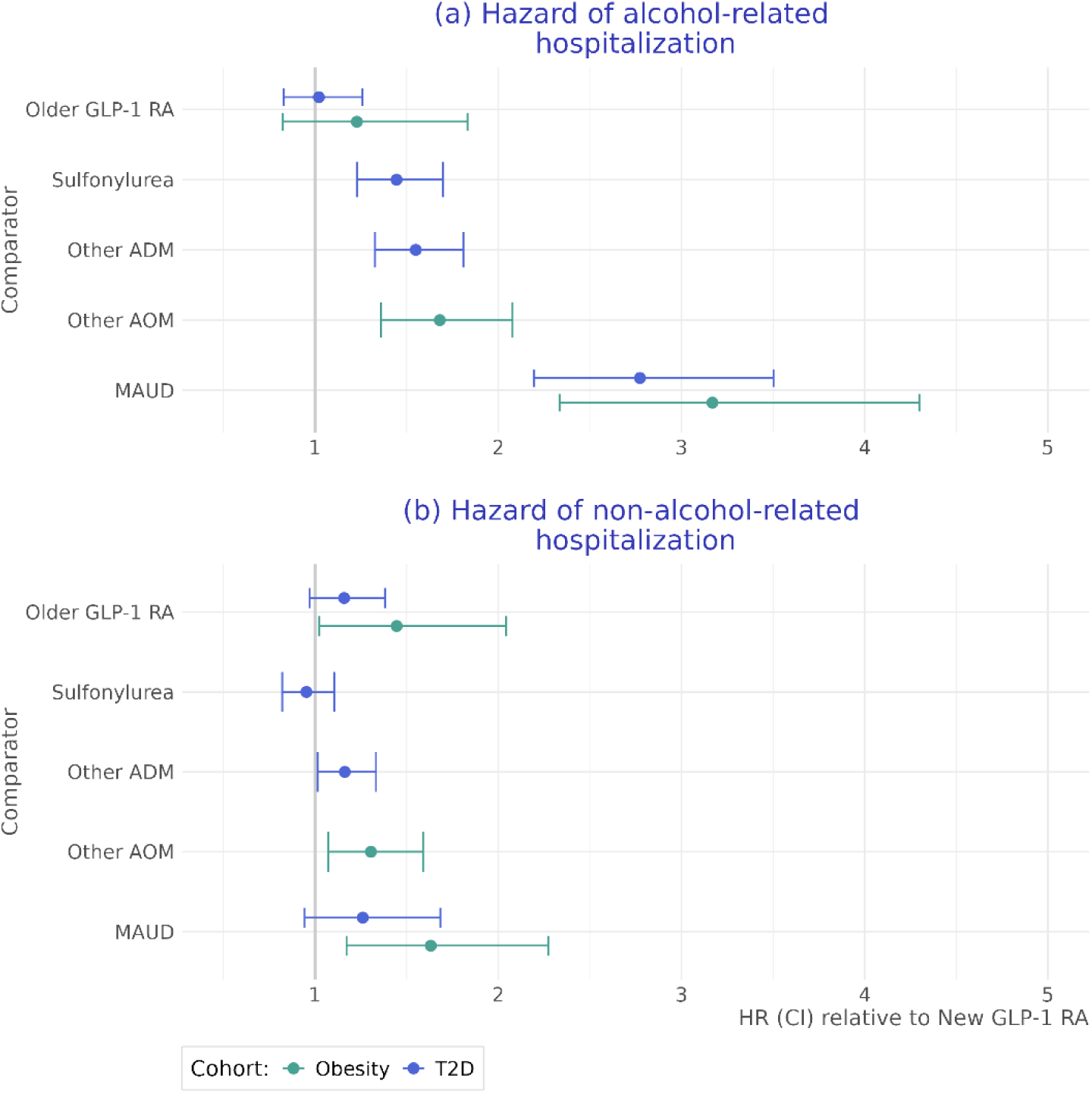
Hazard of alcohol-related and non-alcohol-related hospitalizations relative to initiation of a new GLP-1 RA (semaglutide and tirzepatide). Panel A: Alcohol related hospitalizations. Panel B: Non-alcohol-related hospitalizations.

### AOM trial

Before balancing, patients initiating a newer GLP-1 RA were younger, less likely to be female, and had a higher Elixhauser comorbidity score. IPTW achieved good balance, with all standardized mean differences < 0.1 (supplement, Table S1).

After weighting, alcohol-related hospitalization within one year of treatment occurred in 7.5% (6.1% - 9.0%) of patients on a newer GLP-1 RA, compared to 9.1% (5.5% - 12.5%) of patients on an older GLP-1 RA, and 12.1% (10.6% - 13.7%) of patients on other AOMs (Figure 1B). Newer GLP-1 RAs were associated with a lower hazard of alcohol-related hospitalization compared to other AOMs (HR: 0.59 [0.48 - 0.74]; e-value: 2.8), but not when compared to older GLP-1 RAs (HR: 0.81 [0.55 - 1.21]) (Figure 2A).

### MAUD-T2D trial

Before matching, patients with T2D initiating approved MAUD (n = 3,301) differed from those initiating a newer GLP-1 RA (n = 5,574) in several ways. While demographics were largely similar, patients initiating MAUD had a lower BMI and A1c and higher levels of hepatic biomarkers (aspartate aminotransferase (AST) and alanine aminotransferase (ALT)) associated with heavy alcohol use.^23,31^ They also had several markers of more severe AUD, with more visits in the previous 2 years with AUD, a greater number of recent AUD diagnoses, and more alcohol-related hospitalizations in the previous year. MAUD initiators were also more likely to have evidence of conditions associated with prescribing opioids and opioid antagonists. Propensity score matching yielded a balanced sample of 3,178 patients that approximated the MAUD population, with all standardized mean differences < 0 (supplement, Table S1). Unmatched patients included those on both newer GLP-1 RA (n= 3,985) and MAUD (n= 1,712).

Among matched patients, 1,290 (81%) of those initiating MAUD were censored due to discontinuation before 365 days, compared to 773 (49%) patients initiating a new GLP-1 RA.

After matching and IPCW weighting, alcohol-related hospitalization occurred in 12.9% (10.7% - 14.9%) of patients on a newer GLP-1 RA and 31.1% (25.4% - 36.3%) of patients on MAUD (Figure 1C), yielding a hazard ratio of 0.36 (0.29 - 0.46; e-value: 5.0) (Figure 2A).

### MAUD-obesity trial

Before matching, patients with obesity initiated on approved MAUD (n= 9,036) were younger, less likely to be female, and more likely to be White compared to patients initiated on a newer GLP-1 RA (n = 2,003). While those initiated on MAUD had a lower Elixhauser comorbidity index, they had a higher prevalence of substance use disorder and greater prior prescribing of opioids and opioid antagonists. Similarly, they had a higher rate of AUD-related hospitalization in the prior year and higher AST and ALT levels at baseline. Propensity score matching yielded a balanced sample of 3,276 patients that approximated the MAUD population, with all standardized mean differences < 0 (supplement, Table S1). Unmatched patients included those on both newer GLP-1 RA (n= 365) and MAUD (n= 7,398).

Among patients with obesity, 1,344 (82%) who initiated MAUD discontinued treatment before 365 days compared to 860 (53%) of those initiating a new GLP-1 RA (supplement, Table S2). After matching and IPCW, alcohol-related hospitalizations occurred in 8.1% (6.0% - 10.1%) of patients on a newer GLP-1 RA and 22.8% (17.5% - 27.7%) of patients on MAUD (Figure 1D), yielding a hazard ratio of 0.32 (0.23 - 0.43; e-value: 5.79) (Figure 2A).

### Negative control outcomes

Non-alcohol-related hospitalizations did not differ significantly between groups in the ADM and MAUD-T2D trials (Figure 2B, Figure 3A). In the AOM trial, however, treatment with a newer GLP-1 RA was associated with a lower risk of non-alcohol-related hospitalization relative to an older GLP-1 RA (0.69 [0.49 - 0.98], e-value: 2.3) and other AOMs (0.77 [0.63 - 0.93], e-value: 1.9) (Figure 2B). Newer GLP-1 RAs were also associated with lower risk of hospitalization in the MAUD-obesity trial (HR: 0.61 [0.44 - 0.85], e-value: 2.65) (Figure 2B).

**Figure 3:**
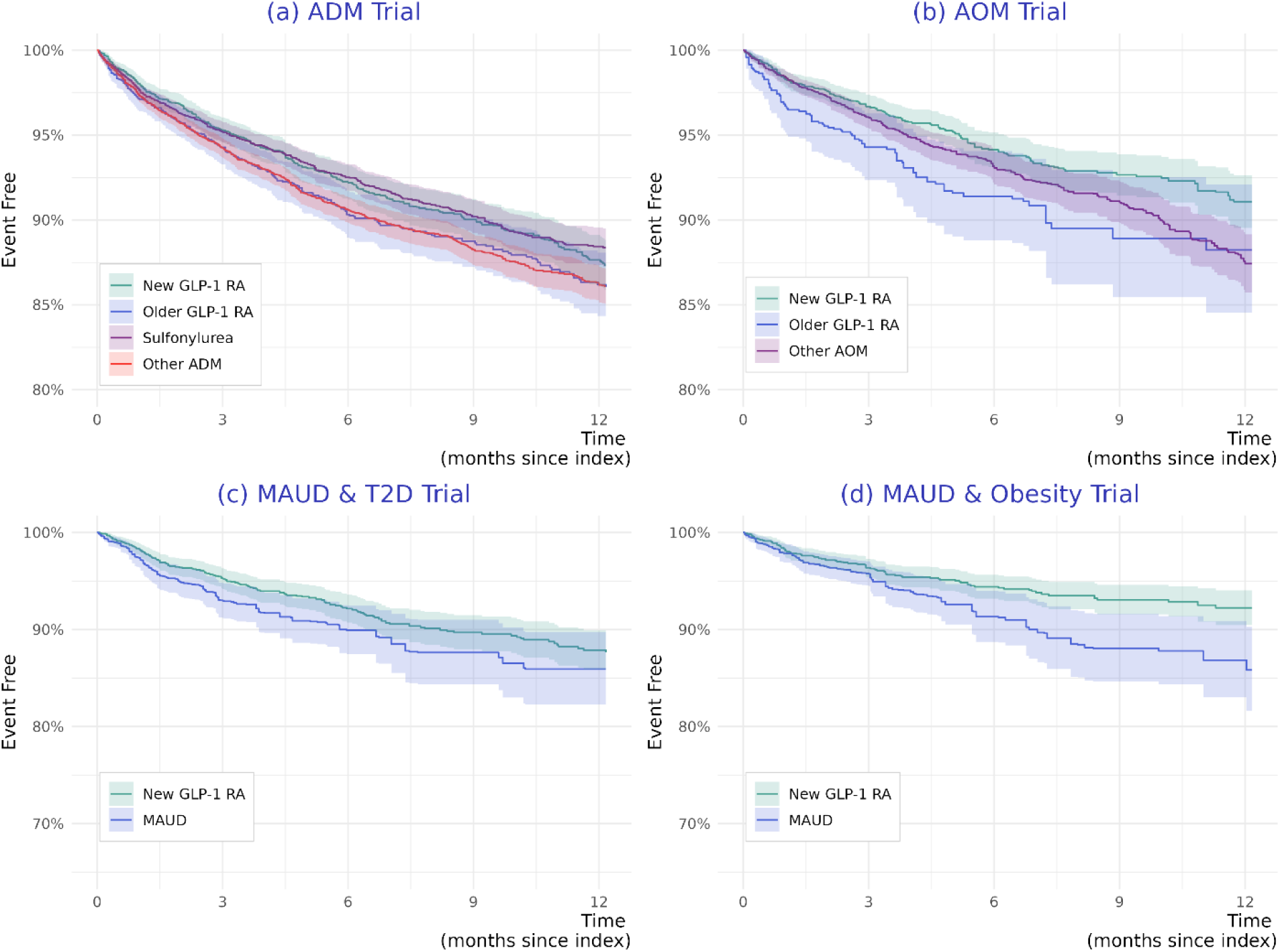
Time to non-alcohol-related hospitalization, balanced populations. Panel A: ADM trial, Panel B: AOM trial, Panel C: MAUD (T2D) trial, Panel D MAUD (Obesity) trial. Y-axis represents the probability of being event free (1-survival), x-axis represents time since initiation. Patients were censored at discontinuation, switching to a GLP-1 RA, or last encounter. Panels (a) and (b) use IPCW*ICTW weights. Panels (c) and (d) use IPCW weights on a matched population.

## Discussion

This target trial emulation study found a significantly lower rate of alcohol-related hospitalization for adults with AUD on semaglutide or tirzepatide relative to non-GLP-1 RA comparators across four clinically distinct cohorts with and without T2D. Effect estimates were similar in the ADM and AOM trials, and similar in the MAUD trials among patients with and without T2D.

This study extends the work of previous studies by including information on tirzepatide, which has been limited in previous studies, and using clearly defined comparators. Despite study design differences, findings observed in this study are consistent with previous reports noting reduced alcohol use for patients on a GLP-1 RA. The findings for newer GLP-1 RA in the ADM and AOM trials (HRs ranging from 0.59 to 0.73 depending on the trial) parallel effects observed in a nationwide Swedish registry study that evaluated the effect of semaglutide on AUD hospitalizations (HR: 0.64 [0.50 – 0.83])^14^ and a US study that evaluated recurrent AUD hospitalizations among patients with T2D (HR: 0.61 [0.50–0.75])^16^.

This study has several strengths. First, multiple trials were emulated to reflect clinically distinct target populations, increasing the accuracy and clinical relevance of the findings. Further, each trial included a large and generalizable population. This is important, as AUD is a heterogenous disease,^32^ and patients with AUD who participate in clinical trials often lack generalizability, due in part to comorbidity exclusions and instability.^33^ Third, inclusion of a negative control outcome further supports specific benefits of GLP-1 RAs on alcohol-related hospitalizations. While GLP-1 RAs were also associated with decreased risk of non-alcohol-related hospitalizations across trials including patients with obesity and without T2D, the magnitude of this effect was smaller than that for alcohol-related hospitalizations.

This study is also subject to limitations. First, AUD is likely under-captured in clinical practice, due in part to stigmatization.^34,35^ In addition, when it is documented, it may be recorded variably within and across health care systems. In fact, previous studies have noted lower reporting among practices in areas of high social deprivation.^36^ Second, before balancing, patients initiating a newer GLP-1 RA differed from those using other medications across all trials. While balancing approaches provided good balance on measured covariates, differences may exist with regard to unmeasured factors, particularly in trials of MAUD. Reassuringly, the e-values calculated in this study suggest that strong unmeasured confounding would be needed to negate the protective effect observed in MAUD trials. Third, while alcohol-related outcomes were based on alcohol-related diagnoses and tests for alcohol concentration during the hospitalization, misclassification and under-capture is possible. Finally, this study was limited to on-treatment analyses which censored patients at discontinuation or initiation of a comparator medication. As such, inclusion of post-baseline treatment information has the potential to introduce bias and limit generalizability. Importantly, on-treatment effects were selected because they tend to better approximate clinical trial results in real world analyses given differences between these populations.^37^ However, treatment effects observed in clinical trials may not reflect those expected in an average clinical setting, given well-described differences between trial versus non-trial populations.^33^

## Conclusion

In this target trial emulation study, patients with AUD who initiated a newer GLP-1 RA experienced a lower risk of alcohol-related hospitalization, with a similar effect between disease cohorts for T2D and for obesity. While further investigation is warranted, GLP-1 RAs may represent an effective treatment option for AUD, including those not actively seeking treatment.

## Data Availability

The data used in this study are available to all Truveta subscribers and may be accessed at studio.truveta.com.

