## Supplement for "GLP-1 Receptor Agonists vs Alternatives for Alcohol Use Disorder: A Multi-Target Trial Emulation"

#### Supplemental methods

Supplemental Figure S1: Study diagram ADM trial. Figure displays the application of study criteria and outcomes to data.

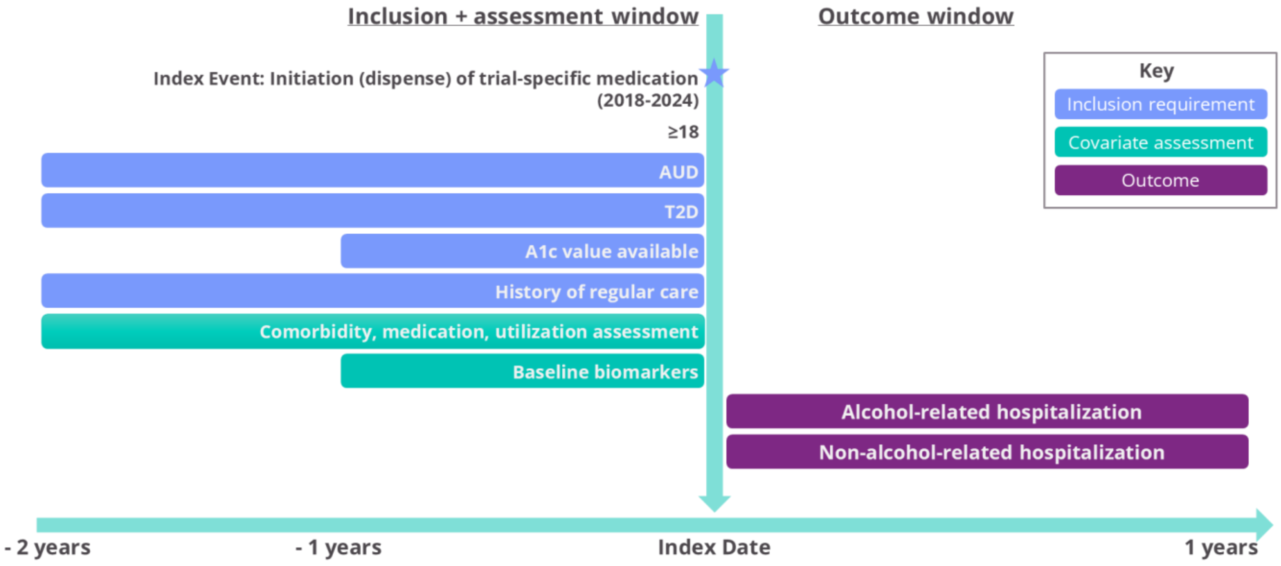

Supplemental Figure S2: Study diagram, AOM trial. Figure displays the application of study criteria and outcomes to data. *BMI ≥ 30 kg/m$^2$ refers to the most recent BMI in the 12 months before baseline.

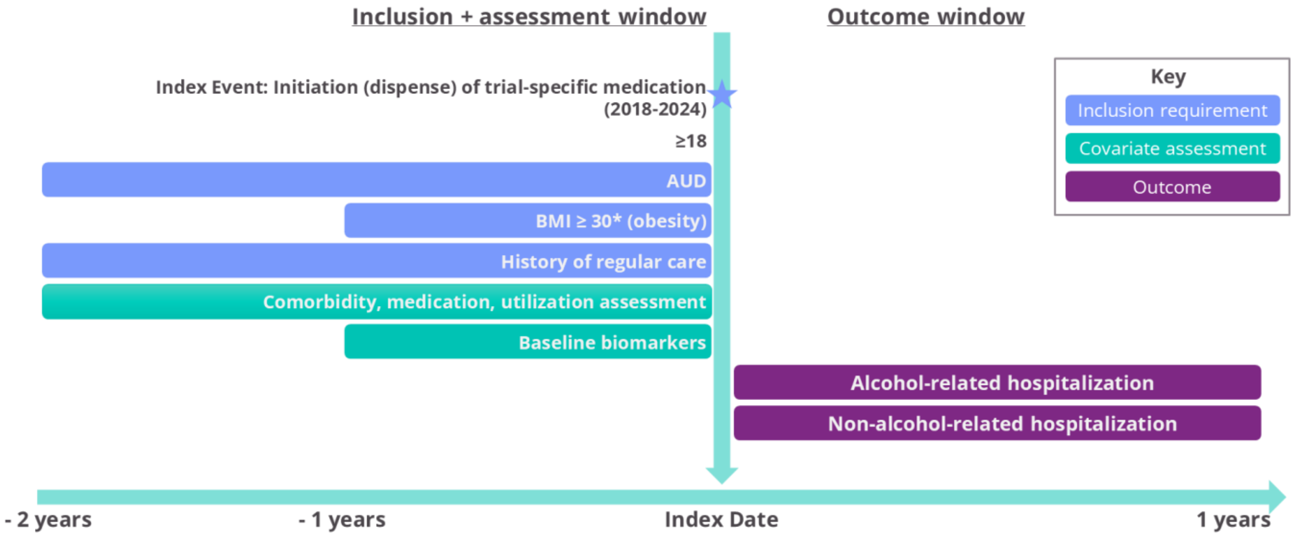

Supplemental Figure S3: Study diagram MAUD-T2D trial. Figure displays the application of study criteria and outcomes to data.

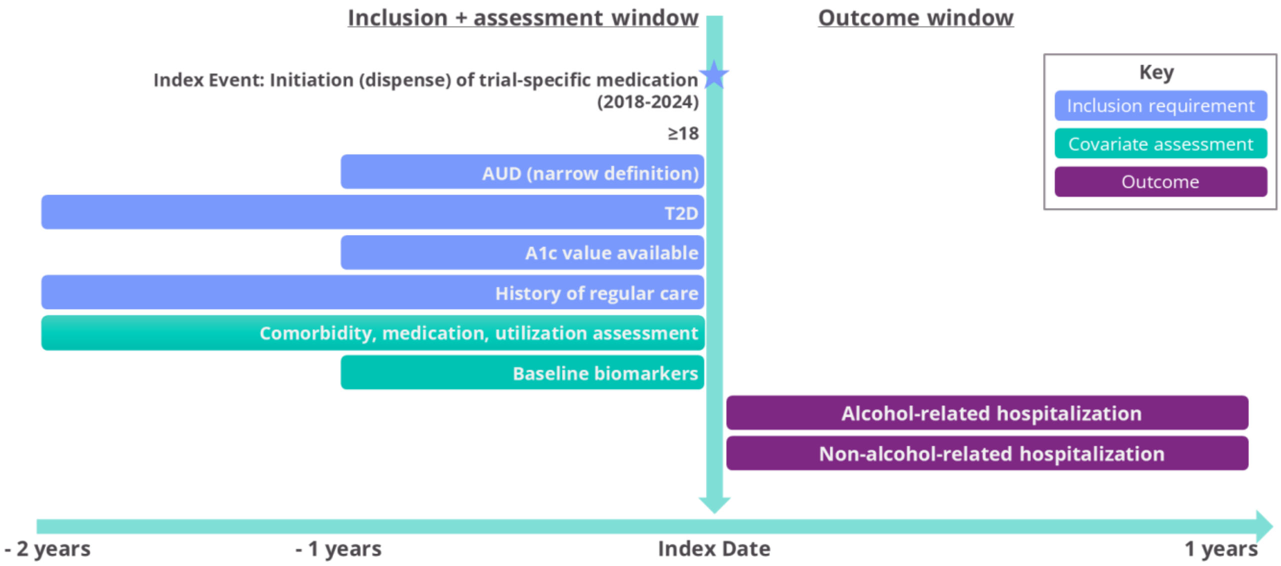

Supplemental Figure S4: Study diagram MAUD-obesity trial. Figure displays the application of study criteria and outcomes to data. *BMI ≥ 30 kg/m2 refers to the most recent BMI in the 12 months before baseline.

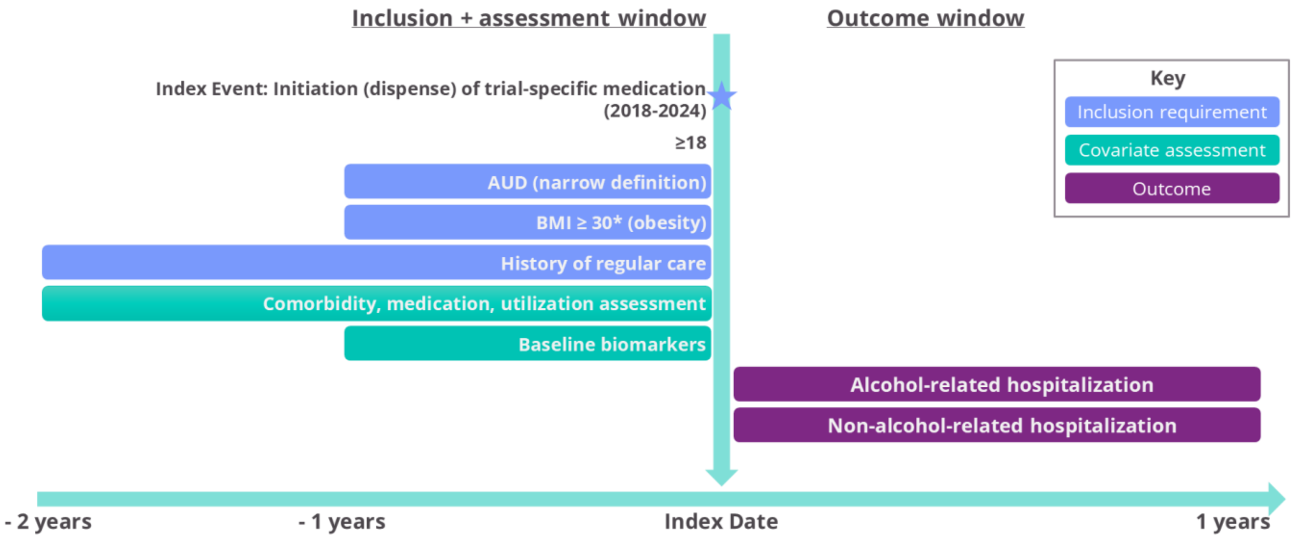

#### Negative control outcomes

Selection of negative control or tracer outcomes was challenging due to the wide-ranging known or hypothesized impacts of GLP-1 RAs and other study medications, and the potential for alcohol use to be a mediator for common tracer outcomes. For example, reduction in alcohol use could influence risky behaviors and social mixing, leading to changes in accidents and injuries, sexually transmitted infections, and other infectious diseases. Unfortunately, tracer outcomes expected to be wholly unrelated to the exposure, including hospitalizations for skin cancer, acute myeloid leukemia, and Huntington's disease, were insufficiently common to be used in this study. As such, a composite outcome of all hospitalizations not classified as alcohol-related was used.

#### Propensity of treatment models

Models of treatment selection varied slightly by trial.

The ADM and AOM models were fit using multinomial regression, given the presence of multiple treatment choices. Factors in both models included demographics (age [modeled using a nonlinear spline], sex, race, presence of any college education recorded), AUD-related factors (duration of AUD [years], time since the most recent AUD diagnosis [years], any alcohol-related hospitalization in the prior 1 years, any alcohol-related hospitalization in the prior 2 years, history of MAUD use [prescribing, dispensing, or administration], history of opioid use [prescribing, dispensing, or administration], history of opioid antagonist use [prescribing, dispensing, or administration]), utilization (number of OP office visits in the previous 0-12 months, number of OP office visits in the previous 13-24 months. Both included using second degree polynomials to account for non-linearity), and comorbidities (Elixhauser comorbidity score [modeled using nonlinear splines], cirrhosis, chronic kidney disease, chronic liver disease, cardiovascular disease , obstructive sleep apnea, hypertension, hyperlipidemia, major depressive disorder, opioid use disease, other substance use disorder [excluding alcohol and opioid]). The ADM treatment selection model trial also included A1c, obesity comorbidity, and history of metformin, with A1c modeled flexibility that used a nonlinear spline with 4 knots. The AOM treatment selection model trial additionally included BMI, with modeled flexibility using a nonlinear spline with 4 knots.

MAUD treatment selection models were fit using logistic regression, given binary treatment choices (newer GLP-1 RA or MAUD). Given a desire to generalize to a population seeking treatment for AUD, treatment selection of MAUD served as the outcome. In addition to the variables above, additional variables were included to adjust for AUD in the relatively more severe population having more than 5 hospitalizations in the previous year, more than 10 hospitalizations in the previous year, and more than 5 outpatient visits in the previous year.

#### Supplemental Results

Supplemental Figure S5: Distribution of index time by trial. Panel A: ADM trial, Panel B: AOM trial, Panel C: MAUD (T2D) trial, Panel D MAUD (Obesity) trial.

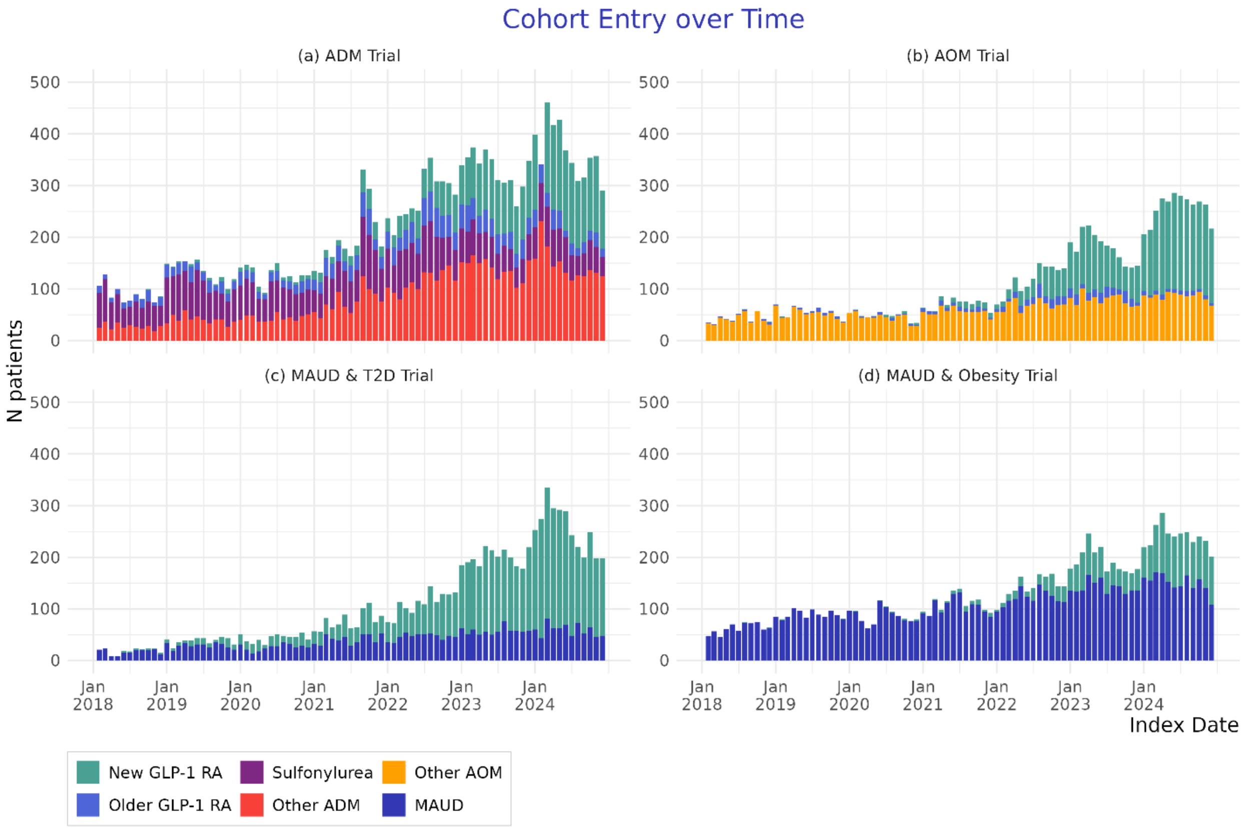

Table S1: Characteristics of patients at baseline, after balancing. Quantitative variables are expressed as mean (standard deviation). Categorical variables are expressed as number (percentage). Utilization counts are expressed as median (IQR). Durations refers to the time (years) since first evidence. Other race includes Asian, American Indian or Alaska Native, Native Hawaiian or Other Pacific Islander, Other Race, Unknown, Declined to answer. Abbreviations: AUD = alcohol use disorder, AST = aspartate aminotransferase, ALT = alanine aminotransferase, A1c = glycated hemoglobin, BMI = body mass index, DPP4 = dipeptidyl peptidase 4 inhibitor, MAUD = medications for alcohol use disorder (includes acamprosate, disulfiram, and naltrexone), SD = standard deviation, SGLT2i = sodium/glucose cotransporter-2 inhibitor, SSRI = selective serotonin reuptake inhibitor, T2D = Type 2 diabetes.

| | **ADM trial N = 18,515** | | | | **AOM trial N = 9,256** | | | **MAUD-T2D trial N =3,178** | | **MAUD-obesity trial N = 3,276** | |
|---|---|---|---|---|---|---|---|---|---|---|---|
| Characteristic | Newer GLP-1 RA N = 8,287 | Older GLP-1 RA N = 1,700 | Sulfonyl urea N = 3,562 | Other ADM N = 4,967 | Newer GLP-1 RA N = 4,398 | Older GLP-1 RA N = 555 | Other AOM N = 4,304 | Newer GLP-1 RA N = 1,607 | MAUD N = 1,571 | Newer GLP-1 RA N = 1,622 | MAUD N = 1,654 |
| Age | 59.1 (11.6) | 56.2 (11.5) | 60.1 (11.9) | 60.9 (11.8) | 50.6 (12.8) | 52.2 (12.7) | 49.6 (13.1) | 56.1 (11.4) | 56.6 (11.6) | 52.0 (12.5) | 51.7 (12.7) |
| Female | 2,485 (30%) | 701 (41%) | 858 (24%) | 1,212 (24%) | 2,618 (60%) | 306 (55%) | 2,641 (61%) | 483 (30%) | 485 (31%) | 853 (53%) | 847 (51%) |
| Race | | | | | | | | | | | |
| Black | 862 (10%) | 217 (13%) | 326 (9%) | 632 (13%) | 381 (9%) | 55 (10%) | 335 (8%) | 181 (11%) | 182 (12%) | 126 (8%) | 109 (7%) |
| White | 6,063 (73%) | 1,203 (71%) | 2,475 (70%) | 3,403 (69%) | 3,384 (77%) | 429 (77%) | 3,314 (77%) | 1,171 (73%) | 1,160 (74%) | 1,267 (78%) | 1,304 (79%) |
| Other or unknown | 1,362 (16%) | 280 (16%) | 760 (21%) | 931 (19%) | 632 (14%) | 71 (13%) | 654 (15%) | 255 (16%) | 229 (15%) | 229 (14%) | 241 (15%) |
| Ethnicity | | | | | | | | | | | |
| Hispanic or Latino | 1,010 (12%) | 173 (10%) | 418 (12%) | 515 (10%) | 375 (9%) | 45 (8%) | 364 (8%) | 207 (13%) | 185 (12%) | 126 (8%) | 97 (6%) |
| Not Hispanic or Latino | 6,983 (84%) | 1,459 (86%) | 2,906 (82%) | 4,157 (84%) | 3,835 (87%) | 488 (88%) | 3,789 (88%) | 1,339 (83%) | 1,287 (82%) | 1,423 (88%) | 1,487 (90%) |
| Unknown | 295 (4%) | 68 (4%) | 238 (7%) | 295 (6%) | 188 (4%) | 22 (4%) | 150 (3%) | 61 (4%) | 99 (6%) | 73 (5%) | 70 (4%) |

| | | | | | | | | | | | |
|---|---|---|---|---|---|---|---|---|---|---|---|
| Any college on record | 2,692 (32%) | 647 (38%) | 910 (26%) | 1,361 (27%) | 2,467 (56%) | 257 (46%) | 2,428 (56%) | 603 (38%) | 545 (35%) | 871 (54%) | 863 (52%) |
| Index medication | | | | | | | | | | | |
| Semaglutide | 6,917 (83%) | - | - | - | 3,213 (73%) | - | - | 1,318 (82%) | - | 1,195 (74%) | - |
| Tirzepatide | 1,370 (17%) | - | - | - | 1,184 (27%) | - | - | 289 (18%) | - | 427 (26%) | - |
| Dulaglutide | - | 1,209 (71%) | - | - | - | 184 (33%) | - | - | - | - | - |
| Liraglutide | - | 401 (24%) | - | - | - | 352 (64%) | - | - | - | - | - |
| Exenatide | - | 85 (5%) | - | - | - | 15 (3%) | - | - | - | - | - |
| Lixisenatide | - | 5 (0%) | - | - | - | 2 (0%) | - | - | - | - | - |
| Sulfonylurea | - | - | 3,562 (100%) | - | - | - | - | - | - | - | - |
| DPP4 | - | - | - | 1,223 (25%) | - | - | - | - | - | - | - |
| SGLT2i | - | - | - | 3,744 (75%) | - | - | - | - | - | - | - |
| Phentermine | - | - | - | - | - | - | 1,237 (29%) | - | - | - | - |
| Phentermine-topiramate | - | - | - | - | - | - | 178 (4%) | - | - | - | - |
| Topiramate | - | - | - | - | - | - | 2,874 (67%) | - | - | - | - |
| Orlistat | - | - | - | - | - | - | 15 (0%) | - | - | - | - |
| Acamprosate | - | - | - | - | - | - | - | - | 352 (22%) | - | 383 (23%) |
| Disulfiram | - | - | - | - | - | - | - | - | 150 (10%) | - | 238 (14%) |
| Naltrexone | - | - | - | - | - | - | - | - | 1,064 (68%) | - | 1,011 (61%) |
| sNaltrexone bupropion | - | - | - | - | - | - | - | - | 4 (0%) | - | 22 (1%) |
| Baseline BMI | 35.9 (7.3) | 36.4 (8.2) | 31.9 (7.2) | 32.3 (7.3) | 37.9 (6.5) | 39.8 (7.4) | 37.5 (6.6) | 35.7 (7.8) | 31.8 (7.0) | 37.6 (6.0) | 37.3 (6.8) |
| Unknown | 1,648 | 365 | 728 | 1,050 | | | | 291 | 236 | | |

| | | | | | | | | | | | |
|---|---|---|---|---|---|---|---|---|---|---|---|
| Baseline ALT | 35.8 (25.8) | 35.8 (24.6) | 36.9 (27.2) | 33.5 (24.3) | 35.1 (26.0) | 35.1 (23.9) | 33.3 (24.8) | 35.9 (27.6) | 43.2 (33.5) | 36.3 (26.8) | 42.0 (32.6) |
| Unknown | 646 | 171 | 327 | 432 | 868 | 155 | 994 | 98 | 95 | 278 | 419 |
| Baseline AST | 31.0 (24.2) | 30.1 (22.8) | 32.7 (25.8) | 30.8 (23.7) | 29.8 (23.2) | 29.4 (22.1) | 29.3 (23.0) | 30.9 (24.1) | 47.1 (42.9) | 30.7 (25.5) | 43.0 (39.7) |
| Unknown | 600 | 176 | 332 | 423 | 859 | 152 | 995 | 96 | 88 | 278 | 419 |
| Comorbidities | | | | | | | | | | | |
| Elixhauser comorbidity score | 10.5 (10.9) | 9.8 (10.6) | 10.5 (11.1) | 12.8 (11.6) | 4.4 (10.1) | 5.8 (11.1) | 4.1 (10.1) | 10.3 (11.1) | 10.7 (11.6) | 4.3 (10.1) | 4.6 (10.3) |
| Cirrhosis | 890 (11%) | 169 (10%) | 441 (12%) | 613 (12%) | 239 (5%) | 51 (9%) | 206 (5%) | 232 (14%) | 209 (13%) | 114 (7%) | 137 (8%) |
| Chronic kidney disease | 2,507 (30%) | 493 (29%) | 1,046 (29%) | 1,869 (38%) | 473 (11%) | 74 (13%) | 463 (11%) | 453 (28%) | 468 (30%) | 173 (11%) | 166 (10%) |
| Chronic liver disease | 1,267 (15%) | 237 (14%) | 624 (18%) | 839 (17%) | 374 (9%) | 70 (13%) | 352 (8%) | 324 (20%) | 316 (20%) | 161 (10%) | 202 (12%) |
| Cardiovascular disease | 2,874 (35%) | 542 (32%) | 1,159 (33%) | 2,015 (41%) | 417 (9%) | 73 (13%) | 407 (9%) | 501 (31%) | 453 (29%) | 148 (9%) | 157 (9%) |
| Obesity | 7,230 (87%) | 1,507 (89%) | 2,845 (80%) | 4,107 (83%) | 4,398 (100%) | 555 (100%) | 4,304 (100%) | 1,351 (84%) | 1,314 (84%) | 1,622 (100%) | 1,654 (100%) |
| Obstructive sleep apnea | 3,264 (39%) | 744 (44%) | 994 (28%) | 1,793 (36%) | 1,462 (33%) | 219 (39%) | 1,335 (31%) | 588 (37%) | 559 (36%) | 554 (34%) | 532 (32%) |
| Hypertension | 7,278 (88%) | 1,460 (86%) | 3,098 (87%) | 4,448 (90%) | 2,687 (61%) | 370 (67%) | 2,502 (58%) | 1,390 (86%) | 1,361 (87%) | 1,035 (64%) | 1,035 (63%) |
| Hyperlipidemia | 6,933 (84%) | 1,406 (83%) | 2,886 (81%) | 4,193 (84%) | 2,216 (50%) | 281 (51%) | 2,085 (48%) | 1,281 (80%) | 1,227 (78%) | 859 (53%) | 873 (53%) |
| Major depressive disorder | 2,633 (32%) | 710 (42%) | 996 (28%) | 1,423 (29%) | 1,669 (38%) | 175 (32%) | 1,800 (42%) | 614 (38%) | 603 (38%) | 564 (35%) | 593 (36%) |
| Opioid use disorder | 505 (6%) | 143 (8%) | 223 (6%) | 275 (6%) | 291 (7%) | 40 (7%) | 331 (8%) | 119 (7%) | 120 (8%) | 102 (6%) | 112 (7%) |
| Other substance use disorder | 758 (9%) | 226 (13%) | 329 (9%) | 533 (11%) | 505 (11%) | 73 (13%) | 598 (14%) | 206 (13%) | 201 (13%) | 175 (11%) | 202 (12%) |
| Previous or concurrent medications | | | | | | | | | | | |
| History of metformin | 5,692 (69%) | 1,176 (69%) | 2,613 (73%) | 3,276 (66%) | 414 (9%) | 100 (18%) | 195 (5%) | 1,011 (63%) | 985 (63%) | 179 (11%) | 49 (3%) |
| History of MAUD | 538 (6%) | 151 (9%) | 174 (5%) | 280 (6%) | 776 (18%) | 82 (15%) | 789 (18%) | - | - | - | - |

| | | | | | | | | | | | |
|---|---|---|---|---|---|---|---|---|---|---|---|
| History of opioid | 3,811 (46%) | 815 (48%) | 1,599 (45%) | 2,477 (50%) | 1,747 (40%) | 226 (41%) | 1,804 (42%) | 865 (54%) | 775 (49%) | 610 (38%) | 634 (38%) |
| History of opioid antagonist | 1,378 (17%) | 290 (17%) | 527 (15%) | 839 (17%) | 930 (21%) | 115 (21%) | 941 (22%) | 595 (37%) | 589 (37%) | 265 (16%) | 256 (15%) |
| History of SSRI | 2,681 (32%) | 720 (42%) | 1,065 (30%) | 1,480 (30%) | 2,044 (46%) | 250 (45%) | 2,068 (48%) | 624 (39%) | 771 (49%) | 712 (44%) | 842 (51%) |
| AUD severity | | | | | | | | | | | |
| AUD duration (years) | 3.1 (3.0) | 3.1 (3.2) | 2.8 (2.8) | 3.1 (3.4) | 2.8 (2.8) | 3.1 (3.0) | 2.7 (2.7) | 3.2 (3.1) | 3.3 (3.1) | 3.2 (3.2) | 3.1 (3.2) |
| Dates with AUD diagnosis, previous 2 years (n) | 3 (1-9) | 4 (2-11) | 3 (1-9) | 4 (1-10) | 3 (1-8) | 3 (1-8) | 3 (2-9) | 5 (2-12) | 5 (3-12) | 4 (2-9) | 4 (2-8) |
| Most recent AUD diagnosis (years before index) | 0.4 (0.5) | 0.4 (0.5) | 0.4 (0.5) | 0.4 (0.5) | 0.5 (0.5) | 0.5 (0.6) | 0.4 (0.5) | 0.2 (0.2) | 0.2 (0.2) | 0.2 (0.3) | 0.2 (0.3) |
| Had AUD (narrow definition) | 6,917 (83%) | 1,503 (88%) | 3,181 (89%) | 4,356 (88%) | 3,470 (79%) | 493 (89%) | 3,731 (87%) | 1,607 (100%) | 1,571 (100%) | 1,622 (100%) | 1,654 (100%) |
| Had any alcohol-related hospitalization in prior 2 years | 2,303 (28%) | 475 (28%) | 1,074 (30%) | 1,683 (34%) | 1,103 (25%) | 152 (27%) | 1,183 (27%) | 695 (43%) | 675 (43%) | 401 (25%) | 398 (24%) |
| Years in Truveta (since first encounter) | 12.5 (12.0) | 12.1 (11.6) | 11.3 (10.8) | 11.5 (11.1) | 12.7 (11.5) | 14.4 (12.5) | 13.0 (12.1) | 11.4 (11.5) | 13.2 (12.1) | 12.3 (10.9) | 14.7 (12.8) |
| Outpatient visits in previous 0-12 months (n) | 9 (5-19) | 11 (5-23) | 8 (4-16) | 10 (5-21) | 7 (4-15) | 8 (4-18) | 8 (4-18) | 10 (5-20) | 10 (5-19) | 7 (3-15) | 7 (3-15) |
| Outpatient visits in previous 13-24 months (n) | 7 (2-15) | 10 (3-21) | 6 (2-14) | 7 (2-16) | 5 (2-13) | 6 (2-14) | 6 (2-15) | 6 (2-15) | 6 (2-15) | 4 (1-11) | 5 (1-12) |

Supplemental Table S2: Reasons for censoring before 365 days by balanced populations. This table considers available follow-up <365 days, regardless of events. Patients may have experienced events before these censoring events. Patients who started on a newer GLP-1 RA were not censored at the time of addition of a different medication.

| Trial | Group | Discontinuation | Started GLP-1 RA / Newer GLP-1 RA | Last encounter |
|---|---|---|---|---|
| **ADM trial** | Overall | 8663 (47%) | 1328 (7%) | 2742 (15%) |
| | Newer GLP-1 RA | 1791 (44%) | - | 1041 (26%) |
| | Older GLP-1 RA | 1108 (51%) | 158 (7%) | 162 (7%) |
| | Sulfonylurea | 2681 (51%) | 418 (8%) | 487 (9%) |
| | Other ADM | 3083 (44%) | 752 (11%) | 1052 (15%) |
| **AOM trial** | Overall | 5868 (63%) | 226 (2%) | 1649 (18%) |
| | Newer GLP-1 RA | 1830 (52%) | - | 1112 (32%) |
| | Older GLP-1 RA | 376 (62%) | 73 (12%) | 51 (8%) |
| | Other AOM | 3662 (71%) | 153 (3%) | 486 (9%) |
| **MAUD-T2D trial** | Overall | 2063 (65%) | 57 (2%) | 453 (14%) |
| | Newer GLP-1 RA | 773 (49%) | - | 347 (22%) |
| | MAUD | 1290 (81%) | 57 (4%) | 106 (7%) |
| **MAUD-obesity trial** | Overall | 2204 (67%) | 27 (1%) | 612 (19%) |
| | Newer GLP-1 RA | 860 (53%) | - | 490 (30%) |
| | MAUD | 1344 (82%) | 27 (2%) | 122 (7%) |

Supplemental Figure S6: Time to alcohol-related hospitalization on unadjusted, unweighted data. Panel A: ADM trial, Panel B: AOM trial, Panel C: MAUD (T2D) trial, Panel D MAUD (Obesity) trial.

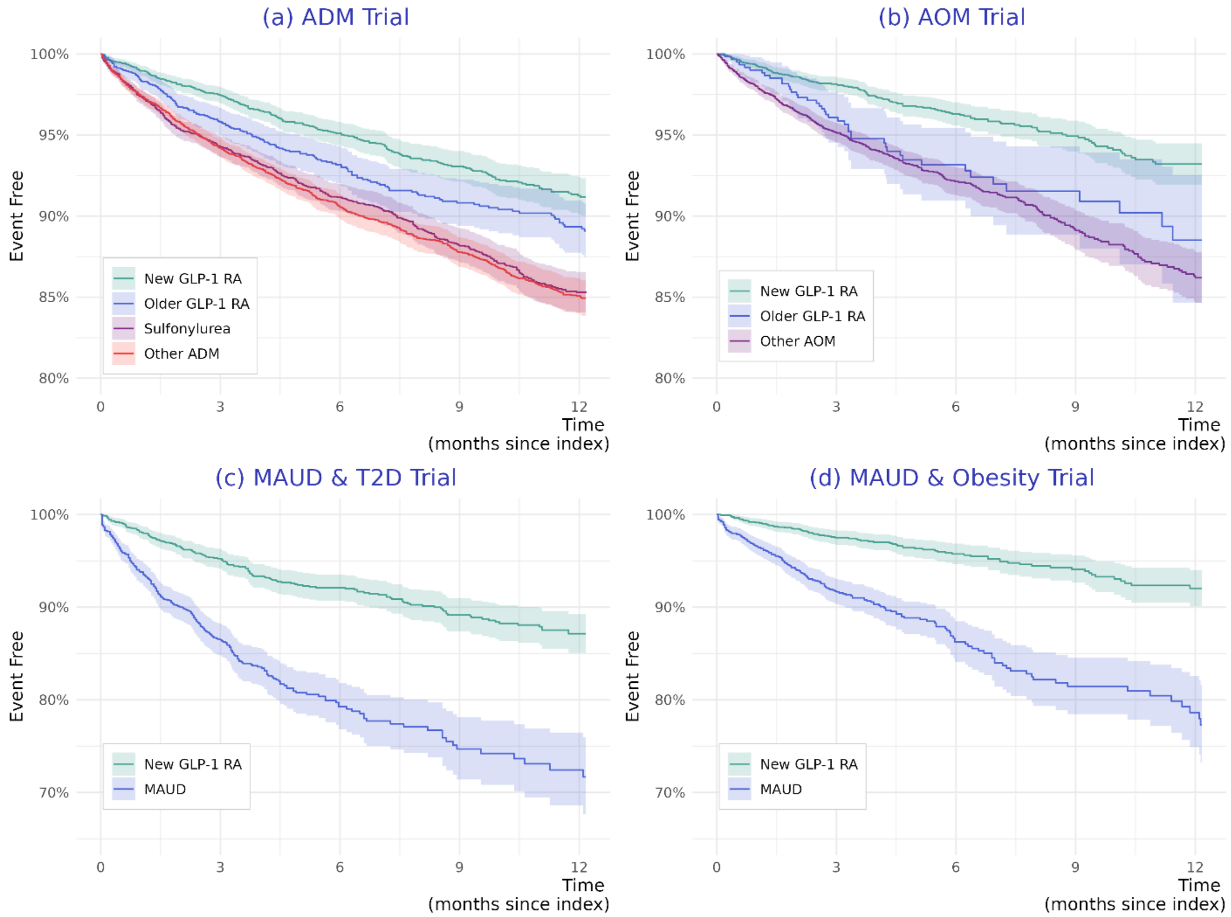

### Code definitions

| Code System | Concept Code |
|---|---|
| **Newer GLP-1 RA** | |
| NDC | 00169413001, 001694524, 00002145701, 00169450514, 00169451701, 00169450101, 00002147101, 001694525, 00002148401, 001694505, 00002145780, 50090513800, 00002146080, 001694517, 00169452414, 00002149580, 001694772, 001694132, 00002150661, 00002147180, 00169452501, 000021457, 00169477290, 00169477211, 500905824, 001694130, 50090582400, 000021506, 00169413290, 00169452401, 00169451714, 00002148480, 00002150680, 001694136, 00169450114, 00169413297, 00169477212, 000021484, 001694501, 70518214300, 000021471, 000021495, 00002149501, 00002150601, 00169452594, 00169413212, 500905949, 50090513900, 00169452590, 00169413013, 00002146001, 00169452514, 705182143, 000021460, 00169477297, 500905138, 50090594900, 500905139, 00169413211, 00169413611, 00169413602, 00169450501, 00002247180, 000022423, 00002247101, 000022506, 00002250601, 000022495, 00002115201, 00002200201, 000022471, 00002024301, 000022460, 00002142301, 000023002, 000022457, 000022214, 00002246080, 000022484, 000022340, 000021243, 000021152, 00002248480, 00002249580, 00002250680, 00002015201, 00002300201, 00002242301, 00002221401, 00002245701, 00002246001, 00002248401, 00002124301, 00002249501, 00002245780, 00169418197, 00169418190, 50090605100, 001694181, 00169418113, 00002134001, 00002234001, 00002250661, 00002121401, 500906051, 00169418103 |
| RxNorm | 2601755, 2601772, 2601768, 2601758, 2601771, 2601775, 2679322, 2601757, 2679316, 2601754, 2679327, 2679325, 2601774, 2601776, 2679319, 2679320, 2679317, 2679321, 2679328, 2679326, 2679324, 2601777, 2679318, 2679323, 2644401, 2601785, 2601781, 2601746, 2669716, 2601736, 2601737, 2669708, 2669705, 2601730, 2669711, 2601743, 2601784, 2601773, 2644417, 2669706, 2669715, 2644396, 2601745, 2601761, 2601742, 2669720, 2644407, 2669714, 2601759, 2601764, 2601747, 2669713, 2601770, 2644403, 2669702, 2669717, 2601778, 2601780, 2644415, 2644399, 2601783, 2601734, 2669709, 2601760, 2601762, 2669707, 2644419, 2669718, 2644411, 2644413, 2644409, 2601765, 2669703, 2601744, 2669722, 2669710, 2669721, 2669712, 2644405, 2601767, 2669719, 2669704, 2601763, 2601731, 2644410, 2644400, 2644397, 2644414, 2644420, 2644398, 2644395, 2644402, 2644408, 2644406, 2644412, 2644418, 2644404, 2644416, 2619154, 2553700, 2553508, 2553602, 2619153, 2553803, 2553905, 2553505, 2553903, 2554104, 2553503, 2553608, 2553605, 2553504, 2553603, 2553902, 2554103, 2553506, 2554106, 2599365, 2599366, 2599364, 2601769, 2601723, 2601766, 2601756, 1991307, 2553502, 2398844, 1991309, 1991308, 2398842, 1991317, 1991311, 1991310 |
| **Older GLP-1 RA** | |
| NDC | 500906453, 500906456, 500906571, 50090657100, 00169291197, 00002021007, 003106530, 66914103505, 00169406097, 545680434, 000021433, 66780021902, 00024574000, 003106520, 54568043463, 00310651201, 00310654004, 54868538402, 00002318280, 00310653001, 50090425700, 500905467, 54868538401, 003106512, 500903484, 001694060, 000245761, 00024576101, 00169406012, 00002143401, 00024574502, 00024574702, 000245745, 000021434, 50090285300, 66780021008, 00169280090, 667800219, 54568043471, 00024576302, 54868538400, 00024576102, 54568043363, 003106540, 00169291115, 66780021201, 00310653085, 00002143361, 500904503, 00002223661, 00310651285, 00002223601, 548685384, 66914103504, 66029021007, 66780022601, 00169406090, 66780021009, 00002318201, 003106524, 00002143480, 500902853, 00169406013, 00002021008, 00002021009, 00002143461, 00310654001, 667800210, 00024576105, 66780021007, 54569650700, 50090348300, 66780021904, 500904257, 00024574101, 00169280097, 000245747, 667800226, 50090546700, 00169406098, 545680433, 00169280013, 00002318261, 00310654085, 00002143380, 000022236, 00310652401, 66029021008, 667800212, 500903483, 00169291190, 00310652004, 00169280015, 000023182, 00310653004, 00169406099, 001692911, 54568043371, 00002223680, 50090450300, 50090348400, 00002143301, 001692800, 68258894802, 68258894701, 50090645300, 00480366720, 00480366722, 50090645600, 00480366719 |

| Code System | Concept Code |
|---|---|
| RxNorm | 1551296, 1242964, 60548, 1598264, 604751, 897123, 1440051, 475968, 1803887, 1551291, 1858996, 897120, 1858998, 744863, 1803889, 1551307, 1803886, 1653611, 1860170, 1803892, 1803897, 2395778, 1803894, 1803888, 1163790, 1803891, 1860168, 1803903, 847914, 1653613, 1544919, 1860165, 1803898, 2395787, 2395786, 2395781, 1551292, 1860164, 1169415, 1858999, 1242965, 2395784, 1598265, 1990867, 1551297, 897122, 1803896, 1803895, 847908, 1990868, 1649584, 1859002, 1990866, 1360495, 2395785, 1360105, 1653600, 2395783, 1858994, 1440056, 1186578, 1990871, 1440053, 1649586, 2395780, 1653594, 847915, 1653625, 1598268, 1859001, 1360454, 1242967, 1359979, 1551295, 1551305, 2395779, 1860166, 1858997, 1359802, 1803890, 1551301, 1858993, 1859000, 1551303, 847916, 1598269, 2395782, 1860167, 1598267, 1544916, 1860173, 847917, 1990869, 1990870, 1163230, 1858992, 1860171, 1653610, 1860174, 1803893, 2395777, 1860172, 1858995, 1990864, 1440052, 1653614, 1653619, 1551293, 897126, 744862, 2395776, 1551308, 1653616, 1551304, 1242963, 1653597, 1551306, 1551300, 1544920, 1242961, 847910, 897124, 1860169, 1990865, 847913, 1858991, 1242968, 1727493, 1551299, 1803885, 1803902, 1551302, 1544918, 1359640, 847911, 1242966, 897121, 1440055, 1440054, 1598266, 1551298, 744865, 1242962, 604753, 604748, 847912, 604752, 744864, 1440057, 1551294, 604749, 1544917, 604754, 604750, 897125, 847909 |
| SNOMED CT | 714081009, 763570007, 416525003, 714080005, 440246006, 708808004, 764330009, 416859008, 438958002, 417734003, 779728007, 1010537001, 775712006, 1237218000, 1010541002, 1155638004, 1010538006, 775913009, 776560001, 1155637009, 1155639007, 1010536005, 1010539003, 1010540001 |
| **Other ADM** | |

| | |
|---|---|
| NDC | 70518365201, 71335236701, 50090632200, 50090718702, 71335961403, 70518365202, 72888011605, 71335961405, 50090718700, 70518112404, 50090647303, 72189045390, 50090619601, 50090641503, 70934066660, 68071354806, 50090619608, 50090642601, 68071283908, 70518368800, 71335961401, 82868003530, 71335961404, 71335961406, 68788824301, 50090619609, 53002446103, 50090632201, 51655096626, 71610068080, 68788834101, 71335961402, 68788824309, 68788863408, 713359614, 71335965703, 60760079390, 71335974203, 71610066860, 53002446106, 82804006530, 68788824303, 70518378600, 68788824306, 50090632202, 60687076811, 68788824308, 680712839, 500906322, 51655049226, 68071283906, 71335961407, 50090709000, 72189048560, 50090709001, 72789027501, 60687069001, 71610066853, 727890275, 72888011530, 60687070101, 71610066870, 514070620, 71610068060, 71610066880, 71610066894, 71610068045, 68071283909, 60687070111, 60687069011, 716100668, 606870690, 68788850803, 51407062010, 71610066892, 50090619602, 716100680, 43063094630, 50090100602, 71335240901, 50090647301, 71335235303, 68788843206, 596510780, 70518378601, 68788851201, 72603011602, 68788851206, 71610068073, 50090647300, 51655012026, 71205077390, 70518389801, 71335236702, 71921021501, 70518366501, 51655018025, 50090676500, 71335965502, 53002417103, 70518368801, 50090317202, 71335974202, 51655048926, 68788848703, 68788848709, 530024111, 59651078299, 687888512, 68788855003, 70518414000, 72189048590, 70518372200, 72888011630, 82868003590, 68788834106, 71335965404, 53002411106, 62135058430, 596510782, 68788863409, 62135058590, 71335965501, 50090641500, 70518402500, 70518362702, 50090674602, 71921021601, 50090639402, 709340878, 72888011500, 50090697500, 50090642600, 71335235305, 68788855009, 51655098252, 59651078101, 68001058500, 71335236705, 530024461, 62135058330, 68788834108, 71205077330, 70518389800, 70518055205, 50090684900, 51655038352, 59651078201, 71335235301, 68788855001, 71921021450, 68788834306, 51655098326, 53002411103, 687888432, 71610068098, 68788850806, 51655048952, 52817038550, 62135073260, 59651078105, 51655018052, 71921021550, 687888487, 72888011730, 68788855008, 50090676502, 596510781, 68001058403, 70518403900, 68001058400, 62135073360, 70518389600, 68788855006, 50090676501, 50090684801, 71335965401, 82868002530, 50090642603, 50090647302, 72162221500, 59651078030, 82804006590, 62135073160, 70518364501, 68788850809, 43806003570, 00093834493, 00839794016, 00378111001, 00686029320, 00009344901, 38245047749, 11726041166, 43353037945, 00039022307, 00093943301, 43063043386, 10768715001, 42571010105, 00894625503, 00223208101, 00056071185, 43063012230, 00440758790, 00068320161, 00302718222, 165710773, 00179151330, 43353037960, 00725206910, 33261083530, 332610209, 33261096190, 12280002700, 42571010405, 00480725652, 00615359639, 43063043398, 23155011601, 00173315113, 00088022302, 00364257705, 43063012090, 00615657631, 16590041860, 005910900, 00069155066, 00223208205, 00009017111, 00405536101, 00093725561, 00172571260, 12634072974, 43353020794, 000490174, 38245043310, 00894625501, 00781504510, 00686021701, 00039005101, 26053016301, 00615796830, 00603374621, 167290140, 001151648, 00088022202, 00555041704, 21695047060, 12634079069, 433530379, 12634079085, 43547039450, 10544022190, 00228275150, 216950568, 00378401101, 00378034093, 00009372707, 00005306923, 00480745601, 12634079098, 00855203650, 43353020780, 00143991805, 35356011330, 000390223, 35356012190, 001151742, 24236018002, 12634079071, 43353036970, 00536474402, 430630946, 00349841901, 00677158001, 00781517105, 21695074760, 42291042910, 23155005701, 167140872, 00039005211, 00781517101, 00069155057, 00093725510, 00088022247, 001730844, 216950993, 10544019360, 12634079078, 000876072, 21695046990, 00088022227, 231550057, 00039005201, 23155011701, 33358016030, 001439920, 12783007411, 00247229630, 00247228990, 00536569801, 10544019230, 00719197510, 000937255, 00781145301, 00591046001, 00068121161, 42571010301, 00182264705, 00378034210, 005910845, 00904023461, 12634079081, 00378034010, 00615359631, 43353089518, 00182199589, 00349841827, 00007314813, 00172365010, 00182264710, 458020770, 00039022110, 00536564106, 00677095501, 00615558539, 00378110505, 00172365070, 00904792540, 43063003490, 11845110003, 430630121, 12634079042, 12634072959, 001790205, 353560932, 12634079054, 17856084402, 00405502501, 00781145201, 000938343, 000938036, 00182199500, 00615450939, 10544019260, 43353058280, 00179178972, |

| | |
|---|---|
| | 38245038120, 00121092991, 00102313501, 009046123, 00378313101, 00093571119, 00603375528, 33358015730, 35356099560, 00839675506, 00039005103, 00093725501, 43063043315, 00039005005, 00039005252, 16729014016, 00536473910, 00615254510, 00039005250, 00480571005, 00172297965, 00247127006, 00009017112, 00228275350, 35356093190, 00049017030, 00781194213, 425710104, 105440579, 19458033601, 00182122105, 23155005610, 24236017202, 12634072994, 00173084513, 00068121101, 10370074505, 00009017101, 00009017109, 00172571210, 10544023230, 00440757020, 35356089660, 00832096813, 006158407, 00172365060, 00364072290, 33358016190, 425710103, 00725207001, 00615796939, 00179178970, 00115174301, 00093571201, 12280002800, 00536575101, 00009034102, 167140871, 00615752339, 12634072979, 00247176060, 00117193205, 24236043102, 00121093190, 00223208301, 43063012290, 12634079067, 12634079001, 12634079050, 12783007212, 332610813, 00904792440, 33261083190, 00405538002, 33261020960, 00179184172, 00615359531, 00339590511, 00172571160, 216950467, 00339641614, 00591558204, 00781517201, 21695046760, 00725206901, 00039005150, 24236021602, 003781110, 00662411066, 00047046324, 216950967, 21695047000, 43806052440, 00049017808, 00247127003, 00378401205, 12634079082, 42571010101, 145500518, 00179141612, 00536570201, 23490563201, 00364260401, 43353037973, 00228241421, 13411035003, 33261082199, 23155023301, 000090341, 00247228890, 003781142, 17236044210, 00009372704, 00228275111, 21695047078, 35356089790, 43353026160, 00172571084, 002282751, 33261039760, 00440756514, 00378034001, 44514038536, 00603609732, 00172571010, 00781145713, 12634079052, 43063069990, 00182164510, 00069155061, 00839793916, 00115165001, 00172571170, 33358016160, 00173084413, 430630697, 00677095404, 24196054510, 00378055105, 42291030601, 00007315113, 12783007211, 43806034270, 002282753, 21695056830, 00106193204, 23490064103, 00247176202, 00615558439, 00363695510, 00677154401, 16571077550, 23629013310, 00093571115, 00364072005, 00781563506, 00009034101, 10544021730, 16571077350, 00179141602, 00781119101, 332610892, 00591090001, 00039005203, 00378111010, 00440757160, 33261089230, 00172364910, 00173315364, 00440758590, 00032471001, 00339643414, 00580111201, 00182122101, 00615155639, 00068121241, 00781530505, 12634079099, 00855203050, 00049155066, 00904612361, 00603374528, 00039022210, 23490064206, 43806052541, 00068320165, 00904023540, 00781145313, 00781119111, 216950469, 00093571193, 005913973, 42571010010, 00440757130, 00173315164, 00049017403, 167290139, 00615833439, 00480726205, 231550116, 43063043393, 10544030090, 00781145210, 00591084410, 42571010001, 12634079080, 003784013, 33261041130, 00247176000, 00536575201, 000390221, 00677154505, 00228289910, 00093834398, 00039022349, 00781145310, 00093803550, 35356093290, 216950468, 00480571101, 00677095301, 12634072942, 23629023001, 001730841, 00093726205, 43063086190, 458020822, 23155005710, 003784011, 00662412066, 23155023310, 43063043314, 000491560, 00093726101, 12634072954, 00247216030, 33261082130, 167140894, 00009017106, 10544057930, 13411022010, 00839794006, 00603376421, 35356089990, 17236044101, 00677158105, 00179151360, 11819006595, 21695056860, 45802077093, 00904507760, 00603374428, 12634079096, 009046124, 00007315266, 21695046978, 006155584, 00049162030, 35356036030, 103700745, 12634079092, 23155011501, 00378114201, 00247170430, 00143992001, 10465005201, 33261053890, 425710100, 16571077450, 00088022110, 00247228977, 12634079009, 00480726001, 00615155663, 11819008798, 16729000316, 00088022327, 00480571001, 00247176002, 00009017104, 13411034703, 00781192213, 00591046000, 145500517, 33261043330, 00093571293, 00555041705, 00179141660, 12634079059, 000490178, 430630587, 00378034201, 00615833239, 001151649, 00781563410, 00223208105, 00182199505, 42291030790, 00172571270, 00228241450, 00591558203, 16729000317, 00247229230, 10370019101, 00781114601, 006158332, 11819008791, 33261053830, 00087607411, 17236044201, 00904023580, 00603375621, 14550051802, 00603374628, 00247229090, 00047046330, 000876073, 001730842, 43353036980, 00009011402, 16729000301, 00247044460, 00182264601, 13411022110, 00179136830, 00555041502, 00143991910, 00725206810, 23490064203, 00009014102, 00182122110, 45802082278, 00603609821, 26053016001, 17236044205, 43353065653, 00143992010, 00781145705, 00378055101, 00536570305, 00093834393, 425710101, 00182164589, 16729000117, 12634079057, 24196054501, 00087607312, 15548096705, 005913971, 11819008797, |

| | |
|---|---|
| | 24236038102, 00247229077, 21695046872, 009046139, 003780431, 002282900, 12634072961, 23490563901, 38245043320, 00087607211, 00405536050, 12634072993, 00088022147, 00591558201, 00904023460, 005913972, 43063063030, 00615450931, 003780342, 353560970, 11819006560, 00093725601, 00093726001, 16364000201, 14550051904, 00087607831, 00781530605, 43063011990, 11819006591, 00615796839, 16571077401, 00009017102, 003781125, 00536564201, 00056071170, 11726015557, 35356009960, 16729013900, 00179120502, 33261089260, 12783007312, 00405538101, 00781530510, 10544058090, 00009047706, 422910317, 430630119, 001723650, 00839701406, 33261043360, 00781563431, 00039022207, 24236026619, 43063086114, 00378111005, 00117193202, 12634072963, 23490563203, 00591084530, 00179136850, 16571077501, 16590028660, 23490563202, 353560360, 00069503302, 12634072909, 216950894, 10768770002, 000090352, 00182122001, 00009344903, 00009344998, 00172297961, 42571010410, 14550051702, 15548096805, 000937457, 00894760002, 353560875, 00591397301, 00179120801, 00677095410, 00378043101, 00378112510, 00378112501, 16729000216, 005910460, 43063086160, 33358015830, 167290001, 00591559003, 00440756691, 006154509, 00781113801, 00115164902, 167290003, 00555041604, 12634072950, 00009014101, 00172571188, 00182121919, 43063043390, 00049411041, 43683012430, 00179136860, 43353037998, 33261096160, 00405537501, 00088022201, 00839793912, 33261083199, 11819008795, 00247228860, 12634072992, 00049411073, 00005306723, 00364072001, 00591046105, 35356087590, 000939433, 00603609828, 12634079094, 00068320050, 00049156073, 33261089200, 35356089730, 38245036410, 00182264789, 00143992005, 00182167901, 00781504610, 42254028130, 23490563503, 002282752, 00591046101, 00855203550, 00339590611, 167290002, 00172297865, 00093834498, 13411022103, 16364000301, 00440757190, 00615155629, 00182199501, 00068121265, 00904023561, 00093803501, 105440193, 43806052480, 11726041266, 00182264700, 43353036953, 00157069901, 000093449, 33261089290, 23490563301, 00591046010, 231550117, 35356093130, 23490563802, 00172364900, 00832049110, 38779076506, 422910316, 00088022249, 43353037980, 00173315266, 353560897, 00039005111, 16714087101, 00719197610, 35356087560, 00039005106, 42549049830, 12634079000, 24236063202, 00049411061, 24236025202, 105440219, 00904663761, 35356089630, 00904507560, 26053016201, 00223208201, 00615359663, 33261041199, 00039005215, 006157523, 00781517010, 00440757191, 23155023410, 16590028630, 13411022101, 35356093260, 00039022302, 16714089401, 00591558206, 42571010310, 00378313305, 00093803505, 17856087302, 43063063090, 433530207, 00009011404, 24196054505, 21695099372, 00069162057, 00440757194, 05165506324, 00093571215, 43353020760, 00440756530, 00115174401, 00182264600, 00364072201, 00093571083, 00781517110, 00182264605, 00555062602, 23155011610, 00615155653, 165900286, 00555041602, 003783132, 00405537601, 00686029220, 33261083030, 33261020930, 16590028782, 00093725610, 00591397201, 38245038110, 00615657639, 12634079061, 105440300, 00173315200, 00247228877, 43063039790, 00832103600, 00603609721, 19458051801, 00378313201, 12634072978, 103700191, 24236048902, 43353037909, 00302718201, 00677154405, 00039005270, 00088022347, 430630698, 001439919, 23155011510, 00904023661, 00662411041, 00087607731, 35356089930, 00440756692, 40039034001, 00093834401, 42254009030, 00005306823, 006158333, 231550234, 00247044402, 00172364960, 00781504701, 00440756590, 00615657531, 00049017001, 00904023560, 00615840705, 00009017107, 43806052441, 00247229360, 00182264501, 00228289905, 33261083090, 00228241610, 43353065980, 11726015530, 00364072101, 000938344, 21695046860, 00247229260, 00536473901, 00378043110, 23155011710, 19458055901, 00904792561, 00536564305, 00032475001, 12634072901, 12634072952, 00179141630, 00049412073, 10544022130, 00904792580, 00904023425, 00364072190, 00781530501, 11845110004, 00247229060, 00378313205, 16729014000, 44514091288, 42254007130, 12634072982, 00009035201, 00247206990, 00440756990, 00179151390, 12634079045, 12634079066, 00839701606, 24236006402, 26053016401, 13411022002, 16590028771, 353560899, 00247228960, 12634072980, 00603374521, 00121093090, 00093834301, 00603376221, 001151744, 00440757195, 425710102, 00179120560, 00781530401, 00781563531, 38245043350, 00032472501, 165900287, 332610831, 006155585, 17236044105, 00179139870, 00093834450, 23155023401, 332610830, 38245038150, 00173315100, 00555041504, 00839701612, 42291031650, 006033744, 45802077078, |

| | |
|---|---|
| | 12783007311, 05165506352, 00088022349, 126340729, 00615833339, 12280025930, 16729013917, 00088016447, 00093936405, 00157069720, 167140896, 11819006597, 00172297860, 001730843, 00228289996, 00536473805, 00182199401, 435470394, 00480725401, 12634072995, 00603375521, 216950470, 12280002600, 23490112807, 43806034370, 422540090, 00005306834, 425490498, 433530659, 00228241210, 23490563801, 33261089299, 42254009060, 000937456, 33261020990, 33261043390, 00615155631, 23629002301, 00009035202, 00677095505, 00093571219, 00405536203, 24196054605, 21695046960, 005910844, 23490563403, 00009372603, 00615254301, 42571010401, 003783131, 26053015701, 00172571070, 35356087530, 00049017402, 00781504710, 430630397, 43806034440, 00049411066, 00615359505, 00115164801, 00615558431, 23629002310, 00247229530, 008320491, 00615659639, 00603374421, 16729000217, 00615840739, 00143991801, 33261039700, 00093834410, 16714089601, 35356036060, 00069503301, 00555041505, 00115174203, 00904613760, 00480745501, 00378313105, 00686029120, 105440217, 231550235, 12634072918, 00591397101, 23155023510, 001210930, 00068121050, 26053015801, 00009007002, 001439918, 00440758990, 00781530405, 00832096800, 000491620, 00068121210, 00781145501, 145500519, 00440756560, 00364072105, 00093745701, 231550233, 00405537603, 00603376432, 00725206801, 002282899, 000491550, 00228241410, 10768747501, 00172298060, 38245043355, 00480571205, 003783133, 000937256, 12634072990, 35356093230, 00009013101, 00781530601, 00904792480, 422540071, 00088016347, 00093425652, 43063086198, 00662412073, 11819006598, 13411022001, 00615254313, 000937254, 00039005110, 00302718001, 00781119110, 00440757192, 00839804016, 00009035203, 001151650, 00904612461, 00009014199, 00039022301, 167140873, 24236026602, 42571010501, 00093947753, 00009014103, 003784012, 43063069993, 00179120530, 00093725410, 00405536201, 00591084510, 43806052580, 105440232, 00781563510, 001730845, 00039022201, 004406564, 00088022210, 00069503101, 43353020745, 00093571093, 16590028760, 00440756892, 23155023505, 21695089400, 43353065692, 00093745601, 17856289802, 43063069793, 003780551, 00009035214, 000390051, 12634079018, 43353037992, 00480571201, 42571010510, 00405502601, 00039005177, 43806003640, 00686055101, 33261083130, 16714089602, 33261039730, 000938342, 00049155073, 00781193210, 00339643314, 15548096701, 42571010110, 10370019001, 00247176330, 00223208202, 10370019005, 00093425601, 00615840839, 00615359553, 43806052540, 00247206960, 00102313401, 00069162030, 00302718401, 00087607311, 00339641511, 43063069890, 00839793906, 00378401305, 00364072002, 43063069830, 00093571019, 00364257805, 000390053, 000937455, 216950747, 00247215930, 00339643411, 005910461, 332610961, 00349841701, 10544021930, 00087607212, 353560896, 00894760003, 00182199400, 00179120536, 33358016130, 00832049211, 00039005305, 00115164802, 00603375628, 00725207005, 16571077301, 00179136870, 16590028690, 00591046005, 00069411066, 00781145701, 23155005801, 00378021710, 33358016060, 13411022106, 33261083160, 00007315313, 007815635, 33261041160, 00172297970, 00580111305, 00247176077, 00047046430, 43063094693, 00172298065, 00781563508, 00603609621, 33261041100, 00068121261, 42291031710, 00536473902, 43806034380, 35356097030, 007811453, 00364257701, 00781193201, 00121093091, 00247127000, 10370074601, 43806003441, 003781105, 165710775, 00115164901, 00591084515, 422540281, 11819006590, 12634072985, 00223208305, 26053015901, 00378021701, 000935711, 00247229160, 00102313601, 00179184170, 00364260405, 00615558563, 24236079902, 00093834350, 24236027602, 43806034470, 00781113810, 45802082293, 000390222, 10370074501, 12634049060, 23490064209, 430630630, 42291030501, 43353065680, 23490046100, 433530656, 42549049930, 00536473810, 00247044430, 00069155073, 00247229330, 10370019105, 21695074790, 00228241650, 00039005115, 12634078480, 00591084430, 00855202950, 43806052470, 00839804106, 001723649, 43063012190, 103700190, 24196054601, 000090131, 00093725401, 00247206930, 33261081390, 006156575, 000090171, 00247176302, 13411022003, 00440756660, 13411034603, 00093834319, 33261083560, 12634072945, 13411034803, 006153596, 00378313301, 105440220, 12634072991, 00039005208, 38245036455, 00056071470, 43353058260, 00009011407, 38779076503, 00093571001, 43063043301, 00228289901, 23490563902, 00719197113, 433530369, 00603609723, 00093725652, 00339635814, 43063039786, 43063012130, 43063043360, 11822518601, 00615840730, 43353065690, 009046138, 00591084401, 42254009090, 00839804116, 00781504601, 00781530610, |

| | |
|---|---|
| | 00781194201, 00182167704, 00047046424, 00093936410, 21695047030, 43806034480, 165710774, 00364260505, 00228275196, 00093834305, 12634072969, 23629002401, 00093834419, 33261096130, 00480726201, 42291043010, 23155023501, 00088022301, 00115174403, 00182264689, 00247228930, 43063003430, 00093745501, 16714087301, 00677158101, 00007315265, 00007314913, 00405538001, 00364257801, 00591046100, 00172364970, 00781145613, 10768715002, 00591559004, 00173315300, 00247044403, 00480726101, 24196054610, 00182199405, 00904792460, 21695046830, 00615359653, 00904792560, 19458050601, 43806003580, 008320492, 24236036802, 00405502503, 00378401201, 00781517205, 00009011405, 00039022310, 00904023525, 33261043300, 43063069893, 425490499, 00115165002, 33358016101, 00440756690, 11845110001, 332610397, 43353036994, 12634079060, 00069503202, 00781193213, 007811452, 00615359539, 00069087130, 00536570301, 00182164504, 42571010210, 422910429, 12634072960, 35356036090, 16729000201, 12634079040, 00009017110, 00007315364, 38245036420, 000935712, 43806034240, 000490170, 000939364, 00536564301, 00179151370, 12634072966, 00677154501, 12634072900, 00247127060, 003780217, 12634079012, 50090709002, 10768747502, 33261082160, 000876074, 000494110, 00182264701, 00378401105, 13411022102, 43353065960, 105440580, 00725206805, 00247215900, 009046637, 00904613860, 23490744901, 33358015860, 35356089760, 00172571080, 332610835, 00172297960, 21695046878, 12634072996, 00143991901, 00247229030, 00228289803, 43063086130, 00182199489, 35356099590, 433530895, 001210931, 00093936401, 00179120590, 43353036960, 00364260501, 231550056, 00093725661, 00378114210, 00615796805, 38779076504, 00115174201, 12634079097, 00247228830, 00087608131, 00172571180, 00247127090, 00039022249, 00591090010, 38245036450, 00781145601, 00093571205, 00093571005, 00339643311, 00555041605, 12634072967, 00781145710, 00839675516, 42571010005, 458020947, 00536570205, 33261039799, 00904613960, 12634079079, 12634072981, 00591084415, 000938034, 00093834310, 12634079084, 00555041702, 00247229430, 11845110103, 23155023405, 009046137, 006033746, 00179136890, 00725206905, 21695047090, 00009014106, 00440656501, 00049412041, 43063043330, 33261083060, 23490064106, 10370074605, 00056071485, 00093571101, 00904613980, 12634079063, 00121093192, 00349841810, 33358015760, 00223208302, 35356097090, 435470396, 00480745701, 00069412066, 00480725601, 42571010505, 00247229130, 00228290096, 00440757114, 33261041190, 00440756630, 00440756992, 00009034104, 14550051704, 00378110501, 43547039610, 35356089960, 00839804006, 00440856390, 00093834405, 43806003440, 43806034327, 00615254402, 00093725461, 00405502401, 00068121141, 00480571083, 00172571110, 10544058030, 00228289950, 105440221, 12634072912, 00781145213, 35356097060, 00615558531, 00405538102, 00405537401, 16714089501, 00172571060, 00172297980, 00364072090, 00480725501, 00480571105, 21695074630, 00480571115, 12634072940, 10544057990, 00832049111, 00179184171, 00591559104, 435470395, 006157969, 430630861, 00349841905, 231550115, 12634079090, 00007315213, 00440756890, 21695074690, 00228275311, 00049017807, 35356093160, 000390052, 11819008790, 430630122, 00904023640, 42708008330, 00580111401, 00179151350, 00378401301, 00093803401, 00480726105, 00440656401, 11845110104, 00009372706, 35356011300, 43063086193, 00591084501, 16729000116, 000939477, 16714087202, 16714087201, 35356099530, 007815634, 00725207010, 00591559101, 15548096801, 43063058730, 00173315213, 16364000101, 12634079091, 00009011406, 00039005206, 12634072971, 00536474401, 00832049210, 00405536102, 332610433, 00039022202, 427080109, 43353065660, 422910306, 00247044400, 425710105, 12634079095, 242360266, 00781517210, 23629024001, 10768770001, 45802094793, 00009017113, 00603376321, 42254028190, 45802094778, 00039005210, 42571010205, 002282898, 00228290050, 00173314913, 33261082190, 00088016247, 00349841801, 21695046730, 12634079074, 00536569701, 000935710, 11845110101, 00182122005, 23490064200, 43806003541, 00093726105, 00173314813, 12280001200, 000090141, 00121092990, 00536575205, 00157068301, 006153595, 00039022111, 00591559103, 43806003540, 422910430, 16729014017, 10544022030, 13411034903, 00719197109, 00009017108, 00069155030, 00173315313, 43806034340, 00904792461, 00039022211, 00832096713, 35356089690, 00555062502, 00172365000, 00179020588, 00839803906, 00039022311, 00555062702, 00143991905, 19458033600, 00115174303, 43547039550, 00781563406, 00339641611, |

| | |
|---|---|
| | 00093834201, 00068121165, 00781114610, 00179120524, 16714089502, 12634072957, 00781517001, 23155023305, 167140895, 00093943305, 006156576, 430630433, 21695074730, 430630120, 43353065694, 23155005810, 006156596, 004406566, 33261081360, 00662411073, 00228290010, 43063069790, 44514091290, 23629002210, 00662412041, 00093571105, 00781530410, 43547039650, 00172571288, 17236044110, 12634072999, 33358015800, 12634079093, 006157968, 00009014107, 43063058790, 00247176030, 00677095401, 00781192201, 00179141601, 42571010201, 00904507660, 33261081330, 00009047797, 00009035204, 353560931, 21695096730, 00440656601, 00172571280, 16729013916, 126340790, 33261039790, 16729000101, 00904613840, 16590094060, 43353036992, 43547039510, 26053016101, 43683012360, 427080083, 00093803510, 001210929, 00228275296, 00440758995, 14550051902, 00228275250, 21695046930, 430630034, 43806034335, 105440192, 12634072997, 11819008760, 00591046110, 16590028730, 00839794012, 33261083590, 231550058, 43547039410, 422910305, 43353065670, 000938035, 00173084213, 00536473905, 16252051901, 006158334, 00536473801, 16252052001, 12634072984, 00781517005, 42571010305, 00009017197, 004406565, 00339635811, 16252051801, 00009017103, 00832096700, 00049412066, 23490046000, 00009017105, 00068121201, 42291030890, 00173315265, 12634072998, 00007315164, 00173084313, 003781113, 42708010930, 38245072510, 00591090030, 00049156066, 00228290011, 00904023660, 006033745, 14550051804, 24236025502, 00894625502, 00228241496, 00228275396, 000494120, 00615359563, 00093803601, 00009372501, 00378111301, 00536473802, 353560995, 00009017194, 00179120701, 00173084113, 00480571215, 00781504501, 00781563408, 00247127030, 00182167701, 103700746, 006151556, 42291030990, 00069503201, 00093726201, 216950746, 00182122010, 16590028790, 00179120512, 00009011474, 00172298070, 13411022006, 001151743, 00904507780, 10544021960, 43806052570, 00904507740, 00580111301, 00179178971, 00615657539, 00894760001, 003780340, 23155005601, 006158408, 00228275211, 430630699, 65243018318, 58016037697, 498840745, 52125076123, 58016054483, 614420117, 57664072418, 67544051194, 65862057901, 51129203401, 63874093910, 538080942, 680714857, 58016069105, 63629339703, 55289060630, 61392070960, 52125076419, 59115001401, 51129407702, 63629101601, 65862002905, 66336073060, 63187023490, 58016038305, 68071323906, 63739011915, 58016044792, 63874043224, 552890779, 55154558800, 49349009202, 63629139806, 51129284401, 607600176, 500903069, 48695120902, 52125076719, 58016044775, 68071445308, 636292907, 59762503202, 61392059425, 66105098411, 55111032101, 64764030447, 68071314109, 53978201402, 60491064434, 66336071290, 58016037802, 55370014707, 61786028623, 58016037627, 64725013901, 636296930, 58016033407, 49999051430, 51138037230, 53978300009, 55700058590, 58016041100, 67544061380, 58016046700, 52125048120, 58016038669, 55370086307, 50268035615, 500905776, 58016055530, 54569855603, 521250761, 50090317201, 53746028610, 54868514801, 61392076625, 61392083439, 636298181, 50090050304, 55953003570, 58016038350, 58016069102, 52544056001, 51285032504, 65427037706, 49999078160, 58016054326, 51285059902, 68071445303, 67544065353, 53489015204, 50436087403, 50090317200, 68071309703, 61392059430, 551545588, 68071413308, 67046023707, 60760022990, 658620890, 493490170, 60505014201, 58016038650, 51129469201, 58016054308, 67544012980, 55045213802, 55370014708, 663360269, 51138025230, 63739011710, 60505014102, 67651032005, 53217025090, 636292793, 59762702009, 58016044714, 67544009732, 55887053690, 572370151, 54569383100, 68084078895, 60429091901, 63187065990, 58016054325, 500904911, 66336085090, 58016033473, 50090207600, 65243018518, 50090429700, 680713097, 597625032, 58016038367, 66116043230, 51079081101, 61919072230, 67651004803, 65243028509, 51129324702, 58016033497, 58118046106, 52959088830, 54569383002, 67253046110, 68071413302, 58016038687, 55154053406, 58016037610, 67046023615, 55045261700, 55111032279, 64764030233, 65841082416, 51079042520, 67544092780, 64720016750, 55370046111, 54868420603, 54868521001, 55953003580, 51129349502, 647200291, 53217032390, 61392006391, 68071422309, 58118032206, 65862088830, 58016037626, 65841082501, 617860831, 58016038604, 54868326506, 57315001003, 68071410909, 65243028506, 65427034101, 58016033490, 51129355601, 680711910, 61392006430, 65841082610, 50268036015, 54868484200, 61392006439, 58016054476, 68084032701, 552890125, 67651002300, 55887065730, 619190250, 60491041230, 59762054001, 63629641301, 68071437806, 54868420500, 58016054356, |

| | |
|---|---|
| | 59762503101, 59762702205, 52959082320, 58016054306, 54697007901, 58016054428, 66105098502, 58118834408, 55111032130, 52125097202, 68071449808, 65084018010, 54441006901, 670460240, 58118145206, 58016087628, 636296840, 58016054415, 55887069330, 54868326502, 58118233108, 61919028760, 52817012110, 55953034327, 55289017330, 63629101505, 52817012100, 54569855600, 49884012406, 66105075610, 68115016030, 493490537, 57480039201, 64720029110, 51138026830, 49999011390, 63874043281, 58016038332, 58016069180, 61392072495, 63629220401, 57237002101, 64720016710, 62540725603, 58864095230, 52125047902, 66105075602, 65427035204, 55289060690, 551545476, 63187023430, 55111069601, 500900495, 58864022460, 45963075304, 46193069710, 551542677, 50090577602, 64764030230, 51432046303, 49884084701, 54868441200, 50090207501, 54868333503, 63629290702, 66336066260, 60505014100, 68071486909, 57237002001, 55887036830, 52544055910, 658410661, 68071414203, 66116080560, 63629139204, 54868037302, 68071240209, 58016038679, 68115015630, 60491028390, 55045226606, 548683327, 65862008218, 63874043221, 66267010260, 55154053908, 55700025630, 52959017730, 51129309802, 59930163903, 55048024330, 552890976, 63629139802, 680712589, 58016033401, 68071401008, 67046023721, 54569220601, 54868521000, 55111032098, 58016038644, 50090162602, 65862089030, 596510270, 58016033498, 59762378303, 68115015860, 58864068930, 61392059325, 54868371100, 63874031624, 63629525601, 54569561803, 68071191008, 50090049508, 58016087610, 50090049602, 46193069705, 63187069860, 54868099704, 65841083210, 68071401002, 68115015490, 516550103, 51079029320, 64720014211, 58016038696, 63539016257, 66267023060, 65243037509, 61392006490, 557000594, 65862088901, 50090291500, 51129257200, 66105098509, 54868108901, 58016044707, 59930162202, 58016038625, 47202250903, 60491041401, 65862008201, 52125076119, 50090152601, 658410659, 52728022710, 67544041770, 67544075680, 58016037870, 68071218903, 58016069106, 636296309, 65427003201, 55370046011, 62540725604, 58864071130, 500904199, 551110696, 54569393700, 57237002501, 528170121, 65862008030, 54868099602, 64909010508, 59762233208, 65862088905, 68071402006, 51129014301, 59762503301, 54868342600, 58016054450, 51079042620, 54569469500, 50090050301, 58016037696, 54868420501, 61392011732, 54868332701, 58016054373, 67544056631, 557000624, 67544030280, 55111069505, 54868331907, 61392059525, 65243037809, 62584012000, 63629724901, 521250971, 51138033730, 58118233103, 516550381, 63629525602, 54569369001, 61919028782, 55045330001, 58016033471, 548683266, 58016037840, 54569383000, 500901080, 68071218906, 66336002830, 551545070, 605050142, 59651026901, 53978022609, 680712181, 61392011731, 572370153, 680711991, 65862008005, 54569384102, 58056012210, 68071191006, 53746028605, 49999010700, 502680357, 59762378101, 680711967, 55289089297, 53261095430, 54569856500, 58016044772, 68071300902, 58016044728, 57664039888, 58016054484, 50090050202, 500902492, 59762503302, 63629642103, 66993016202, 548685243, 55289042497, 504360024, 67544009770, 58016054332, 61392006425, 53002044660, 65841066010, 52189029124, 58016044797, 521250728, 55953003440, 61786046719, 58056012310, 58016038671, 61392072439, 581188344, 55953034370, 55700059490, 61392059631, 680840327, 65427003101, 504360141, 58016038621, 58016069156, 658410660, 55154798700, 58016038302, 66105075402, 55370086407, 52959088800, 58016037882, 58016054471, 54868452900, 66267023130, 54868099703, 54569383102, 61392083460, 66105098450, 58016044708, 55154120107, 54569383109, 62269029324, 51129377401, 51813022830, 49884096805, 55953034440, 67046023720, 51138047130, 63629139407, 60491064430, 53002211103, 631870641, 58016054405, 66267010010, 51079087317, 58016044756, 61392059331, 58016054481, 66267031630, 58016054360, 61392011751, 54569585500, 61392011760, 52125048123, 58016005803, 45865057430, 63629407103, 53002446003, 58016000501, 63629139202, 680714643, 538080409, 58016038672, 64720016810, 58016054408, 68071009430, 57866646202, 50090207506, 58016038312, 61919025082, 50111058501, 500900852, 58016044784, 58016069148, 58016044715, 58016038370, 63539015530, 65841083305, 66336093860, 49349027202, 619190720, 62540725404, 51129301401, 500904215, 50090085200, 63874031750, 51079087219, 51655038152, 53002041750, 680714939, 58016037897, 59012041157, 67544092760, 55953034335, 636296476, 63874066514, 58016069135, 58864021430, 65427015573, 58016054384, 52125039320, 66336026930, 611450203, 605050141, |

| | |
|---|---|
| | 647200292, 58016038636, 680711836, 64720014010, 58016038677, 548685148, 49884096705, 662670102, 54868498803, 66336057390, 54868326503, 58016044727, 50090291502, 68071074190, 57664039988, 58016038632, 49349022802, 54868460901, 58016038660, 63629220301, 47679056262, 50090419902, 55154215000, 58016033445, 538080259, 64980028101, 58016005802, 54868124503, 54868498801, 58016037669, 58016044732, 58118032208, 68115016020, 54569383001, 58016069177, 58016000505, 66105098511, 552890301, 61145020250, 51129381701, 55700061290, 530024170, 65862002919, 58016033467, 67253046111, 58016054342, 50090400800, 55045226601, 49884045105, 49349071902, 59930159201, 572370022, 59762372703, 65427035202, 50436087301, 67046024060, 551110695, 62540725601, 55887053501, 521250764, 65841083205, 50268036211, 51138047030, 58016037648, 64720016811, 63629139305, 65862002901, 55700062460, 68071240206, 58118032100, 680713141, 502680360, 55370059608, 680010177, 67253046211, 53506053030, 57866640906, 65862003005, 63739011703, 548685210, 500900502, 61392012051, 58016000500, 49999080800, 55154053400, 64764030430, 55289028997, 58016037814, 58016069183, 52959082360, 66336057360, 57237002205, 51129176101, 67544064392, 67651002401, 58016037893, 51129021500, 68071194203, 58016054448, 54569384204, 50090050501, 53808094202, 59012041166, 51432046306, 54868536401, 58016033427, 53002211106, 55370046210, 66267009991, 61392059530, 58016037645, 59115001205, 63629818101, 61786073023, 58016038602, 65862002850, 61392006345, 55887072701, 59012041241, 675440302, 52446047721, 63874058810, 51129351001, 61392070939, 53808036901, 63874066510, 54569220602, 61392083465, 55887021260, 680714994, 55370046250, 680714232, 63629125503, 63629559801, 49999010800, 60760017590, 552890892, 66336066294, 58016000502, 54569585501, 63629724902, 680714904, 58016054436, 57237015201, 58016038640, 58016069150, 680712228, 54569020603, 604290919, 51129241801, 63187069830, 57866646101, 62269029229, 58016033418, 49349036002, 65862058005, 548684609, 64720029211, 54907042501, 50090088701, 57315001001, 670460235, 63629315801, 54868420601, 60760014190, 61392072425, 58016038656, 67544066141, 61392012031, 675440129, 58864095730, 64720012406, 49999010890, 680714133, 58016005805, 52959082260, 51138026530, 68115015830, 54458096716, 68071402003, 58016084400, 60491028334, 607600329, 672961247, 63874043204, 572370152, 51138037210, 67544092732, 61392011939, 50090088702, 63304042510, 54868331901, 61442011501, 581182332, 50090307400, 54868124403, 55111069701, 55700057690, 55289097660, 61442011605, 50090203004, 58016037836, 51813026060, 65862002830, 55887053560, 57315001101, 67544075153, 65841083277, 67544061360, 51129241701, 57664039918, 58016037873, 499990108, 51129176601, 48695120202, 66267023115, 49999080790, 54868484201, 54348010430, 68115015590, 51129145501, 67544056660, 658410834, 636291392, 51927311100, 52125076519, 51079081119, 636295650, 58016038384, 54868326500, 66267010092, 67253046150, 65862002999, 538080369, 58016054418, 58016037896, 68071493901, 60429019401, 58016069107, 52152013402, 68084011201, 66336053560, 50090100401, 58016054480, 58016037602, 57480039106, 63629279302, 58016038376, 65841082405, 55370059407, 67046023660, 60760002460, 52189029229, 58016038328, 58016069142, 607600746, 68071469503, 58016069170, 66336073090, 58016044725, 636297248, 658410826, 68071194408, 50268035711, 47837005800, 51285032730, 58016054460, 54569599200, 58016037620, 60429008260, 63739011603, 62528034435, 50090537702, 63739011910, 61392070931, 57866707302, 55154507000, 54868342601, 66336085060, 58016054402, 65841066177, 61392006431, 51138033930, 49884074605, 60429008527, 636291393, 60760017690, 52555029201, 55370059207, 63874043250, 61919028690, 65841083301, 50090108000, 60491027801, 55154623809, 61392059225, 66105075409, 51129377402, 50090100403, 65841083310, 61392083490, 66267009960, 50090049402, 54569384103, 68084080701, 49884012306, 55289042430, 50090491102, 649800279, 55045226501, 60760022930, 58016033416, 63629279307, 58016054487, 66116082860, 51285032602, 58016069199, 597620540, 58016037614, 54569384202, 65841083206, 50268036011, 58016054489, 61392011730, 65862058125, 62939322101, 58864002730, 57664072788, 53808025701, 54569384205, 68071437802, 47679056101, 58016054497, 55154522407, 636295256, 55887017930, 61919071530, 57664072588, 67544056653, 58016038314, 58016038340, 54868331805, 58016038607, 572370020, 50436087303, 58016054392, 58016054440, 50090085202, 58016037687, |

| | |
|---|---|
| | 50090504000, 647200141, 55111032197, 63874031628, 68071314108, 63629139801, 54868108903, 58118233100, 58016033432, 63629641003, 63874058804, 647640304, 66105098406, 52544055801, 58016000530, 459630753, 57664072513, 50436640901, 61392059231, 61392072465, 636293158, 57237002401, 51129301402, 58016038690, 55887053660, 68084011101, 49884045201, 58016038325, 68084074525, 597627022, 55887033960, 66336078460, 49884012206, 58016037603, 53217028560, 52125038920, 63629630902, 54868326600, 548685185, 51285059905, 51813006499, 458650710, 67544030232, 49999066060, 65841083377, 60505014202, 54868545702, 62584035200, 500902076, 67544030245, 58016038308, 51079081019, 53808040101, 62037087130, 606870480, 67544012960, 61392006451, 61392076630, 52125038923, 58016054426, 58016038360, 49999011360, 65841082577, 50090085302, 67544061370, 55370050607, 54868124505, 61919072330, 572370025, 55700025660, 53002111106, 680840326, 63187081190, 63739011810, 54868333402, 53217031760, 55700072230, 59115001305, 58016087612, 66336053530, 63629139404, 596510269, 63629139805, 59762378203, 67544087532, 672530460, 54868333500, 60505014200, 604290082, 68084013801, 64720014250, 63874043202, 61919025090, 54868441201, 54868332700, 58016037636, 498840746, 581180461, 68071423201, 54868333404, 58016044744, 55887069301, 50436707403, 63304042701, 65841066016, 68084074595, 58016054328, 61392011745, 60491027860, 68071038860, 58016033409, 619190476, 67544075160, 62584011050, 67544051170, 50090421502, 58016038684, 58016044750, 61392012039, 58016037810, 54868441202, 49349053728, 54868371101, 54569020601, 53808040001, 502680356, 61919071330, 52125039223, 544580966, 51129355602, 61392059560, 58016054345, 500901082, 55289028130, 65427041266, 61392012091, 61786056223, 68071302406, 63874031704, 67544029670, 63874058820, 49999080700, 54868528802, 58016069127, 54868108902, 64980028105, 58118046108, 53261017130, 619190713, 52817012200, 59651027005, 58016044799, 51432046503, 55048024560, 61392011739, 617860245, 60429008310, 55048024230, 60491028430, 61442011601, 68084013711, 52152013505, 58016054379, 58016069125, 53002211100, 53978031509, 51138026730, 51129014401, 57866640902, 55289018797, 61392006351, 54569383105, 66105098410, 63629641001, 49999010760, 49999066030, 576640398, 597620542, 54868301700, 54868168801, 60505014204, 59651027030, 61786022602, 51129136601, 58016037660, 52953009801, 64909010409, 63629806201, 58016046760, 636292203, 53002044690, 54868099601, 68071412402, 55045213801, 61786024502, 58016038673, 50268035811, 54868409100, 58016038356, 63874031790, 528170122, 60491041334, 66105098506, 50436087201, 54569554702, 63874058890, 55887022260, 55289028186, 58016054416, 51655081025, 54458096710, 68071030730, 538080678, 58016038396, 53489015205, 65841083477, 50090050402, 67046024030, 51129455502, 55700002260, 59762702105, 63874043290, 68071194206, 58118032103, 53808040901, 63629565005, 58016054493, 530022111, 57866646203, 55887069360, 636298062, 59762375706, 52125057302, 53489015301, 658620889, 500903172, 63739011615, 55289097693, 63629139303, 53978201305, 59930159202, 516550106, 57664039808, 65841066077, 493490360, 53746028701, 67544065394, 66267023120, 58016054340, 59762054102, 58016069120, 66336078430, 619190725, 55370050608, 680712385, 54458097310, 63629315803, 61786074519, 63629693001, 55154120101, 58016054375, 636291015, 529590822, 636296410, 680715215, 516550120, 63874031614, 51129196301, 58056027610, 58016069197, 55953003540, 58016033499, 58016038682, 58016069169, 54868521003, 55111032278, 63629139803, 51655010552, 67544066181, 607600229, 60760056890, 67544009760, 55045360702, 57237015105, 61392012032, 46193069901, 65841083230, 58016038609, 52959088860, 53489015210, 60429008201, 68071502900, 63187081130, 59651027001, 55154522400, 49349027219, 66105098409, 67544064394, 64720016411, 68071238509, 58016054327, 67544069031, 52544046001, 66116082760, 50090050300, 66993082130, 51079081017, 61392011932, 54868331903, 510790425, 617860286, 58016033444, 51655081052, 58016037845, 58016038316, 54569561801, 61786002319, 680840111, 65841082401, 63629304305, 55887022290, 66116080660, 54569860102, 61392083425, 54868420502, 51129018500, 538080408, 58016054435, 58864016160, 60760014198, 680714718, 500903189, 68115016090, 53489015201, 57866641001, 54868333502, 50090050500, 50436646304, 52125048102, 51129289601, 65243032518, 58016033415, 58118233109, 58016054303, 56126010311, 59115001255, 67046023620, 500903074, 66267023110, 54868331800, 67544056692, |

| | |
|---|---|
| | 680714093, 54868452902, 50090491100, 60491027980, 63539015561, 680840112, 60760023090, 61392059230, 68071038830, 50090248600, 51285032605, 58118233203, 53489015303, 47837005710, 55887053530, 60760074530, 58016069136, 59012015557, 55887072790, 64764030202, 58016069160, 61786073019, 58016054490, 68071449803, 500903547, 50090207505, 58016044748, 54569220603, 51129272801, 59651026830, 63629139405, 63874031612, 680840556, 62269029429, 551542678, 65841082605, 63629290703, 51655075324, 68084013601, 68071410908, 61786032619, 54569445301, 50090248604, 51285032704, 504366408, 68071413309, 502680359, 58016044735, 65841082506, 68115016010, 52959044901, 55154390901, 55154619000, 68071445203, 53746028602, 68115015930, 647640302, 532170317, 49349009230, 500900887, 54868326601, 65243017618, 57664072713, 680840807, 58016054469, 55289026590, 631870698, 65841066110, 58016087615, 58016069103, 61442011705, 60491028401, 663360028, 61919028660, 54569383104, 50090222002, 52446047521, 50090049501, 50090580600, 58016041102, 63874043260, 55160012305, 58016038383, 58118834309, 62528034440, 60491028234, 45865057460, 48817770101, 58016044730, 55887036896, 58118834409, 50090050204, 61392011991, 47679056204, 63629290701, 64909010107, 61145020350, 58016037884, 617860373, 58016033448, 57315001002, 47837005701, 50436707501, 51138026630, 61442011610, 58016033403, 51285059802, 58016037607, 50090203002, 54868518802, 66336093830, 55370014607, 58016054301, 551548270, 67544012953, 50090354700, 50090108201, 52152013304, 65243018536, 63629630901, 57664039908, 50090342901, 50090050502, 57237002301, 57866630301, 51129354601, 54569393800, 64720012503, 65862058105, 504360873, 58016033472, 58016037691, 68071196708, 63187065960, 54569384104, 59930162201, 63874031621, 64909010108, 55045226708, 63629139306, 65841083430, 61919028730, 680714378, 58016038303, 58016005860, 617860326, 51655036126, 58016033493, 51129381802, 50090203003, 55048026930, 636291398, 50090050506, 68071521500, 521250105, 55045232208, 58016037820, 55289061401, 55154267803, 58016054400, 53002417000, 57866646001, 504360874, 68071314102, 54868337702, 58016038389, 63629139203, 636291394, 680714124, 47679056235, 58016069132, 63874058814, 52125076402, 58016038348, 510790873, 51079087201, 58016038306, 54868518502, 55700072530, 58016037693, 51129365201, 51079087319, 66267022930, 63187048630, 57866641002, 53002446203, 55111032201, 68071402008, 63629304301, 54569561903, 63187048690, 68071469509, 46193069902, 58016037880, 64720016510, 63629399801, 50268036215, 544580968, 66993016247, 68071016530, 50090537700, 54569384101, 680714695, 58016037826, 58016037692, 516550361, 54868518800, 58016038618, 51138037130, 64909010507, 49999078100, 63629304304, 57664072488, 58016054310, 54441006925, 65841066030, 55887084530, 67296089401, 607600228, 63874043220, 55045226609, 66267010205, 57237002050, 58016084490, 52125038819, 55289028190, 58016054330, 680713009, 58016037898, 65841066101, 54441006905, 57866646301, 51129140501, 51138036830, 51608011402, 63629279305, 63187066730, 61919025030, 62939323101, 61392011790, 55953003541, 636293998, 658620579, 63874093930, 51138036930, 62269029224, 63874031760, 68071485701, 53978201404, 672961246, 61392006432, 58016037804, 53489015305, 63874031730, 58118145308, 55370014609, 67544012930, 68071449806, 58016054412, 680010179, 500905806, 58016038301, 51129289602, 63874031650, 672530461, 67263019301, 548683335, 58016069124, 68071191003, 65427037501, 54868545700, 55700039590, 680712189, 66105098550, 59762233108, 50111058502, 54868331801, 68071414206, 658410832, 61392011754, 538080401, 55700058530, 60491041330, 54868333400, 65862002930, 53217025099, 58016038627, 58016054369, 58016038381, 58016033404, 54697007902, 58016037615, 68071445206, 54569383101, 52125047920, 53808026001, 54458096610, 58016037600, 63739011610, 61392083495, 58016037667, 53002111100, 65841082630, 54868099603, 636296421, 52817012210, 51129186801, 57664039913, 55045232201, 55953003441, 58118145203, 51655010352, 55887053590, 532170349, 65243017636, 58016054382, 50090580601, 65841083216, 548683377, 67544064360, 680711557, 65427035201, 68071306806, 58016069198, 54868528801, 647250058, 58016037682, 49349033802, 58016044742, 54868331804, 55887053601, 52959059890, 61392059539, 61392070995, 55289097601, 64720016711, 54697007905, 55154599200, 58118046109, 61392059460, 68071082330, 58016069116, 60760017530, 63629565003, 53217025030, 66267010030, 55154566200, 68788834303, 50111058402, 680715029, 607600165, |

| | |
|---|---|
| | 52189029129, 60491064534, 54868333403, 63874043228, 54569855602, 51285032604, 50090100402, 63629630904, 58016037891, 63187066790, 55048024530, 50436646201, 61786022619, 54569384206, 551110322, 675440511, 57866630201, 63629641302, 50090110400, 58016087660, 607600568, 60951071185, 530024462, 51655028924, 53746028624, 58016037860, 66336078490, 46193069801, 591150012, 58016037800, 68071402009, 617860023, 59115001405, 58016054475, 50090248504, 61392059625, 64720029011, 57866641102, 604290918, 68071002830, 54569020050, 54868524304, 59930163901, 51079029120, 55045226602, 68001017903, 58016038372, 54569554802, 662670099, 57866646201, 581180321, 49999040130, 52959044930, 50090198700, 60687048021, 54569384200, 597620541, 64764030402, 63739011701, 54569860100, 53746028705, 63874031610, 68071191009, 50090088803, 66336093890, 53978069402, 58016037883, 55154455709, 54458096810, 58016054393, 58016037828, 67544064380, 58016038392, 55289097630, 58016054390, 63629139205, 50090085300, 68071194209, 67046023630, 53002417003, 58016038698, 67046023628, 647200140, 67544030260, 68071490409, 58016069172, 68071309706, 54868518503, 55289080630, 54569561900, 61919072590, 51813010660, 47837005705, 64720014050, 53002041700, 59115001201, 504366302, 50090342909, 51079081120, 62584012050, 68071002860, 54868490600, 58016054387, 63304042601, 51129309801, 52125038820, 680714142, 58016038326, 55953003640, 59012041266, 557000469, 55289080690, 55700062490, 67651002301, 58118834300, 58016037881, 51285032505, 54569383200, 51129191401, 65862003075, 51079081001, 66336002890, 65243028503, 68071314103, 60429008536, 54868331906, 55370014709, 68071183601, 548680795, 59762233206, 58016038382, 58016033410, 58056012366, 51655033726, 68071437809, 55154599204, 61392006331, 57237002505, 58016033424, 500902485, 63187084890, 67046023714, 51129253601, 500902486, 50436707401, 50090088700, 51079029260, 68084078825, 66105075603, 58016033420, 672961267, 54569020000, 61392012045, 636292205, 54907042601, 67544075645, 67544061392, 68071409306, 54569585503, 680714453, 61786083119, 68071306803, 51655094226, 58016069182, 50090207604, 58016054391, 66993016347, 661160360, 57866707402, 581180065, 63187064130, 55111032001, 50090207504, 50436014101, 58016038648, 49884084710, 67544047882, 65841066130, 61442011505, 67253046011, 57480039206, 52152013405, 51432046103, 57237015205, 50090203101, 55289012530, 67046023715, 61392059431, 50090219700, 636293043, 50268035715, 51129284402, 55154053404, 62528034427, 65243018527, 63629565001, 55953034340, 63187047730, 61442011725, 66105098610, 58016037806, 68071196709, 49999040190, 58016037670, 47679056301, 50090248503, 62584036150, 58016037675, 58016084460, 58016069130, 51129148701, 54868409102, 57237002201, 58056029701, 51129351002, 55953034240, 54868331904, 55289097690, 68071409302, 52152013404, 57866640802, 50090301201, 58016054371, 63874031660, 58016033440, 68071413303, 54569855601, 50090049609, 58016033489, 51129148601, 50090100502, 58016033406, 52125038919, 65841065977, 68071323909, 57237015301, 54868524303, 53261095460, 58016054407, 521250972, 58864068960, 58016069184, 551110697, 60760002430, 58016000560, 58016038318, 62584017100, 50090162601, 66267010090, 67544064332, 53002044600, 59762372706, 51138037030, 60491041460, 54868518501, 67544087560, 62318014101, 58016034430, 64720029010, 619190850, 58016037681, 58016033479, 538080625, 64720016611, 50090050305, 63187099330, 57237015101, 63874031605, 65862057999, 60429029301, 58016038380, 68071521502, 66993016302, 51316010102, 54458096616, 680714498, 597627020, 680714109, 58118145208, 51079042601, 63874066530, 50090049500, 68001017703, 58016037672, 65243037812, 63629139302, 66116042760, 631870477, 58016054406, 50090318900, 672960894, 57866640904, 68071414209, 636292202, 576640399, 57237002199, 60491064630, 55887036860, 60687048011, 54569599201, 680840295, 54868331803, 64980027903, 52125047919, 60491028230, 59762233005, 59762054202, 61392006360, 58016033483, 45963075302, 55700002230, 63874031702, 52959082230, 68071300908, 680714990, 65841082516, 604290083, 63874093901, 60760023030, 54868514803, 63629139804, 68115016160, 51129941601, 662670100, 58118006508, 68071521700, 58118233208, 65243032509, 66336026990, 68071000860, 530021111, 58854002730, 67544061394, 500903429, 52817012250, 661160432, 50090088802, 68084011111, 61392059630, 58016069171, 64720012511, 63629642102, 47679056135, 649800281, 58016054344, 59762054201, |

| | |
|---|---|
| | 63629407104, 58016054324, 51079081117, 636297249, 67651002400, 58864095330, 54868545701, 58016054467, 58118834403, 61392011951, 53217028530, 68001017900, 680714020, 46193069810, 51813025730, 63874043230, 66116036060, 58016037809, 65427034403, 557000395, 51079087301, 58016038691, 58016038342, 68071218909, 50436646303, 54868521002, 65841082606, 61786015719, 619190715, 63874066504, 65841082406, 68071001730, 64909010407, 64764030290, 65841082616, 68071302409, 521250215, 500901526, 51655096125, 58016037887, 65243034609, 54868326504, 59762372501, 62939321101, 51138033830, 61392011960, 53978201408, 64909010222, 68084078811, 61442011525, 54441006950, 58016034490, 54868524300, 548685364, 52555029301, 637390119, 551545224, 58118145108, 532170285, 58016038336, 63874066501, 58016038391, 49999011300, 61442011710, 53808040801, 58016087650, 67253046010, 55154455609, 58016000503, 58016037830, 54868326501, 500900505, 47679056201, 64909010109, 680711944, 680840745, 53978201405, 55289097614, 58016054367, 60760002498, 49884096701, 53002417006, 67544030273, 63874031714, 68071409303, 58016044780, 57237002405, 50436002402, 64720012311, 50090248502, 58016033456, 55154317506, 58016037812, 54458097210, 60491028434, 58016044745, 54868528800, 58016069191, 59762702209, 52446047523, 59930162203, 58016038615, 61392059660, 55154622409, 52125047923, 68115015560, 614420115, 50090050505, 51655012052, 63187064190, 67544056632, 49999011330, 65862008205, 557000722, 60429008301, 66993082230, 51138036730, 597622331, 68071499009, 68071323903, 680714730, 636294071, 59762702109, 59762372707, 62318014201, 65862008036, 58016037632, 54569561800, 65427003302, 68071412408, 636292204, 51655018026, 500902222, 55289080686, 68084029521, 597627021, 58016044720, 51129350901, 58016038626, 63874031728, 65427037603, 61392059330, 53489015101, 58016054425, 58016033408, 58016069114, 58016033425, 50090207601, 54868337703, 58016044790, 55111069605, 58016038300, 50090317203, 55154827009, 68071314106, 597625031, 52446047421, 55154547600, 66116044060, 68084055621, 61392072460, 53808025801, 551546238, 58016069108, 67544012992, 47202250801, 58016069196, 504360908, 607600024, 68084013701, 67251046011, 64720016410, 64980028010, 64764030433, 60760024890, 521250765, 62528034480, 59115001455, 50090198701, 63874031740, 68115015530, 68071444408, 581180141, 58864021460, 58016054302, 50090155001, 52125010502, 54868539901, 52125097119, 58016037824, 64720014210, 65427034401, 68071444406, 68071309709, 63304042530, 63874031601, 60491041360, 58016005830, 65862008230, 50090152600, 58016038310, 63874031715, 61392011945, 51655006324, 54569599300, 62037087305, 58016069192, 55045236108, 65427003301, 55887072730, 52817012010, 65841066001, 68084011211, 66267010312, 49349033820, 61919037860, 63629139807, 58016054432, 65243037806, 63629101705, 552890265, 58016044716, 658620028, 62318014200, 66267010290, 607600175, 65841082510, 58016054409, 58016054404, 55370046110, 58016038624, 58016038620, 58016038676, 658620030, 59115001299, 58016037683, 50090429702, 49884084610, 55289089298, 58016033426, 51655096124, 55887021290, 58016037618, 572370024, 53217028590, 58016037640, 55045213806, 58016038344, 54569860101, 58016054401, 56126012211, 63629304303, 58016037842, 68071445306, 65862002801, 58016038345, 64909010422, 58016037624, 67544051130, 62584005250, 58016087600, 658410825, 68115015960, 52544056005, 62269029424, 58016069173, 58016037872, 58016038642, 55289089286, 551110321, 54868498802, 61392070930, 58016033480, 51079029360, 63539015557, 53746028601, 53978069305, 55370014907, 54569383108, 55289080660, 68115016140, 58016069110, 63304042710, 54868524302, 680840788, 58016033414, 54868331902, 55700046930, 58016054399, 619190448, 58016037848, 67253045250, 58016069128, 57866641101, 61919071390, 55289080693, 49349033823, 55370046211, 58016000590, 68071300903, 54697007904, 51079087220, 68084032711, 548684420, 631870234, 51079029220, 61392006454, 58016069190, 58016044724, 58016044740, 51129374102, 51285032502, 502680358, 58016033430, 58016054309, 51129203301, 53002041730, 557000725, 619190287, 62037087201, 631870486, 58016038377, 607600745, 49884096801, 60491027880, 636291017, 680713239, 55370059408, 58016038692, 58016037680, 55048024830, 55160012205, 59012015530, 52934020000, 61919033060, 51138027030, 68084013611, 66105098602, 50090162600, 67296126703, 51129205501, 58016069112, 65841083406, 58016033496, 57866646302, 50090050504, 58016037827, 58016087620, |

| | |
|---|---|
| | 68001017803, 58016037816, 63629279306, 68071482801, 51655090425, 60429008518, 500902556, 63874031690, 58016054492, 50436707301, 54569554800, 65427087305, 57866640901, 66105098603, 52152013302, 58118145209, 548683334, 63874058880, 55048027030, 68115015440, 64720016950, 60491019230, 50090050200, 68071410906, 50436646204, 680714223, 54907042701, 58016038603, 63629139406, 516550105, 62540725504, 66336073030, 557000256, 60491027901, 63874031604, 61392006445, 58016038630, 54868498804, 51813026099, 58016037644, 64720014011, 54868420604, 54569393801, 52544046101, 55045226700, 58016038327, 55111032205, 55370046010, 50090049600, 63629139401, 58016033436, 47837005801, 604290920, 58016054376, 58016033477, 55887017330, 60491064634, 54868420600, 54868460900, 58016054370, 54868102000, 58016069193, 63629647601, 68071240203, 500902031, 60491041234, 67544012994, 49884012402, 68071445209, 658620581, 58016005801, 68084032601, 51129374101, 58016037879, 52985007701, 60491041405, 67253046050, 47837005810, 50090317204, 58016037690, 68071444403, 64720029111, 65862002817, 68071408503, 58016037601, 55370050609, 500904297, 63187081160, 504367075, 54868546701, 61392059239, 48817700201, 55111032005, 62528034470, 67544056690, 55111032030, 53808067801, 61392072431, 58016054312, 58016054445, 68115015730, 68071199106, 68071194208, 65862003030, 65841083330, 50090198703, 552890806, 58016054482, 66105075410, 58016069144, 499990401, 63629139403, 65841083306, 65862002900, 54868333401, 50090155000, 54569860103, 50090248501, 53217031799, 68071449802, 521250573, 60429019301, 52985001702, 50090207500, 64720016850, 64720016511, 65841082505, 49999010730, 58016037628, 581181451, 57315001004, 51129381801, 66267010391, 51129302401, 51813006460, 51129455501, 530024460, 49884074505, 55111032078, 49349063502, 58016038680, 65427087201, 61919028630, 50090198702, 54569561902, 61392006354, 51129349702, 64980028110, 68084055611, 538080260, 63874031630, 64720014110, 62682500408, 66105098650, 52934020200, 51129179001, 55887096930, 67651032100, 557000022, 53978201303, 55700046960, 50090110403, 63629315802, 58016054491, 58016054316, 58016054318, 619190286, 63874031701, 58016054398, 67544036470, 58016033491, 521250766, 51285059804, 61392083430, 62682500403, 51129350902, 49999010790, 65243018509, 65427087205, 68115015430, 61786037319, 62269029129, 55289030193, 548685467, 62318014102, 68115015540, 51813023899, 58016054473, 53002111103, 54868539900, 52817012150, 49999078190, 55048024730, 68071464301, 68071002805, 68071196703, 57866640701, 597622332, 55700046990, 66105075403, 631870811, 675440097, 59115001399, 65084015010, 58016037850, 63629101605, 66105098503, 61786002320, 63629641002, 55154599206, 55289077907, 58016054420, 63629101701, 63874058801, 516550337, 49884084501, 55700002290, 54569020602, 51285059904, 52125097102, 63874093960, 54868452901, 557000612, 68071194406, 52125097223, 54569561802, 58016038399, 64764030247, 538080258, 58016054380, 670460236, 68071194409, 58864070530, 57866640905, 68084013811, 61919071590, 55154053408, 65862088930, 64980028005, 57480039101, 65841082477, 67046023614, 55111032079, 55829051010, 50090203001, 55887036890, 58016037825, 53002041720, 58016054427, 58016038330, 58016037818, 59762054101, 532170196, 51655081026, 65841083401, 52152013502, 55289089214, 64720029210, 493490730, 59651026905, 64909010208, 51079029160, 68084080711, 60760014160, 636291255, 51129286701, 58016054498, 65862003001, 67544056680, 66267010060, 61392059531, 58016044721, 54458097110, 52189029224, 64725005801, 52125064602, 49349073002, 68071445302, 51129349701, 63629647602, 58016054372, 54569607202, 57315001104, 58016087614, 65427015566, 52555029101, 55887065760, 68071194402, 61786067702, 58016037604, 500904008, 54569561904, 59762233007, 58016037642, 58016069126, 52125097219, 62682500201, 54868536400, 58016054444, 57866660501, 581182331, 68071300906, 58118032108, 61919025060, 49999010720, 54868402601, 68001017800, 658620888, 54868573900, 51655006352, 50436646302, 619190418, 51655090424, 57866707401, 58016037815, 58016033475, 607600230, 58016041190, 58864095630, 55700072590, 66267010091, 680711773, 58016038387, 58016069187, 63629139402, 67544056670, 61392059339, 50268035911, 55953034270, 58016054321, 58016033428, 500905377, 52152013504, 50090049601, 52125019902, 58016033421, 67046023530, 52125097120, 61786015723, 58016038616, 504366462, 58016069104, 65841083416, 63187084860, 58016037616, 50090050306, 67544019980, 55048024890, 63629279301, 50090050400, |

| | |
|---|---|
| | 58016033400, 58016033470, 54868452903, 51285059805, 64720012310, 65841065901, 58016087640, 50111058401, 58016038320, 58016069115, 67544009753, 619190722, 59115001499, 61919037890, 55289089290, 57664039818, 64764030490, 67651032200, 63187069890, 52728017810, 528170120, 67046023730, 51079081024, 47202251002, 500901550, 54868409103, 50090222202, 63629125501, 510790426, 680715217, 53217028599, 63629279304, 61786056219, 58016038681, 548684842, 58016054410, 58016054442, 58016054496, 61919044860, 58016037030, 60429008330, 61392011990, 68071412409, 51316010002, 58016054479, 49999057160, 500905040, 62939323100, 51813025799, 493490037, 49349017002, 636296413, 50090580602, 54569599100, 521250199, 50436640802, 58016037684, 58016038667, 50090207603, 67544012970, 55111032105, 58016037676, 49884074501, 66267010360, 493490338, 58016054307, 65862008105, 53217031730, 52959044960, 55700062430, 60951071485, 499990107, 493490092, 51129323502, 55154267703, 500900494, 50436707303, 58016054414, 63187048660, 68071222806, 54569554700, 63187025560, 60491041468, 51079042501, 49884045205, 63874043210, 50090049509, 55700025690, 60760039430, 65243017612, 61786073020, 58016054305, 50090050203, 581188343, 55045226608, 66267009930, 67544065390, 59012041273, 521250767, 54697007903, 58016046790, 50090222000, 551110320, 597625033, 68071218106, 63874031712, 66336066290, 58016037673, 54569445300, 60491019234, 680010178, 50090152604, 51138047430, 67296124703, 68084078821, 65427087301, 55289080614, 50090152603, 55887084560, 67544076460, 53978069403, 52544055901, 58016069167, 61392011930, 66105075406, 59762702101, 63187025590, 500903012, 50090255600, 67046023621, 58864002760, 61392070965, 551545992, 66105075606, 58016037890, 54868331806, 63629647603, 67046023607, 51138027130, 59762378301, 49999040160, 65841083201, 631870667, 680713024, 58016054477, 47202250901, 65862003099, 63187084830, 58864016130, 67544030298, 58016054499, 502680362, 50090222201, 66267009990, 64980028001, 52544055905, 61919085090, 55154817500, 60505014101, 58864002714, 50090342908, 65841065930, 493490719, 60429008230, 51129196601, 58016054403, 58016044702, 65862058199, 504367074, 65243037818, 66267010020, 58016037698, 54569607200, 50090088804, 65084016610, 55829051210, 58016038699, 51129407701, 63874031710, 63304042501, 66336031930, 58016037821, 55045213800, 58016038304, 58016044703, 58864085830, 61392011931, 58016033484, 548684091, 59762702201, 55048024360, 58016038683, 61919072060, 61392059690, 581181452, 65862058101, 65841082430, 55160012301, 658620029, 58118834400, 55154827000, 532170323, 67046023760, 51138047230, 58016087621, 65862008130, 55289077990, 551540534, 510790810, 64720016910, 55154317500, 63304042610, 59762503201, 680711942, 58016038393, 52125058302, 504366409, 572370021, 64720012510, 53467017100, 61392070990, 58118032106, 65862058099, 55048024430, 58118032203, 532170320, 636295598, 60491027968, 55154317504, 631870659, 68115015460, 58016054383, 58016044709, 58016033481, 619190378, 63629279303, 65243028518, 54868326505, 548683265, 551547987, 591150014, 50090110401, 62584005200, 63629407101, 619190723, 51129402201, 63187047790, 636291016, 68071194403, 548685188, 58016055500, 51129191301, 50090228202, 61392070925, 51129324701, 58016037630, 61392011791, 53002044630, 50268035915, 51129354602, 52985006001, 516550180, 54868536402, 55289097698, 51813010699, 50436630201, 58016038315, 63187025530, 50090318904, 61392006332, 58118046100, 50090318901, 66336053490, 647200123, 557000585, 58016054348, 63739011715, 68071473003, 66105098510, 46193069802, 54868326603, 53217019660, 50090248601, 55288124301, 55370046150, 557000576, 55370014608, 58016037856, 58016037635, 55289089293, 51813028760, 548684988, 58016044796, 61919044830, 58016054336, 55111032295, 680714444, 58016037679, 58016005804, 54868331900, 658620080, 67544009792, 55111069610, 54868518500, 60491027868, 67544112970, 548685712, 68084029511, 61919072390, 67253046250, 50090085400, 63629304302, 49999080760, 63739011601, 58016054456, 59762233106, 57664039813, 63874043201, 61392006460, 50090222200, 51655096224, 63874031602, 52125097123, 58118834406, 581181453, 68071155703, 63629125502, 66105098606, 58016038675, 58016037609, 53978018309, 58016069145, 67544087580, 500900888, 49884074601, 61919033030, 58016033487, 57237002305, 55887017990, 58864002790, 607600141, 58016037889, 61392006491, 58016044710, 58016038335, 66105098609, 55289030190, 63874043214, 55048024630, 58016079630, 51655010652, 49884084510, 58016041160, 58016005800, 58016054397, 58016037689, 54868099701, 631870848, |

| | |
|---|---|
| | 58016038369, 58118233200, 614420116, 54868037301, 51129128801, 55154622408, 54569554801, 68071401006, 52959093660, 500901987, 521250389, 59115001210, 50090306902, 51129402202, 500902075, 58016033460, 60429008530, 66336085030, 51655010626, 68071503000, 54868124404, 658620580, 58016044700, 57237002299, 65841065910, 60429008560, 61786098323, 55887069390, 66267010230, 51129446701, 58016033450, 58016054300, 68071196702, 55289089230, 49884096901, 55700039560, 63187064160, 63629565002, 58016037875, 51129381702, 58016069100, 61392006339, 63874031620, 67544065380, 62318014202, 607600248, 53217025060, 58016044712, 68071196706, 68071445309, 58016044718, 61392012054, 47837005805, 649800280, 51138047330, 52544046110, 52125021502, 51129171901, 516550942, 54868524301, 54569554701, 66105098611, 493490272, 68071409308, 552890281, 52544055805, 67787020501, 67544061353, 66105075609, 54569599301, 63629220201, 50090050201, 53002446006, 50090504002, 50090419900, 58016037671, 53467675500, 55111032230, 64720012411, 58016033476, 58016000504, 55289061460, 597622330, 51129014501, 502680355, 55370059507, 62584011000, 63874058860, 67544075660, 55289077930, 61919025004, 68071199103, 68071444409, 50090088703, 50090291501, 65841082530, 65427037707, 55289028160, 59762233003, 532170250, 521250481, 65243018512, 66105098402, 54569585502, 68071469506, 647250139, 55045226508, 58016033469, 45865071072, 65862057905, 53217032090, 54868124501, 54868326602, 60760032930, 54868409101, 58016069176, 58016038309, 65427003202, 68084032611, 617860043, 67544030292, 63629684001, 52959093630, 58016044776, 63629630903, 57664072518, 58016038601, 55887027330, 67544065392, 66116044030, 58016038600, 53978069407, 61786046702, 52985007801, 58016037605, 67296124603, 58016084430, 58016037612, 58016038605, 52959082200, 58016046730, 58016069175, 670460237, 54569020500, 53978069303, 49999078130, 58016038371, 66267010390, 58016037606, 63187099360, 61919072290, 65243028512, 68115016120, 51129390801, 53978201308, 54868079500, 636297093, 502680361, 67046023728, 55289089260, 58016037805, 658410824, 58118233209, 58016069121, 58016037807, 51655075877, 57315001102, 680715030, 50090108200, 55289089215, 499990113, 58016037625, 57315001103, 68115016040, 55045156809, 54868099702, 493490175, 65841083405, 50090318903, 53978201403, 516550810, 551548175, 50268035815, 51129444101, 68071177301, 521250392, 58056014610, 63629709201, 50090249200, 66336057330, 61919041860, 53978338309, 663360712, 58016054421, 58016037801, 51129286702, 53978336209, 62682500603, 548683426, 59762378201, 53217019630, 680713068, 58016069179, 59115001301, 538080257, 50090049400, 493490635, 551546224, 59012041141, 617860983, 607600394, 57866707301, 60429008512, 58864022493, 67544065370, 61442011510, 63629220501, 53489015110, 57664072718, 63874031724, 65862008001, 61786074520, 53217031790, 458650574, 58016037832, 58016087630, 64720016610, 47679056335, 50090306900, 58016069118, 58016038645, 65841082601, 65427037704, 500902915, 54868518801, 67253046210, 67544009780, 58016038321, 63187047760, 63629709301, 61392072490, 67544075180, 62037087205, 63187099390, 50090301209, 63187066760, 55370014909, 61786028619, 60491064530, 59762372704, 65862003000, 50090100400, 504366463, 55289089201, 66336066230, 55370046050, 50090088800, 58016037808, 61442011701, 58016033405, 58016033412, 50090491101, 544580967, 61392059439, 58016038606, 54569384100, 58864022430, 55887096530, 55154547607, 52125048119, 510790872, 58056014605, 58016054470, 58016038612, 62584044900, 50090050401, 60951071470, 58056011710, 53978069408, 548683319, 50090088801, 68115015420, 63874058830, 54569469501, 54868514802, 59762372606, 68001017700, 68071258909, 49999080730, 55111032179, 58016044770, 63629139304, 591150013, 60491027960, 55111032178, 66267023030, 66336053430, 58016033492, 65862008101, 54569369000, 658620081, 58016054424, 672530462, 49884084601, 52125076619, 58016037877, 55700039530, 64720014111, 669930822, 58016038635, 68115015520, 65841083410, 53002041760, 63874031615, 61919028772, 55111069710, 58016038693, 63874066560, 68071000830, 58118834308, 62037087301, 647200125, 55111069501, 58016005890, 54569384201, 58016037656, 58016044760, 68071409309, 50090198704, 68071238508, 548685457, 55887022230, 68071306809, 521250393, 60491028330, 50090203000, 66336084630, 50436646301, 54569607201, 50090085401, 55953034480, 58016054381, 54868498800, 53978069405, 58016069140, 617860226, 60760022830, 521250479, 61919028790, 51129469202, 67544009794, 59762375704, 58118014108, 54868546700, |

| | |
|---|---|
| | 52544046005, 57866640801, 67544061390, 50268036115, 64720012410, 54569607203, 58016037803, 55160012201, 59115001355, 50090577600, 64720014150, 50436640902, 57866640903, 58016037608, 58016037844, 54868337701, 58016069181, 65084018510, 55953034380, 65862089001, 63187065930, 647200142, 63874031681, 58016037867, 58118032200, 53217028500, 680712402, 49349003702, 58056012266, 58016069109, 58016038390, 50268036111, 67787020401, 53808025901, 53217034990, 647200290, 52728022793, 61392006325, 68071300909, 60429008210, 58016054314, 67544065398, 65243037803, 55953034470, 51138025130, 65084018410, 60429092001, 67544054882, 58016038628, 67544065360, 61392072430, 57664072413, 68071408509, 55154120100, 54868333501, 51129316902, 63629724801, 552890606, 53467675503, 66267023140, 65427041166, 54868124402, 55048024130, 504367073, 60429008312, 53217019690, 596510268, 58016054350, 63629101501, 65862008118, 52934020100, 58016037699, 680714452, 50090207502, 572370023, 58016038397, 58016044736, 59115001310, 63629407102, 65243017609, 58016033435, 58016037892, 63874066590, 58016038307, 60505014108, 61392083431, 58016033402, 52544046010, 58016037876, 58016038670, 60491019260, 60760016530, 51285032702, 58016087624, 59762233203, 60760074690, 68071402002, 54868124401, 59762378302, 57866646002, 50090050406, 54868514800, 504360872, 68071437808, 55887053699, 581180322, 661160427, 58016037677, 51813017730, 500902030, 67544056694, 55045361304, 60429019201, 631870993, 61786098119, 58016033442, 58016038610, 617860981, 68115016130, 51655010625, 51129349501, 54441006910, 61442011625, 50090248603, 58016038697, 58016069101, 52125097220, 68071199109, 54868546702, 65841082410, 67651032207, 493490228, 65841082677, 67544080860, 58016037899, 63304042630, 63629139301, 54569383106, 521250583, 55829051110, 63739011815, 50090085402, 68071413306, 58016054315, 51129111401, 68071471803, 55887072760, 658620082, 521250646, 58016054430, 54868571200, 63629642104, 551540539, 55289080698, 58016054389, 59762375707, 51138047530, 68071401003, 63629139201, 65841066116, 54868337700, 49884096905, 59012015566, 64720016911, 58016038689, 58016037869, 58016054472, 55045213808, 59012041173, 611450202, 58016038373, 63304042730, 54868442000, 59930163902, 59762233103, 58016044701, 60429091801, 55887033930, 51079087217, 68071043506, 59651027099, 67787020301, 617860730, 51079081020, 662670103, 49999010830, 52544055810, 617860157, 66267009992, 52544046105, 62318014100, 680714828, 65841065916, 49349017502, 680714869, 61919047630, 57866799101, 63874031720, 51129172001, 51129390802, 65841083316, 63629642101, 55045343408, 57237015305, 53002446206, 60429008360, 54868124400, 58016038614, 52125072820, 54868331905, 51129302402, 61392011954, 55111069510, 57237002105, 619190330, 53002446000, 62269029124, 50090050405, 58016054377, 58016037621, 647200124, 61786004319, 55370059607, 68071437803, 50090207602, 510790811, 669930821, 58016038608, 53978018308, 680714085, 47837005700, 51079087320, 54569561901, 49999010860, 54868331802, 680714010, 50090050302, 55154619600, 54868326604, 50090203102, 58016038398, 58016054320, 55111069705, 64909010207, 631870255, 63629565004, 62540725401, 49884045101, 63629315804, 54868514804, 50090248500, 53808062502, 68071238506, 58016038375, 60951071170, 50090203100, 58016033482, 66336053590, 51129365202, 54868124502, 58016041130, 55887021230, 52152013305, 49999011301, 50436646202, 68071412403, 68071410903, 62540725501, 58016069189, 61919044890, 65862058001, 67251046050, 60760017630, 538080400, 54868484202, 68071302408, 51813023860, 58016044704, 68071412406, 67544076432, 58016037871, 68071302403, 521250388, 61392006330, 51138026930, 58016044726, 58016054396, 61392012060, 53002446200, 58016038324, 50090228200, 658410833, 68071499403, 67651012300, 59651026930, 66993016402, 50090049608, 51129323501, 66105098403, 66267010330, 58016037650, 59762372603, 68115016190, 51129444102, 58016054304, 548683318, 51129316901, 50268035515, 58016037835, 58016038379, 55887033990, 61392006390, 50436090801, 500900503, 50090049502, 46193069701, 55887053630, 636299281, 50268035611, 68071258906, 71335205003, 68788813808, 68788819209, 705183688, 71335184802, 68788813809, 71335188602, 60687068201, 71335205004, 70518057906, 68788819206, 70934095960, 53002411100, 71335184805, 71205077930, 70518164601, 625590316, 516550983, 71335205005, 516550383, 51655098226, 713352050, 68788813806, 68071408508, 50268035511, 51407062110, 62559031610, 71335188605, 50090632204, 50090632203, 705183504, |

| | |
|---|---|
| | 68788819201, 68788819208, 51655049252, 516550966, 50090100601, 50090639401, 50090699600, 70518370900, 51655065952, 52817038510, 71335240904, 68788863403, 50090685200, 71335974204, 60760079430, 72603011603, 705183709, 68788850801, 68788834309, 51655038326, 53002446100, 82868004290, 72888011701, 71335974201, 705183722, 70518350401, 50090674600, 59651078130, 70934086230, 705183719, 51655049225, 71335240903, 70518403500, 70934057060, 721622102, 713359742, 726030116, 500906765, 500906426, 721622103, 500906394, 721622215, 680713452, 68001058600, 687888243, 726030117, 680713548, 500906910, 516550565, 72789037030, 607600793, 500906607, 721890485, 500906746, 500906473, 712050779, 687888341, 687888343, 528170385, 500906975, 50090641504, 51655098352, 70518055203, 687880141, 709340098, 72789013090, 70771109803, 70786000303, 691890524, 68788641109, 70518184800, 76333015412, 70771109906, 70518001900, 70518269501, 713350114, 69543012350, 71335018001, 68382018601, 70771110000, 705182888, 70518011502, 71335006902, 71335015001, 69543012450, 686450151, 72189007090, 71335103102, 68382065616, 712090010, 68788972801, 68382065810, 70518057900, 71335038802, 71335036005, 68382033610, 71335103103, 68788806603, 71335003602, 70518241200, 70518035801, 713350590, 68788797001, 71335061803, 70518002902, 707711100, 76333015413, 71209001105, 71335061807, 70518040500, 71335096703, 705181848, 68382065577, 68788785606, 71205027090, 68382018616, 68382033714, 70518055200, 68382018401, 713350851, 71205027060, 68788991809, 69543012510, 71335015002, 705182308, 70771109805, 68788691508, 716100393, 71335159202, 68788809501, 709340855, 75834020201, 705180373, 70518112402, 68382065301, 71209000911, 71610017560, 70518099803, 68382065816, 687889966, 709340237, 68788046103, 709340272, 68382065877, 68788014103, 68382033506, 71205004178, 68382065830, 71610038880, 70518241201, 71610039360, 69189000701, 70934084390, 71205027030, 70518288801, 68788809509, 68382033605, 705180116, 672962042, 70518267003, 71335015006, 71335018002, 71114022200, 68788641101, 68788797003, 75834020300, 68788681801, 705180389, 68788046006, 68382033706, 68788809506, 705180907, 70518037300, 76237017130, 722410040, 68788718006, 72241003811, 70771109802, 68382033577, 70934030030, 69189936401, 709340569, 68382065710, 71335003902, 70786707303, 71335003906, 68788685301, 70518011501, 68788785609, 68382018630, 70518055202, 705180631, 68382065516, 68788917609, 687887694, 707860900, 71209001111, 68382065506, 68788681808, 71610010560, 71335159201, 71335061804, 71335048401, 70934071590, 68533003645, 70518264400, 758340202, 71335011401, 691899364, 705180405, 705182695, 683820184, 705180029, 686450572, 694520128, 71335003601, 71209000905, 68382033601, 70518011600, 68382065401, 70518011500, 68382065430, 687887250, 69452013030, 68382033606, 71114022105, 713350127, 68788755306, 713351778, 71610010553, 72241004010, 70518090700, 71335038801, 68788769406, 687886411, 705182670, 713350039, 68382065501, 705180019, 68788797009, 70518057400, 721890453, 705181646, 68382033505, 68788996608, 68788641106, 687880142, 709340862, 68382033701, 71335003903, 71610017545, 68382065316, 68258101401, 68788972809, 69189091001, 76237016930, 68115061600, 71335096704, 71335061801, 68382033510, 71335003904, 68382065530, 687889918, 68382065510, 68382018677, 76333015512, 71335036004, 705180296, 762370169, 71335003606, 50090718701, 687886854, 70934030090, 70518099802, 68788991709, 71610039353, 68788691509, 68533003500, 68382065330, 621350585, 68382065310, 712050041, 71335003905, 68788014106, 71209001110, 683820654, 70518254900, 75834020200, 71114022205, 713351885, 713350179, 705182573, 71335006903, 68533003640, 68788971609, 68788996601, 68645057390, 68382033514, 70518106302, 68788971606, 70771109809, 621350584, 68788688501, 68788685309, 709340391, 68115062200, 709340726, 68382033730, 68788643607, 68788912801, 70771109902, 709340232, 68258101301, 68788725002, 71610010592, 70934032330, 709340300, 70934057090, 71335104202, 71114022100, 70518163900, 70934009890, 70518120100, 68788912808, 68788014203, 68382033677, 71335011403, 68382033516, 70518035800, 70518057401, 68382065605, 68788014109, 68382065305, 71335017903, 71335188501, 713350069, 70934027230, 68788685406, 70518090800, 70518099801, 72888011600, 68788971601, 71610039394, 70771109801, 70934039130, 68645057559, 71335003603, 709340173, 687887180, 713350036, 722410038, 70934017390, 71335177802, 762370171, 71335113004, 705181513, 68788014206, 71335177801, 687887710, 70771110009, 69452012820, 68788991801, 764390125, |

| | |
|---|---|
| | 71335036007, 70771109800, 68788755301, 687886801, 68115084000, 68788681803, 70786707301, 70518257900, 72241004005, 68382033501, 68788691506, 709340409, 68788991706, 705180115, 68788769403, 70518262401, 71335061806, 70518002900, 686450575, 683820657, 683820653, 68382065606, 68788643601, 71209001011, 71335061805, 71335093007, 763330156, 68788771006, 70518159401, 68788681809, 70771110001, 68788643609, 687888066, 76333015411, 71610038845, 687887970, 71610017594, 68788771003, 70786090001, 70518090701, 709340843, 705180998, 705181639, 70518267001, 71335018003, 713350980, 68788972803, 68382033710, 70934023230, 71610017592, 686450574, 76333015513, 691890007, 68788680106, 68788688503, 68382018430, 70518209200, 713351042, 68258105101, 71335015005, 70518011503, 68788119100, 70518106300, 71335036006, 705180552, 71335012703, 70518184801, 68788685403, 70518040503, 70518057905, 50090100603, 68788688509, 68788834406, 68382065777, 68788680103, 68788718009, 683820336, 709340167, 68788921606, 68533003412, 68788972806, 68788643606, 72789013030, 695430124, 68382065801, 68788809503, 68382033705, 70771110005, 68788642606, 71335096706, 705180288, 71335017901, 762370170, 71335003901, 70518035802, 68071218908, 71610017580, 71610039392, 76333015613, 68533003548, 68788771009, 70934009860, 71335965504, 69189145201, 68382033716, 71335011404, 758340204, 709340570, 71335085105, 71335015003, 71610038860, 76333015611, 705182412, 687886853, 70518037301, 71335012704, 71114022201, 68382065505, 68258101201, 68788797006, 71335103104, 68788642603, 76439012410, 68382018416, 72241003910, 763330155, 71335159204, 71335098003, 68533003600, 68788046106, 705180579, 71335036008, 70518267000, 70518164600, 72241004011, 68788806606, 70934072690, 71335003607, 71335096705, 71335059003, 707711099, 68788725009, 709340715, 71335006901, 68382033530, 71610039380, 71335096701, 71610038898, 713351031, 68788046009, 68645015159, 683820655, 705180104, 705180908, 713350967, 722410039, 68788096903, 71335012705, 68533003445, 69452012920, 70934086290, 71335113003, 69543012310, 68788785601, 68788014209, 68788971603, 70934069030, 71610017573, 68788912806, 71335188502, 70771109903, 69452012930, 68382018477, 681510765, 70771110002, 68788723001, 683820658, 68533003400, 68645021154, 70518028800, 71335003604, 71610010570, 71335017905, 70518010400, 68788119106, 71610010594, 68382065377, 68788119103, 705181121, 68788806609, 71335036001, 716100175, 71335085103, 68788014201, 68788921601, 764390124, 69452013020, 71335085101, 68788725001, 68382065477, 686450211, 68788723003, 71610010580, 709340690, 705180456, 71335011407, 68382065410, 687888095, 70518057903, 705182644, 68788725003, 713350618, 68151076500, 721890070, 70518038900, 71335113002, 70518402501, 70786000301, 70518055204, 70771109900, 68382065610, 70771109806, 70518112100, 71335012701, 68382065730, 71335003907, 71335011408, 68788014101, 70518288800, 727890130, 686450210, 68788755303, 68382065677, 687889176, 68645015054, 70771109901, 68788996609, 70934023730, 687886885, 71335093002, 69452012830, 70934017330, 70518040502, 68382065806, 68788643603, 70518057902, 70518002901, 691890910, 70518230800, 691890845, 687887553, 705182196, 68382065705, 68382065805, 70518209201, 705182092, 71335093003, 70934056930, 70518029601, 683820656, 68788769401, 68788096906, 70518045600, 68788680109, 68382065406, 683820186, 686450573, 75834020401, 70771109905, 68788685401, 713350150, 71335093005, 686450150, 68788912809, 70518055201, 70934085530, 70518082700, 705180358, 71335059002, 71335104201, 76333015612, 70934016790, 68788991803, 71209001005, 75834020400, 694520130, 72241003911, 70934039190, 70518011601, 71114022101, 70518267002, 687887230, 687886818, 607600800, 70518090801, 695430125, 71335096707, 68382018577, 758340203, 70518106301, 68645057454, 709340323, 694520129, 71335098004, 70518057904, 71335048405, 70934072790, 68788834403, 76439012510, 70518378203, 68382065706, 71335018004, 71335011405, 705180574, 68788996603, 71335048403, 687886915, 71335061802, 70934072630, 70518090702, 68382033616, 687886436, 705182579, 712090011, 70518269500, 70934040930, 71335096708, 68382018516, 70771110006, 71335188504, 70518159400, 71335188503, 75834020301, 71209001010, 68788991701, 70518040501, 68788046003, 705181201, 71610017598, 716100236, 69543012550, 68382065416, 68788685409, 71335093004, 713351130, 707867073, 70518029600, 727890048, 68382033614, 68788991703, 71335093006, 705182624, 68788691503, 712090009, 68788718001, 713350484, 71610038892, 68788996606, 71205004130, 70518151300, 71335017904, 68788688506, |

| | |
|---|---|
| | 70518112400, 70771109909, 687887856, 70518257300, 68533003440, 70518057901, 68382065701, 70518112403, 68382033630, 68788685303, 71335017902, 68788725006, 71335103101, 76333015511, 70771110003, 691891452, 68533003540, 707860003, 70518120101, 68788917601, 687889128, 68788691501, 68533003512, 687889728, 68788685306, 68788681806, 687889216, 713350180, 713350930, 68788991806, 76439012310, 71335085104, 68382065601, 713350388, 71205004190, 70518262400, 709340727, 683820185, 68382033777, 68382018410, 68645021054, 68788642609, 72789004830, 70518035803, 68788917606, 70518063100, 69189052401, 72241003805, 712050270, 70518366400, 68533003612, 687889716, 71610023660, 705181124, 68788641103, 683820335, 72241003905, 713350360, 71335036003, 68788769409, 71335177803, 68788642601, 716100388, 764390123, 69189084501, 68382018501, 71335038803, 70934016730, 71335159203, 70934066630, 68645057290, 68788917603, 68788723009, 70518002903, 705181594, 71610039370, 71335059001, 68788680101, 68382065405, 68382065306, 71335015004, 70518112401, 68788921603, 68788718003, 76237017030, 70771109804, 70518045601, 621350583, 71610038873, 763330154, 68788723006, 68788912803, 70518163901, 70934023760, 705180827, 70518099800, 71335098001, 71335048402, 68788755309, 71335098005, 68788785603, 70934032390, 707711098, 68382018530, 71335093001, 71335038804, 70518219600, 68382065630, 71335085102, 70934030060, 687886426, 71335048404, 687889917, 71610023680, 68382018510, 70934084330, 70771110004, 716100105, 70771109904, 71335113005, 68382018610, 713351592, 71335012702, 71335113001, 683820337, 705181063, 70934072730, 71335003605, 68788921609, 69543012410, 709340666, 71335011406, 71205004160, 71335098002, 68382065716, 71335188505, 705182549, 70934009830, 695430123, 70934087860, 63629928101, 713351886, 71335184804, 516550982, 71335205002, 70518262402, 71335184803, 60687068211, 68071486908, 70934066690, 70518350400, 606870682, 68788843209, 625590315, 71335205001, 709340959, 68071408506, 68788819203, 514070621, 713351848, 70934087890, 68071349300, 68071422303, 687888138, 71335188603, 62559031510, 63629525603, 68788848701, 71335965402, 71335188604, 70518184802, 71335188601, 687888192, 68788813801, 71335184801, 68788813803, 71335965704, 70518403901, 680010584, 50090619600, 680010586, 606870701, 500906196, 680010585, 71610068092, 516550489, 71335965403, 59651078230, 500906996, 719210215, 70518362700, 60760080090, 50090639400, 70518378202, 68788863406, 82804006560, 500906852, 72888011705, 70518364500, 621350731, 500906848, 72888011601, 705183652, 72603011701, 68788834301, 70518362701, 72189048530, 728880116, 705183896, 719210216, 68788843203, 68788834109, 705183782, 71335240905, 50090691000, 51655056526, 72888011700, 71335965405, 50090684802, 68071363908, 68788851203, 68788834103, 50090642604, 728880117, 50090674601, 71205077378, 500906849, 500901006, 71335018005, 713359657, 680713492, 50090100604, 71335965505, 705183786, 50090647304, 68001058603, 68071345203, 705183665, 68001058503, 72162210300, 719210214, 705183627, 71335965705, 70518412800, 51655056552, 70518365200, 50090100600, 50090684800, 713359654, 71335236703, 71335965503, 71335236704, 828680025, 50090641501, 51655096652, 687888508, 68788863401, 50090660700, 72162210200, 728880115, 516550492, 82868002590, 59651078205, 72888011501, 71921021650, 71335965701, 71335240902, 71335235302, 621350732, 71335965702, 71205077360, 712050773, 68788851208, 621350733, 60687076821, 680713493, 70518366500, 606870768, 72603011601, 68071349200, 705183664, 713359655, 71335235304, 71921021401, 500906415, 71205077960, 516550659, 727890370, 687888550, 68788851209, 53002417100, 72888011505, 705183645, 71205077990, 705183898, 68788848706, 70518378200, 70934085590, 70518371900, 67296204203, 70518378201, 72603011703, 72603011702, 50090599300, 500905993, 70710198706, 71610030060, 70710190003, 705183976, 70771179103, 70771177103, 50090550401, 707711792, 707101242, 707711790, 500906244, 500905616, 707711770, 707101240, 707101241, 707711791, 707101899, 707101901, 70771186908, 707711772, 707711771, 707101900, 691890725, 705182548, 691890478, 68258599009, 71610030009, 705182046, 70518204600, 69189047801, 69189072501, 716100300, 68258599003, 70518254800, 71610066109, 70518397600, 70771179009, 70771186006, 50090352701, 50090550400, 70710189903, 70710124203, 500901036, 70710189909, 70710124003, 50090550300, 70710198708, 71610030018, 500905502, 500905504, 70710178606, 70710178608, 50090347200, 70771179203, 50090624400, 500905503, 70771177209, 70710198606, 50090550200, |

| | |
|---|---|
| | 70771187008, 70771179003, 70771185906, 70771186008, 70710190109, 716100661, 71610066118, 50090103602, 70771179209, 50090561601, 70771186906, 70771179109, 70771177203, 70710178708, 70710190103, 70710124009, 50090550201, 70771187006, 70771177009, 70710190009, 50090561600, 70710124209, 70710124109, 70710198608, 70710124103, 70710178706, 70771177109, 70771185908, 70771177003, 458020238, 00006007814, 00597016407, 00597038577, 00006057582, 00006536808, 000060753, 00006007882, 00006007862, 458020499, 00597014766, 458020150, 00006027731, 00006057561, 00006053331, 458020169, 00006057760, 00597014010, 00006536806, 00006075331, 00006008062, 00006027754, 00006008154, 00597027012, 45802008765, 00006027733, 005970140, 005970148, 00597039031, 35356010330, 26053045101, 005970147, 00597039582, 00006008061, 000060757, 00006536709, 00597039552, 00006027728, 000060277, 000065368, 00597014070, 00006007861, 00006008182, 005970275, 00006008114, 00006008014, 00006075707, 00597039013, 00006022131, 00006077331, 000060081, 00597018270, 00006057562, 00597014660, 26053045001, 00006027727, 00006057752, 458020087, 45802010365, 00006057503, 000060533, 00006536708, 00006011201, 00597018290, 00597016430, 00006057703, 00006022128, 000060773, 00006057782, 353560103, 45802030465, 00006057702, 45802049965, 00006536703, 00006027714, 00006011254, 00597014760, 00006057552, 00597027094, 00006536807, 00006075754, 005970395, 00006008107, 00597027073, 00597014818, 00006007828, 00006075714, 00597014860, 005970270, 00597027588, 000060080, 00006536706, 005970385, 00597014030, 458020402, 00006536707, 00006008131, 00597038068, 00597018210, 00006057701, 00006075731, 00597027533, 00006077307, 00597038013, 00597014672, 00597018230, 00006053554, 000060577, 000060575, 00006022154, 00597014090, 00006057756, 00006536809, 00597039071, 00597014866, 000060535, 00006057762, 00006077354, 000060078, 00597027581, 00006057502, 00597018207, 00006027774, 00006057727, 00006057560, 00006057501, 005970182, 00006027782, 00597038588, 458020351, 00597014061, 005970164, 00006022101, 45802016972, 00006027702, 00597018239, 00006011231, 00597014718, 45802021172, 45802026065, 00597014666, 00006057556, 00597016410, 353560136, 000060112, 00597038586, 35356013660, 00597038022, 00006008082, 00006027730, 000060221, 00006077382, 00006075382, 00006536803, 45802035165, 00006053754, 12280040130, 00597014772, 00006008028, 00597016470, 458020211, 00597016439, 00006011228, 00597014618, 00006075782, 00006077314, 000065367, 00006053731, 45802040265, 00597039523, 000060537, 00597018203, 00597014872, 00006027701, 458020304, 005970390, 458020260, 005970146, 00006057761, 45802023865, 00006075354, 00597016490, 45802015065, 00006053354, 00006053531, 458020103, 00006057737, 00006057527, 005970380, 63629652401, 548685973, 64764025404, 55154504008, 64764012406, 64764025306, 55154503807, 647640335, 51138048130, 64764012105, 551545042, 58118027706, 55154504400, 548685840, 51138052730, 647640125, 50090408600, 64764012550, 50090554700, 61258011204, 50090103600, 61258027705, 64764025303, 500905547, 55154503707, 64764012590, 50090408400, 500903527, 64764025050, 551545041, 61258011203, 61258027703, 54868584001, 64764025002, 61258673900, 64764012306, 54868584000, 647640253, 64764025103, 58118027709, 64764062590, 50090557400, 64764012106, 500904084, 647640124, 50090561800, 551540410, 55154504102, 64764025030, 50090408700, 51138048030, 55154041008, 61258673800, 647640254, 64764025305, 51138052630, 64764025106, 54868646100, 64764025090, 551545044, 64764033701, 52125086102, 500905618, 54569592500, 55154504208, 548681097, 61258022103, 521250493, 64764012104, 54868109701, 521250861, 500903472, 64764033577, 500904087, 647640123, 55048043760, 500905517, 54868603100, 64764025406, 64764033580, 61258027708, 61258027702, 61258027707, 50090347201, 64764012502, 64764012404, 64764012103, 548686031, 64764033780, 64764012530, 500904411, 50090438300, 55154504108, 64764012304, 64764025104, 64764033760, 50090441101, 64764025304, 61258022102, 54868597300, 66105065203, 64764012405, 64764012305, 50090558501, 551545038, 647640250, 61258027704, 647640337, 61258027706, 52125049360, 64764033501, 50090558500, 500905585, 64764033560, 54868109700, 548686461, 61258022104, 51138047930, 64764025405, 500905574, 55048043460, 50090441100, 54868603101, 551545037, 61258011202, 647640625, 551545040, 64764033777, 500904383, 50090551700, 58118027703, 51129424302, 52125048660, 581180277, 64764025105, 500904086, 61258673700, 636296524, 64764012403, 64764025403, 51129424301, 647640121, 50090352700, |

| Code System | Concept Code |
| --- | --- |
| | 521250486, 64764012303, 64764062530, 647640251, 50090551701, 82381200008, 500906218, 50090645200, 50090621700, 50090618200, 500906231, 51407091830, 500906217, 66993036230, 500906183, 50090618300, 500906182, 50090623100, 00310621039, 71610017742, 50090621800, 701830221, 701830220, 66993045630, 50090705700, 82381217401, 701830240, 66993045730, 823812174, 50090645700, 66993036160, 50090449201, 50090438401, 00310620590, 58198531502, 51407091890, 701830241, 00597016407, 00310697530, 00597038577, 00006536808, 00006536407, 000065373, 005970153, 00006537403, 005970290, 00006537308, 00597029059, 00310628095, 00597029020, 00006537408, 00310626095, 00597015330, 00597030093, 00597030045, 00597015390, 000065369, 003106770, 00006537406, 00006536806, 00310622560, 00006536409, 00310692560, 00597017560, 005970280, 005970152, 00597039031, 00597029074, 003106780, 00597016860, 00006536907, 00310620530, 00310622594, 00597039582, 00006536709, 00597016818, 00597039552, 005970180, 000065368, 00006536408, 00310621030, 00597030020, 00597039013, 00597018270, 00006536903, 003106270, 003106975, 00006537409, 003106250, 003106205, 00006536308, 00310628030, 00006536906, 00006536708, 00597018290, 00597016430, 003106210, 00310678095, 00006537007, 00006536703, 00597029561, 00003142712, 000031427, 00597015918, 00597017566, 00006536807, 00003142811, 00006536303, 00597029588, 00003142713, 005970395, 00597015307, 000065374, 00006536306, 00006536706, 005970385, 00597015230, 005970300, 00006536707, 00597038068, 00597018210, 00597038013, 00310627030, 00597018060, 00006536309, 00003142813, 005970168, 003106950, 00310626030, 00597028036, 00006536403, 00597018230, 00597029578, 00597028090, 00006536809, 00597039071, 00597015370, 00006537407, 005970175, 00006537307, 00006536406, 00597018207, 00006537309, 00006536310, 00597016866, 005970182, 00597038588, 003106280, 000065363, 005970164, 00310677095, 00597015966, 00597018239, 00310626094, 00003142791, 005970295, 00310677030, 00310625030, 00597015960, 000065364, 00597015337, 00597016410, 00310699030, 00597015290, 000031428, 00597015207, 00597018018, 00597038586, 00003142714, 00310625095, 00597038022, 00310627095, 00006536307, 00310621095, 00006536803, 00310695060, 00597017518, 00003142814, 000065370, 00003142711, 00597015270, 003106225, 00597016470, 00597016439, 005970159, 00310626060, 00597028073, 000065367, 00597018066, 00597039523, 00597015237, 00006537003, 00597018203, 00310620595, 00310678030, 00006537306, 00006537303, 005970390, 00003142812, 00597016490, 003106925, 003106990, 003106260, 00003142891, 00006537006, 005970380, 500904364, 504580943, 500905034, 63629826002, 50458014030, 504580940, 50458014110, 50458094102, 50458094002, 50458094101, 50458014001, 50458054091, 50090348100, 50090449200, 551546932, 50458094202, 55154041208, 55154041108, 50458094302, 504580541, 50090502900, 500905029, 504580542, 50458054060, 50090348200, 500904384, 50458094201, 504580141, 50458054191, 50458094001, 504580140, 500904492, 50458054360, 551546933, 50090438400, 504580543, 50090503400, 50458014050, 55154142508, 50458094301, 551541425, 636298260, 50458054260, 504580941, 50458014101, 50458054291, 50090503300, 50458054391, 500903481, 50458014190, 50458014150, 50458054160, 50090503301, 500905033, 504580942, 504580540, 50090503401, 551540411, 50458014130, 55154693208, 63629826001, 55154142608, 55154693308, 551540412, 551541426, 50458014010, 500903482, 50090436400, 50458014090, 70183022007, 50090705600, 00310621090, 70183022107, 500907057, 500906457, 514070826, 500906452, 70183024090, 500907056, 70183024007, 51407082630, 70518244700, 70518198600, 705182447, 51407082690, 71610017709, 70183022131, 705181986, 71610017745, 705182046, 71610017715, 70518204600, 71610017730, 716100177, 82381217402, 669930457, 70183022090, 82381200009, 669930361, 63629325301, 70183022130, 70183022030, 70183024130, 70183024030, 669930456, 669930362, 58198531501 |

| | |
|---|---|
| RxNorm | 25789, 4821, 10633, 4815, 201057, 861737, 367762, 440287, 151725, 731457, 203680, 1156200, 1178082, 205828, 380849, 1157241, 669985, 203296, 1175881, 246524, 440286, 369500, 602549, 368204, 199245, 203295, 260287, 1171930, 849585, 207955, 1157122, 565410, 310537, 565668, 379565, 861733, 566056, 1171248, 565670, 574090, 153591, 607816, 1361492, 861740, 566770, 1008873, 731455, 310488, 1171934, 314000, 201921, 1156197, 566766, 568685, 566764, 847712, 201062, 1166403, 647236, 203679, 563154, 201058, 706895, 199246, 260286, 379559, 201060, 731461, 351274, 844809, 847722, 1179292, 1171234, 1171920, 1171933, 1179741, 669982, 865572, 669980, 647208, 429841, 352381, 313418, 565674, 374635, 700835, 566057, 647237, 565672, 310489, 647239, 1157121, 566768, 1170867, 1177973, 574571, 731462, 374149, 310539, 205877, 1178083, 315990, 198293, 379572, 669984, 1175878, 1185049, 205876, 881405, 205873, 881408, 1171929, 1178369, 565327, 564038, 1178368, 566762, 566718, 430102, 565669, 369562, 1361494, 205829, 1361493, 1171149, 1175879, 105373, 564037, 1165207, 602544, 153844, 440285, 379804, 566761, 1156199, 1186169, 881404, 205874, 203289, 881406, 1179291, 331496, 847707, 865574, 844827, 205879, 1168162, 1179740, 847710, 197737, 847714, 315987, 375952, 1157240, 372334, 1156201, 207953, 315980, 861756, 1165205, 865568, 861755, 153843, 881410, 378730, 565673, 1170664, 881407, 201061, 844824, 1157242, 861732, 1156198, 566763, 861745, 379803, 316834, 105371, 574871, 1185624, 1157243, 261532, 93312, 861743, 1170866, 881411, 310490, 847724, 647235, 225613, 881409, 284433, 369557, 369373, 205831, 574089, 861757, 574612, 566769, 606253, 198292, 847716, 379570, 1171148, 669987, 861741, 1166404, 861748, 847706, 379802, 284431, 198291, 861747, 565671, 316832, 217360, 151466, 205881, 1175659, 153845, 201922, 315991, 205880, 261351, 861736, 861731, 201064, 1361495, 199247, 284743, 102845, 861750, 315107, 669986, 1165206, 352764, 201063, 379571, 669983, 865570, 1170663, 847708, 861752, 861753, 315979, 731463, 152651, 372319, 310536, 205872, 153592, 669981, 205830, 250919, 865571, 246391, 105372, 369304, 1175658, 1157245, 151615, 564036, 861738, 1165845, 205875, 315989, 317637, 1177974, 865573, 865569, 252960, 1157244, 351273, 153842, 261311, 1186170, 372333, 372320, 1175880, 565408, 314006, 565667, 220338, 430103, 1165208, 574869, 865567, 565675, 285129, 315992, 207954, 430104, 1171246, 565409, 566720, 284432, 568686, 1171233, 351275, 217364, 310534, 706896, 1171249, 566765, 330349, 847718, 151822, 315978, 201056, 574533, 568684, 1171919, 379568, 315988, 1171247, 566719, 317379, 861742, 847720, 602550, 261974, 566721, 201059, 316833, 1168163, 602543, 217370, 849589, 615212, 731460, 352300, 542067, 379805, 615213, 861754, 352302, 575996, 707182, 849593, 849583, 847719, 1171250, 1171252, 731454, 542066, 1175656, 575998, 1175664, 706933, 847717, 574870, 847709, 368169, 352301, 615214, 706932, 1171253, 847705, 369372, 847721, 542065, 861751, 668563, 1175665, 847713, 861746, 668562, 668556, 861744, 847723, 541897, 542064, 1175662, 1185626, 615210, 849587, 1185628, 1175660, 368567, 668555, 1171251, 310538, 847711, 575997, 310535, 1185627, 731456, 1175661, 849591, 1185625, 310533, 615919, 1175657, 541895, 1166406, 700836, 541898, 847715, 615918, 707181, 668554, 1175663, 1166405, 379801, 615211, 541896, 861749, 2646330, 2663215, 2660588, 2656937, 2659882, 2658710, 2646768, 2661987, 1100699, 593411, 1368001, 1546030, 2696287, 2696284, 2696288, 2696285, 2696280, 2696282, 2696281, 2696283, 2696286, 1598392, 1243826, 1189801, 1312425, 2359280, 1312429, 1368435, 1796088, 1368385, 1368419, 1368440, 1243845, 1602118, 1368403, 1368011, 2359289, 2359284, 1992837, 1368397, 1992822, 1243026, 1796094, 665043, 2359352, 1189800, 1189806, 665037, 1243833, 1164671, 1368004, 2359354, 1368401, 1992836, 2359288, 1368018, 1243842, 1368412, 1189821, 1368017, 1243018, 1796093, 1243020, 1368008, 1368392, 2359291, 1100703, 861820, 1243016, 1602120, 861770, 1243834, 1368417, 1100701, 1992827, 861771, 1100700, 1159663, 1368381, 1368400, 2359292, 2359287, 1243846, 1368410, 1243843, 1368402, 1368010, 1992830, 2359351, 1243034, 1602109, 1368426, 2359353, 1368399, 638596, 1368437, 1368416, 1368405, 1992831, 1368434, 1992835, 1243015, 2359281, 1368036, 1992829, 1189812, 1312422, 1602110, 1243038, 2359277, 1796091, 2281864, 1189810, 1368003, 1368409, 729717, 1372692, 1243850, 1167814, 1368436, 665033, 1159662, 2359356, 1368000, 1312418, 1368019, 1372754, 1161608, 1100704, 1189818, 861769, 1100705, 1243039, 1368002, 1189802, 665044, 1992824, 1368383, 757603, 1368009, 1368382, 1602113, 1243019, 2359359, 1796090, 1368020, 1189808, 1167810, 665031, 665040, 1368035, 1189811, 1368033, 1161607, 2359290, 1100706, 1368431, 1243848, 665035, 1243037, 1243839, |

| | |
|---|---|
| | 621590, 1368006, 1368423, 2359276, 1368430, 1243040, 1368433, 1602112, 1602111, 1602114, 1602108, 1189813, 1100702, 665041, 1243017, 1243033, 1368424, 1372738, 861821, 1243844, 1992832, 1189823, 1796096, 1368387, 1368394, 665034, 1312415, 665038, 2359360, 1368444, 665042, 1312411, 1992825, 1368438, 1243029, 1602115, 861819, 1167815, 1796097, 1368007, 700516, 1312416, 1243036, 1992828, 1368398, 1179164, 1602107, 1372717, 2359357, 1312423, 1243027, 1372706, 1602119, 1992826, 665039, 1189827, 1368395, 2359285, 2359286, 665036, 1796095, 1368005, 1189804, 1368396, 1992823, 1243827, 1243835, 2359282, 1189803, 1189814, 1164670, 1796089, 1602106, 1167811, 2359358, 2359283, 1368034, 1796098, 665032, 2359355, 1179163, 1368384, 1312409, 2359279, 1243022, 2359278, 1368012, 1368391, 1243849, 1796092, 1243829, 1243830, 1243847, 1243031, 748765, 704929, 757601, 700518, 1243028, 1243828, 705134, 748762, 705138, 705136, 705137, 700517, 757606, 1167812, 1189816, 748764, 1189824, 757605, 1189826, 1243023, 757602, 1243024, 1189825, 1167813, 1189822, 757608, 1189807, 1243831, 1243832, 1243030, 1189805, 1243021, 705135, 748763, 1189817, 757604, 1243035, 1189809, 1243025, 757607, 1243032, 2663153, 2656156, 2656287, 2659839, 2661905, 2655663, 2658588, 2659615, 2646947, 2648313, 2661199, 2662635, 2648864, 2654999, 2648457, 2647786, 2677163, 2677157, 2677162, 2677164, 2677160, 2677161, 2670450, 2670448, 2670442, 2677159, 2670444, 2670449, 2677156, 2677158, 2670443, 2670451, 2670446, 2670447, 2670445, 2662620, 2647239, 2663139, 2658568, 1373458, 2638675, 1992672, 1488564, 2627044, 1545653, 2638684, 2638686, 2638687, 2627054, 2627055, 2627048, 2627046, 2627047, 2659101, 2654503, 2645330, 2647565, 2646991, 2663420, 2646125, 2656735, 2663631, 2656115, 1862701, 1598392, 1545657, 1593832, 1811010, 2359280, 1811005, 1862689, 1925503, 1534343, 1592713, 1602118, 1373465, 1545162, 1925495, 1664326, 1593072, 1545147, 1992680, 2371749, 1940499, 1811011, 2359289, 2359284, 1992837, 1545164, 1593068, 1862702, 1664316, 1593831, 1992674, 1811007, 1992822, 2371728, 1486436, 1862686, 1664313, 1992694, 1593069, 1992685, 2359352, 1664323, 1593826, 1811002, 2371748, 2359354, 1992809, 1373469, 2371730, 1992836, 2359288, 1992681, 1592716, 1992701, 2371727, 1593073, 1545660, 1811001, 1862694, 2359291, 1992686, 1940497, 1545156, 1664315, 1602120, 1992691, 1664311, 1488573, 1992808, 1545667, 1545166, 1545160, 1992827, 1488569, 2169275, 1925498, 1545656, 2359292, 2359287, 1811012, 1810999, 1488567, 1486977, 1592709, 1992830, 2359351, 1373464, 1602109, 1373467, 2169274, 1811003, 2359353, 1992684, 1592717, 1545146, 1488574, 1862700, 1940498, 1992831, 1992835, 1862688, 2359281, 1992807, 1811000, 1992703, 1373466, 1992829, 1992702, 1602110, 2359277, 1488568, 1862693, 2371736, 1486981, 2281864, 1545149, 2371746, 1992810, 1992695, 2371729, 1488565, 2371734, 1373468, 2371745, 1545150, 2359356, 1373460, 1862695, 1545158, 1992700, 1992818, 1545163, 1862684, 2371731, 1992688, 1992690, 1811004, 1940495, 1992812, 1373461, 1486976, 1992689, 1940496, 1992816, 1373471, 2371737, 1593057, 1862685, 1992682, 1488566, 1992824, 1925497, 1373463, 1373472, 1811008, 1811009, 1545152, 1665368, 1593070, 1545161, 1602113, 1545154, 1664324, 2359359, 1545663, 1593776, 1545668, 1992699, 1992806, 1664322, 1992698, 1373459, 1545157, 2359290, 1545655, 1992697, 1592710, 2371724, 1664327, 1593833, 1862696, 1925496, 2169276, 1373462, 2359276, 1992819, 1545654, 1811006, 1664320, 1810996, 1602112, 1545662, 1534397, 1992820, 1602111, 1664321, 1602114, 1602108, 1593835, 1593830, 1545155, 1486975, 1862691, 1664325, 1925504, 2371747, 1593059, 1593775, 1925499, 1862698, 2371742, 1373473, 1546031, 1992832, 1665367, 1811013, 1727500, 2371725, 1862690, 1593828, 1992687, 2359360, 1373470, 1664317, 1992825, 2371726, 1925502, 1602115, 1545665, 1810997, 2371733, 1545659, 1664314, 1992811, 2371735, 1545159, 1545151, 1992828, 1664312, 1810998, 1602107, 2359357, 1534344, 1940500, 1862699, 1545145, 2371741, 1545165, 1602119, 1992826, 1592722, 2371738, 1545664, 1664310, 1593071, 1992813, 2359285, 2359286, 1545153, 1992683, 1992814, 1862692, 1664328, 1593827, 1992823, 1593829, 1862687, 2359282, 1664318, 2371723, 1545661, 1665369, 1862697, 1593058, 1602106, 1545658, 2371740, 2359358, 2359283, 2371744, 1862703, 1664319, 2371743, 1545148, 1486972, 2359355, 2371732, 1925500, 2371722, 2359279, 1545666, 2359278, 1992815, 1593774, 1925501, 1992693, 2117292, 1992821, 1486966, 2627050, 2627051, 2627045, 2627053, 2627052, 2627049, 2638692, 2638691, 2638693, 2638679, 2638685, 2638688, 2638681, 2638689, 2638690, 2638680, 2638682, 2638683, 2648119, 2660038, 2663509, 2663153, 2645846, 2659380, |

| Code System | Concept Code |
|---|---|
| | 2657345, 2658588, 2661611, 2646142, 2659871, 2646947, 2661155, 2655019, 2661199, 2662684, 2658093, 2655509, 2637865, 2637864, 2637862, 2637861, 2637858, 2637859, 2637860, 2637863 |
| **Other AOM** | |

| | |
|---|---|
| NDC | 00247044015, 11534015701, 00527174330, 67544101430, 11534016001, 10702002801, 68382013905, 17772010130, 47335070713, 70710104203, 68382035801, 61919036972, 27241023030, 70771165903, 76420027660, 76420028090, 61919036990, 76420027990, 10370036601, 27241022901, 00480235656, 80425021402, 70710104103, 726030123, 76420027860, 10370036701, 76420028030, 68382035805, 76420027630, 00480235701, 51655042926, 726030121, 76420027960, 67877086933, 76420027830, 76420027690, 804250208, 76420027890, 61919088386, 82009013705, 804250202, 10370036801, 76420028060, 82009013505, 726030120, 76420027930, 619190212, 80425037303, 80425019503, 70710103909, 70710104109, 707711656, 70771165909, 71335231802, 70710104009, 52652900103, 72603012001, 24979021104, 707711659, 80425020202, 70771165803, 804250214, 70771166009, 707101043, 80425028804, 804250195, 764200279, 80425020801, 764200278, 707711657, 70710104309, 707101041, 52652900102, 726030124, 707101042, 50090530903, 707711660, 70771165709, 70771165609, 764200280, 103700365, 713359727, 71205081930, 705183758, 67877086805, 272410230, 712050819, 830080011, 830080012, 272410228, 272410227, 103700366, 68462021860, 500906895, 103700367, 272410229, 70771165809, 17856063902, 42291051890, 29300011501, 33261048102, 12634008380, 430630573, 12634008369, 00781227610, 13668003301, 105440489, 16590022660, 16252056600, 00615813939, 16252056460, 12634045285, 16252056560, 10544062890, 43063073421, 43353069580, 12634045293, 43063060530, 33261010645, 00480015506, 13668003201, 35356047130, 12634045269, 43063099860, 00093733606, 21695034830, 33261040007, 12634045359, 006157565, 13668003274, 00245070760, 10544048930, 353560469, 000937336, 42291051918, 12634008360, 16590082360, 16590082390, 12634045363, 00781227660, 29300011601, 43353069560, 003786103, 008321072, 33261010602, 16590082530, 12634045371, 00045064575, 12634008394, 00093015506, 33261010628, 43063041730, 17772010112, 31722018305, 43602045930, 00245071460, 430630735, 00832071060, 35356047190, 17772010210, 12634045250, 16590022845, 00615563731, 33261010660, 317220181, 216950205, 12634045280, 33261048120, 33261040014, 162520569, 43063018930, 00245107315, 12634045354, 43353069660, 293000118, 16590081745, 33261048114, 42549062060, 12634045294, 16590022760, 42549062190, 00615561539, 00378610191, 31722028005, 00247211630, 00480722010, 16590082482, 216950128, 17772010107, 00245071260, 12634008393, 12634045281, 29300011705, 126340452, 12280002260, 43063072930, 13668003160, 16590022690, 12634045298, 35356047230, 00832107590, 12634045391, 126340453, 353560470, 002450707, 10544042160, 00832107430, 43063099830, 00832070960, 00245107490, 16590082430, 12634045290, 21695034860, 00378610591, 33261048028, 165710707, 12634045399, 17772010407, 42549062160, 33261010620, 16590022630, 433530697, 216950348, 006157562, 43602046030, 12634045292, 00615756539, 00143975860, 42549061972, 12634008366, 002451075, 00781227510, 21695020590, 43063018960, 00378610505, 10544084730, 10544084760, 13668003101, 00247211400, 12634045257, 10544042060, 433530696, 16252056650, 31722018105, 00045063921, 00093722006, 009046929, 29300011616, 12634008350, 002450710, 13668003331, 17772010103, 009046018, 216950130, 33261048014, 00832107115, 33261048130, 13668003360, 16590082572, 430630189, 43063099760, 43063053860, 42291051990, 12634008397, 16252056660, 12634008396, 430630436, 16571070706, 33261040000, 16590082490, 43602045810, 008321071, 425490621, 00179164970, 00093721906, 42291051818, 12634045379, 008320709, 31722028160, 29300011816, 006157563, 12634045260, 00045063942, 17772010415, 12634045261, 42291051810, 00245107230, 11722071009, 43063009460, 10544048990, 21695016290, 43602045910, 00555177909, 000937335, 00832070815, 12634045360, 33261010603, 16590054390, 33261048002, 35356040130, 12634045380, 12634045240, 000937220, 00245070960, 002451072, 00179185370, 16590082560, 13668003471, 17772010230, 177720103, 00179168871, 00832107190, 12634045259, 216950349, 31722018460, 12634045381, 43063073460, 18837015596, 165710708, 00245107330, 00480733506, 42549062090, 16590022672, 136680031, 317220183, 29300011701, 00045064065, 00480733606, 12634045367, 430630417, 16590082471, 353560471, 00045064102, 317220184, 31722028110, 17772010330, 13668003260, 000937540, 317220182, 00378610205, 12634008374, 00615756205, 008320708, 00045064101, 43353069645, 12634008371, 00480754006, 16590082472, 12634045300, 21695012860, 00247211490, 33261010690, 33261048020, 13668003171, 00179185371, 00245107115, 12634008361, 13668003205, 16590081760, 00832107515, 16571070606, |

| | |
|---|---|
| | 00904601704, 16590082330, 00832107530, 177720101, 00093015510, 12634008340, 00045063905, 216950129, 23490900003, 003786105, 31722027810, 16252056960, 29300011810, 33261040028, 12634008352, 00247211530, 16590081772, 003786102, 12634045209, 16252056860, 21695013015, 00832070760, 12280029815, 00093721910, 31722018260, 12634045242, 12634008392, 12634045398, 17772010401, 293000116, 43063073560, 008320707, 00093722010, 436020460, 26053033001, 00247211660, 162520568, 17772010403, 00045064565, 12634008357, 00904601804, 12634045291, 006158138, 16590054330, 16571070506, 12634045297, 00832107490, 12634045279, 00245107190, 00832071015, 43602045830, 16590022656, 43063073430, 126340083, 00045064765, 006157564, 13668003242, 00247211360, 16571070806, 00832107390, 001439755, 16590082486, 43063057330, 001439756, 12634008363, 00045064265, 003786101, 00832107215, 12634008384, 29300011516, 16252056400, 165710706, 00832107230, 00245107430, 12634045393, 12634045299, 001439758, 00832107130, 12634008382, 29300011710, 24236045702, 00855152930, 13668003153, 00045063965, 430630998, 33261040090, 43063061230, 00093754006, 16252056450, 12280002215, 33261048190, 13668003174, 009046016, 001439757, 12634045278, 11722077909, 00480722006, 35356047160, 43353069760, 12634045200, 43063073530, 31722018360, 00247211330, 293000115, 16590081730, 12634008395, 29300011505, 12634045295, 00045064775, 24236085902, 24236096102, 13668003460, 13668003230, 12634008367, 33261048103, 00179185372, 13668003130, 43063043630, 31722018205, 13668003305, 00247211460, 33261040021, 17772010101, 00904692961, 43063057315, 008321075, 00855153030, 00245107130, 002450708, 00480721910, 31722018160, 00555177709, 13668003401, 12634045382, 43063099730, 12634008399, 12634008345, 17772010207, 009046928, 21695016260, 12634045397, 16590022790, 00615561531, 43063073515, 16590054360, 00143975660, 17772010303, 33261048007, 00179168870, 00832070715, 105440628, 00179168872, 18837015530, 13668003271, 33261048090, 008321073, 33261040002, 00378610305, 16590082456, 31722028010, 18837015560, 00615561563, 00093733506, 42549061960, 21695034960, 00480754010, 12634008378, 31722027910, 00245107390, 17772010212, 42291051910, 21695020560, 31722027805, 00615756339, 17772010110, 433530695, 35356046930, 16252056900, 12634008398, 16590082590, 16590082445, 00179169370, 43602045710, 00480015510, 17772010315, 33261048160, 33261048000, 43063011430, 12634045309, 216950162, 42549062112, 105440847, 33261048100, 16590082462, 43353069715, 17772010312, 21695020572, 00832107315, 00378610291, 33261040060, 42549061990, 29300011605, 13668003405, 00245107290, 16590082460, 002451073, 11722077709, 008320710, 17772010410, 43353069630, 33261040003, 42549062012, 13668003477, 18837015430, 43353069780, 10544084660, 12634045263, 16590022890, 425490619, 12634045396, 43063057360, 17772010215, 16252056550, 12634045301, 43353069653, 29300011610, 00378610391, 33261010614, 29300011716, 430630997, 00143975560, 29300011805, 430630734, 00179169372, 33261010600, 12634045352, 00245071160, 00378203691, 13668003105, 12634008354, 24236045701, 12634045284, 00480721906, 43353069530, 136680032, 12634045395, 42291052090, 00832107415, 12634045390, 430630114, 12634045274, 33261048021, 436020458, 12634008342, 26053032901, 43063072960, 43063061260, 33261048060, 12634045252, 002450709, 00555071009, 00247211560, 00615756439, 12634008390, 177720102, 00615813839, 00832070915, 12634045361, 16252056760, 425490620, 43063072928, 43353069680, 12634045271, 33261010630, 21695012815, 33261040020, 12634045384, 00179164972, 00245107515, 43602046010, 42291052110, 11722078009, 177720104, 002451071, 000930155, 136680034, 12634045374, 12634045266, 12634045394, 31722028105, 17772010412, 43602045730, 12634045366, 43353069553, 16590082584, 12634008381, 00143975760, 009046017, 29300011801, 00615814039, 33261040030, 17772010201, 43353069730, 10544084560, 12634008309, 165900824, 12634008301, 16252056500, 12634008379, 00093754010, 430630605, 35356046960, 31722028060, 33261048121, 353560472, 16590081790, 21695016230, 12634008300, 16590022772, 33261048030, 43063072907, 00245071360, 42549061912, 33261010621, 16590022860, 16252056800, 12634045357, 00832107330, 436020459, 13668003415, 31722027960, 00904692861, 12634008359, 35356047060, 165900825, 35356046990, 10544041960, 317220279, 43063099721, 43063073407, 18837015460, 12634008391, 43063072921, 16252056700, 31722027860, 42549062130, 17772010203, 17772010307, 293000117, 12280002240, 00245070860, 317220278, 12280002230, 16590082582, |

| | |
|---|---|
| | 00781227560, 002451074, 35356047260, 13668003364, 16590022730, 33261048128, 00832070860, 332610400, 42291052190, 12634045254, 165900817, 00045064165, 12634045392, 12634045201, 00904601604, 00179169371, 00615756305, 33261048003, 33261010699, 00615563739, 12634045267, 332610106, 42549061930, 00615756239, 00045063901, 317220281, 21695020530, 17772010430, 12280029830, 33358034156, 29300011510, 136680033, 42291052018, 12634045378, 00378203605, 12634045245, 12634045350, 00832107290, 12634045385, 12634008385, 430630612, 43063053830, 12634045282, 165710705, 17772010301, 332610480, 12634045345, 17772010310, 12634045342, 12634045340, 23490900000, 00045063902, 21695034890, 21695034930, 42291052010, 430630094, 00378203505, 430630729, 43063060560, 17772010115, 21695012915, 33261010607, 430630538, 317220280, 00615756405, 38779244304, 00247211430, 00245107590, 008321074, 00245107530, 31722027905, 12280002290, 43063043660, 00378610105, 00245107415, 12280029860, 006158140, 13668003371, 16590022830, 42549062030, 00378203591, 00179164971, 43353069570, 12634045369, 10544048960, 12634045296, 436020457, 24236046502, 006158139, 31722018405, 000937219, 00245107215, 00245071060, 42291052118, 33261048107, 43063009430, 35356047030, 764200276, 72603012401, 72603012301, 72603012201, 80425028802, 50090392603, 80425019502, 004802359, 70710104003, 68382035830, 70771166003, 72603012101, 707101039, 70710104303, 69097012412, 70518346001, 726030122, 16571070810, 65862017305, 63629494705, 70710103903, 63629494704, 820090137, 705183460, 820090136, 71335106400, 820090135, 71335114700, 16571070610, 68382035878, 70771185803, 71205081990, 27241023001, 530021547, 80425020802, 10370036511, 55700099560, 80425028803, 71335972701, 10370036705, 80425039802, 67877086833, 82009013605, 61919067986, 51407089460, 23155089506, 59651045260, 55700099530, 51655074652, 70518375800, 71335972704, 70518150302, 51655084852, 71335231804, 71335972707, 27241022830, 71335004706, 68382035806, 71335972706, 00480235801, 71335231801, 67877086960, 53002154706, 80425037302, 27241022730, 71335972708, 51407089360, 80425020803, 83008001260, 80425009201, 71335231808, 83008001160, 59651045360, 71335972700, 61919021330, 70771185802, 10370036811, 83008001130, 50090689500, 23155089406, 80425021403, 10370036605, 10370036611, 49856276206, 62756071086, 67228040103, 658410651, 61919067972, 631870118, 65841064905, 636293994, 54569613900, 58016096124, 680711971, 516550429, 64376011910, 63187075807, 63629399401, 50090585300, 61919021260, 58118071208, 61919082590, 55887048960, 55289049730, 636296871, 68071081130, 62756071113, 58016096169, 58016047876, 58118071203, 58016035216, 61919082460, 63629818901, 627560707, 58016062600, 55887048930, 67787027910, 58016047880, 58016035236, 538081131, 58016096182, 58016035282, 60429077105, 60505276100, 60505276303, 55045312408, 58016047808, 58016047827, 50436122002, 50436995003, 62541020130, 55700021030, 61919043990, 607600094, 68071308803, 53217029190, 61919017290, 516550608, 658620171, 60429077210, 619190883, 58016062601, 58016062670, 59115012760, 55154192004, 53104012205, 58016096127, 49999069860, 548686016, 49349040202, 58016035273, 60429077010, 61919081760, 57587056960, 493490705, 680840343, 62756071083, 50090113800, 68071190009, 61919069130, 52959099402, 500903752, 58016047824, 57237013560, 61919088330, 47335071283, 58016096145, 680713196, 607600279, 54569613701, 63187022860, 66039011010, 50436995202, 58016035299, 55048079942, 551545371, 65841064805, 61786066802, 63629687102, 55048085530, 61392026225, 58016096125, 55700022760, 65841064914, 54868601700, 50090461400, 500903461, 47463080060, 65841065214, 52959064330, 61786029302, 58016096177, 66039011060, 591150125, 54569613700, 521250061, 50436013901, 58016096132, 53217029102, 50436013902, 510790727, 68071190003, 63629399505, 47335070708, 502680751, 68071048530, 680711612, 55045312406, 67787027810, 55154537200, 619190189, 50090375201, 50090113900, 63629399504, 63187005930, 61786086302, 65841065105, 47335071086, 61919021372, 67256071213, 58016096156, 658410648, 60760028060, 50090172100, 51655084826, 63629336503, 60505276005, 67544047787, 53747011601, 47463079960, 67253075410, 61919031830, 60505276103, 680840344, 53104011905, 60760028730, 55700022890, 50436013903, 52959044101, 680711960, 500903926, 680714756, 49999060530, 47335071018, 607600514, 49999060560, 68071301203, 58016035207, 61919043930, 58016035225, 58016035248, 50458064565, 63187028330, 60760064160, 58016047830, 67787028060, 66394007610, |

| | |
|---|---|
| | 67253075211, 59115012460, 617860580, 63629687101, 63629399506, 493490402, 631870479, 54868601401, 58016047893, 572370135, 58016096128, 51079072601, 68071197109, 493490907, 58016035205, 521250914, 532170291, 504580640, 58016062691, 47335070718, 54569613800, 58016047898, 493490802, 63629494701, 627560710, 55700022860, 58016035227, 60429076910, 63187047930, 49349070525, 51655042952, 58016062644, 58016062640, 58016062603, 50090346102, 47335071113, 67228040203, 59604064065, 538080921, 50090393102, 53104012204, 58016096179, 62756070713, 50090461402, 55154192402, 65841065205, 55048079930, 58016096130, 53104012002, 58016096144, 67787028010, 47463079942, 58016035256, 500902203, 49856276005, 58016047896, 63629336502, 52959099430, 636293294, 54569613901, 59604063902, 63187047960, 58016096172, 58016035235, 65841065001, 617860996, 532170274, 58016047850, 63187011830, 617860732, 58016035242, 63187096360, 64376011901, 53104012105, 59604064765, 58016096192, 61919019090, 631870963, 58016047835, 500905309, 68071081190, 58016047844, 58016062648, 50268075015, 63187075830, 631870801, 55154192008, 58016096106, 55887051760, 55700021790, 58016062692, 58016004760, 60760027990, 54569483100, 572370134, 473350707, 59115012560, 61786099602, 50268075211, 61919019060, 493490116, 52125046302, 55048085090, 60760064190, 53217029128, 60760009490, 53747011501, 53808113101, 53217027402, 68071080860, 55700022730, 49349094202, 63629329407, 60760051460, 63187022830, 52959044130, 55048088760, 58016096197, 619190369, 58016096108, 61919043972, 51079072820, 58016047869, 50090220300, 63629329402, 504580641, 49349090702, 54868601602, 458650551, 557000217, 47335071186, 516550032, 60760028030, 548684674, 633040779, 617860623, 58016062693, 51079072720, 58016035203, 58016035232, 53104011909, 50090113801, 47335071288, 55887051782, 55887012660, 58016096101, 53104012008, 54868601601, 61786073202, 58016062656, 65841065110, 58016062627, 63187077360, 58016062689, 55700089430, 50090174101, 500903389, 65841065201, 58016047875, 53217029145, 61919067930, 504369951, 617860293, 61919018530, 625410203, 66394007560, 58118071108, 63629494702, 58016096181, 49999060515, 504580647, 57237013760, 49349082002, 510790728, 63187007760, 504580639, 50090224801, 58016062628, 49856276103, 49856276106, 50436995101, 53217029130, 55700059627, 50090585301, 67787027960, 58016047809, 591150126, 60505276106, 63187080130, 58016047801, 65841064701, 636298189, 55700059690, 680711900, 49999069815, 60505276006, 58016047804, 58016062680, 55700069190, 58016047879, 63187006030, 53217031514, 64376012101, 58016096100, 55154192006, 53217029414, 47335071118, 58016062667, 54569613702, 58118070706, 47335071108, 53747011716, 50090224802, 67253075210, 52125087602, 61786029860, 458650963, 60429077160, 58016035244, 60429077260, 61919043960, 51927425400, 47335070786, 62756071183, 53104012009, 50090346100, 680713088, 62756071088, 50458064165, 61919082360, 581180711, 49856276003, 63629055990, 55700022727, 52959044190, 58016047873, 58016096190, 59604064565, 58016096103, 61919031860, 54868601402, 617860863, 473350710, 51079072620, 67787028160, 53217031502, 680713012, 557000228, 54868601603, 58016047805, 50090174100, 61919082560, 65862017260, 68071516506, 58016062650, 658410650, 532170329, 53217027460, 633040780, 47335071208, 68071301209, 66039015310, 66039010960, 49999069801, 625410204, 58016035287, 60505275105, 62756071208, 617860298, 65841064801, 50436995002, 50090393202, 658410652, 53217032960, 58016062618, 500902102, 68071196006, 61919088360, 500904614, 631870230, 53217027430, 631870773, 58016096148, 47463080030, 50090393201, 58016062609, 521250876, 58016035202, 49856276100, 59762103301, 55700059660, 53747011516, 60505276105, 58118071200, 557000894, 58016062624, 67253075310, 59604064102, 58016047883, 58016062696, 62541020330, 591150124, 55154714600, 548684672, 55700069160, 51655003252, 50090175500, 53747011710, 63304077810, 58118070708, 60505276203, 67253075311, 500903932, 631870283, 58016062607, 67544102899, 58016096167, 60429077060, 58016096105, 50090585302, 68071080830, 61919088372, 529590643, 63629399402, 61919021360, 58016035272, 47463080090, 680715125, 55700018390, 51655060826, 636294947, 658620173, 67787027805, 58016096135, 60760057790, 55048080060, 58016047800, 51079072701, 60429076960, 55700022630, 52959099460, 68071319603, 58016096115, 60760063930, 50090530901, 631870059, 504369949, 50090393101, 58016047889, 557000226, 50090392602, 47335071286, 502680753, 647250707, 53217029114, 63629399508, 49349011602, 58016004730, |

| | |
|---|---|
| | 58016096175, 60760009460, 619190823, 53747011701, 45865045960, 51655060926, 63187011860, 60760063960, 60760028760, 49856276006, 58016047890, 58016062677, 61919081730, 54868601600, 63629399407, 53217031530, 58016096184, 493490142, 58016035224, 54868467200, 548686015, 61919067960, 50458064065, 58016047867, 58016062610, 65862017359, 59604064165, 53104012209, 65862017119, 500902248, 58016047897, 59762103001, 557000210, 58016047871, 61919031872, 58016035277, 60505276300, 54868519000, 66394007660, 493490395, 64376012201, 58016047887, 68084034321, 68071512506, 58016062606, 65862017160, 58016062608, 58016096187, 68071479303, 53217031507, 54868601701, 49856276205, 58016035267, 58016062671, 54569613902, 58016096114, 58016047899, 58016047828, 55700089460, 50090210200, 58016047881, 53747011610, 63187011890, 61392026230, 47335070788, 50090338902, 68071301206, 58016035209, 53104012108, 65841065114, 60760057760, 58016096171, 680840345, 58016047877, 58016096102, 502680750, 500905853, 552890433, 62756071213, 58016062679, 51079072801, 49856276008, 53808093201, 53217029460, 53104011908, 538080967, 62541020114, 62756070718, 59604064265, 50090196200, 58016047848, 68071029530, 58016062684, 54569547300, 631870060, 50090392601, 68084034521, 58016035271, 63187096390, 60429076905, 58016096142, 62756071186, 63187077390, 63187080190, 58016035280, 50090375200, 50436122001, 55048080030, 60760064130, 49856276203, 58016096104, 604290771, 61919018960, 53104011906, 58016047807, 50436994904, 57237013660, 58016062614, 60760027890, 47335071213, 63629329403, 63187077330, 49856276308, 548685190, 50090174102, 49856276200, 58016096173, 54868601400, 58016035245, 47335071188, 58016035208, 50090375202, 572370137, 58016035270, 58016062660, 607600075, 68071161206, 493490365, 50090167600, 605052762, 49856276108, 49856276208, 49999069830, 50090392600, 50090210201, 65841065016, 58016035276, 510790726, 59762103201, 63304077930, 63187047990, 60505276205, 68071081030, 66267130206, 58016062669, 58016047832, 50090210202, 58016062635, 54868467202, 68071476003, 63629399507, 58016035291, 61919081790, 62756071018, 633040778, 58016062698, 61919018560, 58016062630, 58016047825, 63629399408, 58016096196, 57237013460, 60505276208, 63187069630, 52125085202, 63304078010, 63187069690, 68084034411, 58016035212, 58016047892, 50090167601, 617860611, 59115012660, 53104011904, 50436995103, 504369950, 627560711, 53808092101, 50090530902, 58016062621, 55154714200, 50090338900, 68071308806, 607600278, 58016035218, 53217029490, 61919082530, 47335070783, 62756071118, 631870077, 68071487703, 581180710, 58016096150, 619190679, 67253075110, 55154537100, 58016062602, 55700069130, 62756071008, 45865096360, 65862017460, 53217029107, 53217029402, 680840342, 65862017435, 60429077205, 619190190, 66267130106, 64725071001, 54868467402, 58016047818, 58016035296, 54868467400, 58016062625, 60505276206, 63629399501, 50268075111, 61919036960, 47463079930, 65862017360, 49999060501, 58016096120, 55700018330, 61919069160, 63187023090, 658620172, 59604063942, 62541020430, 53104012106, 58016047840, 58016062697, 63629399502, 50090175502, 607600287, 58016096199, 49999095560, 58118070709, 55045396401, 58016062699, 58016047815, 51655060852, 58016047820, 65841064716, 58016047816, 61919017272, 68084034211, 58016062636, 58016035201, 619190439, 504369952, 55289090130, 65841065116, 551547146, 50090224800, 54569613802, 557000227, 68071032030, 58016035284, 658620174, 680714760, 65841065216, 58016047891, 532170315, 58016096126, 58016047821, 54868601500, 60505276000, 658410647, 47335071083, 521250463, 60505276200, 548686014, 58016035293, 53808113201, 50268075115, 58016096176, 63629643101, 63187096330, 47335071088, 63187005990, 58016035226, 58016035298, 49349014228, 50090461401, 58016062682, 68084034511, 504580642, 551547142, 58016062683, 66039010860, 49349077802, 54868467401, 45865096330, 49856276000, 63629399404, 49349039502, 50090113902, 59762103101, 58016096136, 50090113802, 53217031590, 58016096198, 619190185, 49856276305, 58016047842, 52959064390, 55700022790, 59604064575, 49856276300, 636293995, 58016035269, 591150127, 53104012109, 55154192106, 59604063921, 493490118, 53217029430, 54868534301, 50090530900, 58016096193, 55045307601, 55700059630, 605052763, 617860736, 516550606, 61786061102, 53217029407, 60760028090, 53217029160, 58016062675, 61786058002, 63629336504, 548686017, 53104012006, 67787027860, 58016047803, 58016062687, 68071197103, 619190172, 50090220301, |

| | |
|---|---|
| | 68071081330, 631870696, 63304077910, 63187069660, 55700018360, 52125004702, 58016062612, 58016035206, 64725070701, 50436122004, 60687010832, 58016062632, 55887051730, 50458064265, 50090393100, 61919021390, 58016096140, 58016035204, 50268075315, 50458063965, 55700013460, 62756071218, 68084034201, 50090393200, 50090196202, 68084034401, 58016062616, 58016096118, 58016062690, 65841065101, 53747011510, 58118070703, 58016047870, 551545372, 58016096183, 604290772, 50436995001, 62756071013, 59604063965, 66039015360, 63629399400, 604290770, 58016035297, 62756071188, 58118071206, 58118071109, 61786073602, 61786029802, 66039010910, 55154192104, 49349099302, 504580645, 45865055190, 65862017261, 67228040205, 63629399503, 60760063990, 63629329405, 61786062302, 55887051790, 538080932, 58118071009, 64376012010, 63629332103, 65841064816, 49856276105, 58016096110, 58118071103, 55154192408, 49856276303, 625410201, 538081132, 58118071100, 58016062604, 619190318, 50436994902, 65841064705, 58016035292, 55700017930, 58016004700, 680715165, 493490942, 62756070788, 58118071106, 658410649, 60505276305, 54868601501, 60505276308, 63629329404, 65841065217, 500903931, 50090172102, 52959078060, 58016035281, 49349036502, 65841065014, 68071196003, 49349080202, 58016047845, 572370136, 61919082430, 49999069890, 55700021090, 636293321, 51655060626, 58016062642, 58016096170, 54868467203, 53217031528, 521250047, 63304078030, 58016035275, 60760007560, 58016062615, 65841064916, 62756070783, 50436122003, 619190817, 60505276008, 636296431, 58016047826, 58016035279, 67787028110, 58016047806, 50458064765, 65841065210, 58016047812, 60505276306, 458650459, 50268075311, 52652900101, 60429077110, 63629336501, 50268075215, 63304077830, 55700021060, 631870228, 58016062676, 54868534300, 55700022660, 50090113700, 52959064360, 61919017260, 63629494703, 52125008902, 67228040105, 68084034501, 60760027960, 58016062620, 50268075011, 63187022890, 557000691, 50090220302, 65841064814, 67228040106, 58016047872, 53217031560, 63187007730, 625410202, 647250710, 58016047810, 61919082490, 49856276306, 58016062645, 68084034301, 61919082330, 58016047802, 62756070786, 504361220, 53104012208, 58016047856, 58016062605, 58016062673, 68071081160, 53217029199, 58016047836, 45865045930, 617860668, 64376012001, 53217027490, 55154192108, 58016004790, 58016047882, 581180707, 680714877, 502680752, 63187075890, 63629399409, 58016047884, 60760027930, 529590441, 55045307606, 619190825, 619190691, 58118071003, 58016035289, 55700022818, 521250089, 61919019030, 50090338901, 604290769, 60760028790, 62756071286, 55700022830, 68071197106, 57587056860, 605052760, 504360139, 58016096112, 619190824, 55154192100, 62756070708, 55048080090, 529590994, 60429077005, 58016062626, 62756071108, 58016096109, 63629332102, 47335071008, 516550609, 52125006102, 68071029590, 473350712, 50436994903, 63187028390, 526529001, 60505276108, 55700089490, 55700021730, 58016047814, 493490820, 63629329401, 50090346101, 63187075860, 62756071283, 53104012104, 606870108, 63629336505, 60687010833, 52959044160, 68084034311, 55289043330, 58016047860, 63629399405, 63629329406, 50436995102, 59604063905, 49349011802, 581180712, 55048079960, 58016096180, 58016096107, 63874112806, 557000596, 627560712, 51655060952, 68071319606, 47335071183, 53747011616, 55700021760, 68071308809, 548685343, 50436994901, 68071475603, 493490778, 63187028360, 54868467201, 680714793, 50090196201, 47335071218, 63629332101, 47335071013, 57587056900, 58016062672, 67228040206, 65841064901, 58118071008, 57587056800, 63187080160, 52959099490, 607600577, 58016035283, 63629399403, 49349014202, 65841065005, 52125091402, 607600280, 67544047782, 55154192000, 631870758, 54868601702, 66039010810, 62756071288, 58016096160, 65841064714, 64376012110, 54569613801, 63629399406, 61919067990, 58016096191, 58118071000, 66394007510, 493490993, 60505276003, 68071190006, 58016096116, 61919031890, 58016096121, 521250852, 473350711, 53217029428, 67253075111, 53808096701, 58016096189, 62541020230, 605052761, 54868519001, 58016062681, 68180017111, 71335106405, 71335004705, 68382000416, 68258916401, 687887351, 69097012503, 68788691606, 68788896506, 68382086401, 71335048701, 68180017001, 69097081715, 687889533, 684620108, 68533056650, 68533056660, 70518289900, 69097012315, 71335033703, 68788756809, 705181853, 705182498, 71335049401, 68180017107, 68788682405, 705180370, 71335168402, 71335053805, 68788641706, 68382014001, 71335049405, 76282028010, 705180420, 70518167402, 68533056700, 68462015310, 68788953306, 727890004, 713350487, 71610019353, 70518150300, |

| | |
|---|---|
| | 70771131709, 70518200103, 71335049909, 70934074830, 70771165603, 68382013816, 68788953205, 713350047, 68533056560, 72789000460, 71335048703, 68788949905, 70518171802, 71335114707, 712050195, 713350548, 68382013801, 71335114305, 71335092505, 68788973003, 71610019380, 687889532, 68462010910, 71335054807, 705183067, 70771165703, 71610048530, 71610048660, 683820005, 707711317, 71335049400, 716100486, 70518210401, 71335032504, 68788756803, 70771131502, 70934060660, 70771131702, 27241022801, 68788746009, 70518249802, 69097081803, 681800170, 684620373, 69097081612, 71335004702, 76282028060, 69097012515, 70518119603, 68382086416, 70518295500, 68788677003, 68788896509, 707711316, 71335049402, 71335049406, 70771131605, 68462010805, 70934013330, 705182104, 681800172, 68788746006, 68462037390, 705182322, 70518119601, 70518239100, 68382014101, 71335114301, 70518165101, 71335054801, 712050233, 71335106404, 68788756805, 71335114702, 71335048709, 71335114703, 68387055890, 70934077260, 70518037000, 71335114704, 71335168403, 68258300101, 68180017003, 762820278, 70771131601, 71335032508, 68788746005, 683820139, 69097012512, 70518193700, 68180017101, 71335111601, 68387055512, 682583002, 71610048653, 68462037190, 705180333, 687886916, 70518249801, 76282028160, 71335054806, 705182899, 71335114306, 71335032503, 681800173, 68382086405, 68788953209, 71335048706, 684620374, 716100193, 71335049907, 71335053807, 690970818, 68462037130, 68533056460, 68258300201, 712050214, 71335168406, 70518306702, 70518200100, 71610019330, 69097012203, 76282027810, 71205018730, 69097012403, 68462011010, 68382076901, 71335004703, 68180017207, 716100492, 69097081815, 68382013814, 68382086406, 71335049900, 68382013901, 71205018790, 70518139300, 71335035907, 71335114307, 70518200101, 71335106406, 70518150301, 68788953303, 68788735106, 71335035906, 76282027960, 713351143, 76282028110, 71335033705, 709340733, 71610049315, 68387055960, 687887010, 71335049403, 70518193704, 705180517, 68788637703, 68258300004, 709340606, 71610048570, 713351147, 70518251700, 683870558, 71335049906, 72789000430, 68382086330, 68788896503, 68382076977, 705181651, 70771131504, 804250092, 68788637706, 69097012212, 71335048708, 70518249803, 71335168404, 68788677006, 68256300201, 71335106407, 68382000510, 76282027910, 762820279, 683820863, 71335049409, 68788735103, 71335106409, 68462011060, 71335092502, 68788701605, 709340022, 68382000514, 68788637709, 71205021460, 690970124, 687887568, 68788949903, 68115045615, 71335035902, 68382086430, 68382000410, 705182419, 68382086301, 716100493, 68180017007, 70934076830, 71335035904, 68462010960, 68382014105, 709340748, 71610048560, 68788701603, 71335168408, 68788746003, 70518241900, 705182165, 684620372, 69097012215, 68788636603, 71335035901, 71205018760, 71335092501, 68382000517, 68533056760, 71205023360, 687887460, 71205020260, 705181718, 68387055990, 69097012312, 71335048705, 707711315, 70518165103, 71335106401, 68788636609, 70518119604, 68382013914, 71205021490, 68788735109, 70771131501, 71335053802, 71335049404, 705181196, 69097081703, 68533056600, 713351064, 68788701005, 70518232201, 70518185300, 70518042000, 705182391, 68382014016, 68115091860, 71205019590, 69097081712, 70771131609, 713351684, 68462037330, 68115092760, 68533056450, 709340450, 103700368, 705180344, 71335054804, 70518033300, 68788701006, 682587156, 70518210402, 68387056060, 68462037005, 70771131509, 709340420, 70934074860, 71335114701, 68462010810, 71335049904, 71335111604, 68462037490, 71205023330, 70518216500, 71335168401, 71335035908, 71610048680, 68258904401, 684620370, 70518034400, 705181393, 68788637705, 68180017211, 70934045030, 71335114303, 71335035903, 71335114708, 70518118002, 68258919901, 683820140, 71335048704, 71205023390, 70518232202, 69097081915, 71610048580, 713350925, 71205019530, 70710104209, 68382000501, 68788896703, 68258904301, 68788677009, 68382086306, 682587056, 71335032507, 68788953309, 71335114706, 68462010860, 68382086477, 68788896505, 71335106402, 71335004701, 687888965, 68382000414, 70518118000, 68382000505, 70518239101, 70518195300, 68180017307, 71335033704, 709340133, 71335106408, 68788949906, 71335114308, 70518037001, 683870560, 71335048707, 68533056500, 713350499, 716100485, 687889730, 68180017303, 705180233, 68180017011, 71335032501, 70934060630, 68180017301, 68533056400, 705182001, 71335092504, 68382013805, 69097081903, 709340149, 683820141, 68788682409, 68788896706, 70518210400, 70771185805, 70771131703, 68788746806, 70518200104, 68180017302, 70934077230, 68788896709, 690970817, 68788953305, 684620153, |

| | |
|---|---|
| | 68788949908, 709340768, 68788746803, 687886377, 681800171, 68180017311, 68788641705, 71335168407, 68788643506, 68382013916, 68788701009, 68115045630, 76282027860, 71335168409, 68258715603, 71335111603, 68788701609, 683820864, 68788735101, 70518200105, 71335049902, 69097081603, 68462015360, 70518171801, 68382014114, 687886770, 70518062500, 690970123, 68788746805, 68382076930, 713350359, 70518295501, 68462037030, 68462015305, 68788973008, 68258705606, 713351116, 70518119600, 687886366, 70771131604, 71335114709, 762820280, 68382014014, 69097081812, 68462037290, 705180887, 71335032505, 683870559, 71335035905, 68382076916, 68788682406, 70518251701, 71335054809, 68788643505, 71610049245, 804250288, 71335033708, 71335114304, 713350337, 70518062501, 70771131705, 68788973005, 71335054808, 687886824, 713350494, 68258300002, 68387055630, 70518118001, 68462037230, 71335049407, 71335053801, 68382000401, 687887468, 70518249800, 68462037105, 70771131503, 71610049260, 68788735105, 71335033701, 70518232200, 72789000421, 71610049330, 68258715609, 68382086305, 71205020290, 690970819, 69097081912, 71335033702, 68387056090, 68382086377, 68788691603, 684620110, 682583001, 68382014005, 70518239102, 70518023300, 68382000516, 683820769, 70518306701, 70518210406, 70518088700, 70518193701, 68533056550, 68180017103, 68382014116, 68180017201, 70518200102, 68788643503, 71335114705, 70934074890, 68788636606, 705181503, 705182753, 683820004, 712050187, 71205020230, 69097012415, 68788701003, 687888967, 70934014930, 68788691609, 705181937, 682587159, 705181953, 682583000, 68382000405, 71610049360, 68382076906, 71335111605, 68258300103, 70518193702, 71335168400, 71335049901, 684620109, 68387055560, 68788643509, 68788746809, 70518193703, 70771131603, 71335004704, 68462011005, 690970816, 712050202, 68788677005, 68788691605, 762820281, 70518195301, 68258914601, 705182517, 68788756806, 71335111602, 687886417, 70934002260, 71335032506, 70771131704, 68387055930, 70934014960, 71205021430, 68788896705, 70518165100, 68256705606, 71335092503, 70934042030, 71335049408, 705181674, 70771131602, 705182955, 71205019560, 68462037305, 705181180, 687886435, 70518241901, 709340772, 70518119602, 70518167400, 71335054802, 68462037405, 68788953203, 71335049908, 70518251702, 71335054805, 68788973006, 713350538, 68462037205, 68258300009, 71335054803, 71335053806, 684620371, 70518289901, 71335033706, 70518118003, 70518210403, 71335106403, 68788701606, 68788949909, 690970125, 70518171800, 71610019360, 71335049903, 71335053804, 705180625, 68382076905, 68258300003, 68788746008, 687887016, 71335048702, 70518051700, 69097012303, 68462037090, 68462010905, 70518165102, 70771131701, 70934073360, 68462037430, 71335053803, 71610048630, 68788953206, 683820138, 71335033707, 71335049905, 68788641709, 68382086316, 68258715909, 687889499, 68788641703, 71610049253, 70518062502, 68788682403, 68258300001, 70518210405, 713350325, 68258300109, 71335114302, 71335054800, 70934002230, 70518275300, 68115091060, 71335032502, 71335168405, 70518171803, 68788636605, 70771131505, 68788973009, 70518306700, 80425009202, 690970122, 71335035909, 70518210404, 69097081615, 68180017203, 71610049230, 65862017199, 707711658, 65862017405, 10370036805, 71335048700, 71335231807, 65862017205, 65862017399, 65862017299, 71335111606, 16571070710, 70518346000, 65862017105, 71335972702, 16571070510, 00480235901, 707101040, 70518210407, 71335035900, 65862017499, 557000995, 27241022930, 70771185808, 83008001230, 516550848, 71335092506, 500906828, 80425021401, 71335972703, 27241022701, 50090338903, 70518150303, 24979021004, 713352318, 68462021960, 80425028801, 50090220303, 71335972709, 516550746, 80425039803, 249790211, 707711858, 70518193705, 71335231806, 67877086905, 83008001227, 10370036505, 004802358, 596510453, 83008001290, 00480235856, 596510452, 71335972705, 71335231803, 80425039801, 00480235601, 004802356, 50090346103, 683820358, 80425037301, 50090682700, 500906827, 71205081960, 10370036711, 67877086860, 00480235756, 004802357, 71335231805, 00480235956, 249790210, 51655074626, 10370036501, 70771185801, 50090682800, 625410204, 62541020430, 625410202, 625410201, 62541020330, 62541020130, 62541020230, 62541020114, 625410203, 71610080534, 612690565, 61269056590, 58016036100, 49999043290, 66105075009, 545694742, 53100047221, 001350461, 66336046430, 61269046090, 53100046800, 00004025652, 000040256, 66105075010, 54868415800, 66267068230, 00004025699, 54569474202, 54569474203, 000040257, 00135046107, 66105075006, 53100047100, 54569474200, 12806025609, 55045270003, |

| Code System | Concept Code |
| --- | --- |
| | 00135046108, 00247202430, 00135046102, 53100046750, 00135046105, 63629264301, 54569474207, 55887014090, 66105075003, 49999043215, 55045270008, 66336046414, 55045270009, 66105075001, 00135046101, 00135046104, 58016036130, 612690460, 54569474204, 00004025752, 00135046103, 54868415802, 53100046925, 58016036190, 55289084830, 58016036160, 54569474205, 00135046106, 63629264302, 54569474201, 54569474206, 54868415801, 52959093190, 53100046700, 51129209801, 68115044230, 68115044242, 68115083490, 68115044220, 68115083442 |
| RxNorm | 332316, 373350, 803353, 373349, 803348, 38404, 374166, 1162750, 1302823, 220343, 845478, 1436242, 1436244, 1436243, 1302841, 1494769, 1436247, 199889, 1494778, 1178002, 1302834, 1437287, 1302846, 1302847, 1302839, 1437285, 367661, 1494770, 1302860, 335759, 1436238, 1302838, 901334, 151227, 845477, 565087, 1313063, 1437278, 1302826, 199888, 1812427, 1494766, 1177986, 1436240, 564943, 1436246, 317520, 1177987, 1436245, 1302857, 1437288, 1302827, 452233, 1437283, 151228, 1494772, 1494775, 1302852, 432870, 901328, 205315, 1494780, 1302833, 564942, 1437290, 1494761, 1313060, 1437279, 1302832, 1437289, 432869, 1494777, 1302856, 1372703, 1437284, 1302824, 901335, 845476, 1302822, 1302830, 1494767, 1494782, 1812421, 901326, 1302858, 901325, 1812425, 452232, 901327, 901333, 151226, 1313059, 316839, 1812419, 1302845, 1302835, 1494774, 252413, 901324, 1494763, 1302831, 1494765, 845479, 1302850, 199890, 152855, 205316, 1437280, 1494781, 1313061, 1437281, 151229, 1494764, 1313062, 1436241, 1302849, 1494768, 1302829, 1494779, 1494762, 374165, 1437282, 1437291, 564941, 316840, 1494771, 316841, 1162749, 1178003, 1494773, 901336, 1494776, 1437286, 1436239, 1302825, 901323, 1437292, 2586426, 2586424, 2586431, 2586433, 2586430, 2586427, 573334, 366446, 220344, 213444, 2586429, 213445, 2586425, 2586428, 2586432, 573333, 1302833, 1313061, 1302839, 1302827, 1302868, 1313059, 1302850, 1302845, 1302856, 314153, 330389, 1159582, 226918, 708361, 723846, 1159583, 366436, 692875, 37925, 692876, 373151, 574045, 1186415, 1186416, 723844, 1168454, 226917, 723845, 1168455 |
| SNOMED CT | 780075002, 768105006, 387007000, 317893003, 426638000, 777002008, 116093009 |
| **Medications for alcohol use disorder** | |
| HCPCS | J2315 |

| | |
|---|---|
| NDC | 52934030000, 606870121, 51129455702, 510790241, 55045329601, 60687012195, 51079024101, 51129455701, 60687012125, 54569576700, 54868529300, 51079024106, 009047213, 00904721304, 00456333060, 004563330, 000935352, 003786333, 00378633380, 00456333001, 101350636, 00456333042, 10135063632, 00456333011, 00258400060, 00456333000, 00378633305, 422910104, 00456333063, 42291010418, 00093535286, 002584000, 68462043518, 68151476000, 70771105708, 69189043701, 684620435, 68382056910, 70771105705, 70771105709, 691890437, 681514760, 68382056928, 70771105703, 683820569, 68382056916, 68382056905, 70771105701, 68382056906, 68462043511, 70771105700, 707711057, 68382056901, 65757030001, 69364314306, 50090492600, 16729008116, 231550886, 721621566, 500906820, 70518114601, 621350242, 16729008110, 43063046915, 00406117001, 001850039, 00179179271, 00056008050, 16590089790, 00904703604, 00056001130, 430630772, 165900897, 00756030501, 00555090201, 00406008903, 16590093230, 009047036, 00555090210, 38779088705, 00406009201, 004061170, 00179197772, 00756030603, 35356055030, 35356054930, 00179179272, 00406009205, 00179197771, 167290081, 00555090202, 16590090330, 00179179270, 43063059115, 43063077270, 422910632, 00056007950, 00406011901, 16590093290, 16590089730, 430630591, 00185003930, 005550902, 42291063230, 35356055130, 38779088704, 721622154, 16590093260, 430630469, 38779088703, 00756030401, 16729008101, 00056001122, 38779088706, 00406009203, 16590089760, 00756030601, 00185003901, 00406011903, 00756030403, 00406117003, 00179197770, 00756030503, 00855106030, 16590089960, 00056001170, 35356055230, 00056001102, 00406008901, 71335001404, 70518271800, 71335148004, 691890499, 71335001406, 68788708403, 68788708409, 68788708402, 69189049901, 70518114600, 71335001402, 713350014, 71335148002, 70518131200, 68788708406, 713351480, 68788708401, 71335001405, 705182718, 68115068030, 705181312, 71335148001, 72162231101, 71335001401, 71335148005, 71335148006, 76519116005, 71335001403, 71335148003, 765191160, 687887084, 70518271801, 705181146, 47335032688, 60793043020, 68094090930, 713352062, 62135024290, 51267089007, 23155088601, 72162156603, 72162156601, 72162231103, 23155088603, 16729008117, 51552073701, 63370015810, 68084029121, 50090307600, 532170261, 500902868, 68084029111, 60793043320, 657570301, 64764089099, 657570300, 512240206, 500903929, 65757036000, 53217026130, 51285027502, 636291047, 52125072702, 52372075102, 50090294500, 52372075101, 60793053101, 61392046630, 62991124302, 680712156, 51927354800, 607930434, 63629530404, 500902866, 64764089000, 61392046660, 512670890, 50090286800, 65694010010, 61392046625, 60793043001, 500902945, 65757030302, 68094085359, 60793043520, 61392046695, 60793043501, 63629530401, 51927360200, 607930435, 60793053301, 51224020630, 680840291, 60793053601, 50090286600, 60793053701, 636295304, 50436010501, 500903076, 65757030202, 47335032618, 63629104701, 63370015815, 607930430, 47335032683, 63629530402, 62991124304, 504360105, 51129173501, 65757030042, 680940853, 60793043720, 52152010530, 647640890, 51129368001, 65757030038, 52152010504, 49452483502, 63459030042, 51552073702, 60793053201, 60793053501, 60793043301, 63629530405, 548685574, 512850275, 63459030038, 52152010502, 607930431, 65694010003, 68094085362, 51285027501, 60793043401, 63629104601, 54868557400, 52372075103, 51927275300, 60793043120, 65757030101, 62991124301, 49452483501, 50090392900, 50090492500, 636291046, 62991124303, 500904925, 51129368002, 51552073704, 65757030003, 51267089000, 607930437, 473350326, 52152010511, 51224020650, 68071215603, 63370015825, 63629530403, 60793043420, 63370015835, 521250727, 60793043701, 61392046639, 51267089099, 51927437700, 61392046631, 60793043101, 61392046665, 47335032608, 607930433, 680712721, 71335206201, 71335206205, 71335206202, 68071272103, 71335206203, 71335206206, 62135024230, 71335206204, 68094090950, 50090682000, 72162215403, 680940909, 62135024260, 68071365403, 64764089099, 43063077270, 500902868, 51267089000, 51267089007, 50090286800, 51267089099, 500902945, 512670890, 647640890, 64764089000, 50090294500, 430630772, 00247071014, 00378414105, 00247078807, 00223071901, 00182053301, 00054035725, 00046081091, 003784141, 00781107001, 00591536804, 000935036, 00603343319, 00046080952, 00855115550, 00046081050, 427940028, 00054035625, 00536376701, 00182053305, 00182053205, 00904118060, 00814262714, 00405436301, 00781107050, 000540357, 00603343321, 00719132010, 00536376806, 00839128606, 00247078814, 00046080981, 006033433, 00904118061, 00093503501, |

| Code System | Concept Code |
|---|---|
| | 00591537604, 00779037925, 006033432, 00054035613, 00591537603, 00106117530, 00591536801, 00378414005, 00677100101, 00719132110, 00054035713, 00904118151, 00591536803, 00603343228, 00591537600, 00904118160, 00247071007, 003784140, 00364033750, 000935035, 00814262708, 00536376805, 42794002808, 00603343328, 00364033705, 00364033601, 00719132013, 00719132112, 00223071801, 00182053210, 00814262514, 00603343219, 00603343121, 00247071030, 000540356, 00591536800, 00302235148, 00378414101, 00093503601, 00603343221, 00781106001, 00349846850, 00591537601, 00182053201, 00839128706, 00247078830, 00378414001, 68884070797, 68884070697, 681512694, 68151269400, 62135043230, 65473070687, 64980017101, 49884015401, 60491004130, 62135043190, 512850524, 649800172, 54868503400, 60429019601, 49884015403, 54569179001, 604290196, 621350432, 548685034, 47781060701, 63629685401, 50111033202, 52446017821, 49884015305, 512850523, 60491004134, 54868503401, 64980017203, 49884015301, 54569179000, 50111033103, 49884015405, 477810607, 62135043290, 46193068901, 55045242300, 50111033201, 51285052402, 621350431, 62135043130, 65473070601, 54569179002, 47781060730, 64980017103, 51285052302, 64980017201, 46193069006, 50111033101, 60429019630, 50973080901, 46193069005, 65473070701, 649800171, 51129162001, 54868503402, 636296854, 52446017721, 42794002908 |
| RxNorm | 2662209, 2656257, 2646837, 2660012, 802194, 803315, 82819, 1163659, 152761, 1163660, 602959, 967896, 835725, 967897, 1172304, 835727, 835728, 1172303, 835726, 639524, 602960, 477280, 602962, 349335, 803316, 602961, 802195, 539836, 484659, 539837, 602957, 639525, 602958, 637212, 863853, 1483743, 1806716, 863849, 2657989, 1806736, 859960, 863547, 451735, 2647490, 1806718, 198011, 1806707, 2659171, 1806738, 1806715, 1806724, 859970, 406374, 542208, 359808, 859962, 1806720, 2657036, 2660598, 1806701, 863851, 863850, 863844, 863848, 859967, 328402, 859966, 1806729, 1551463, 542209, 1483766, 863540, 1806722, 1483776, 637215, 105070, 863542, 1806721, 2645695, 1483782, 542210, 1483781, 1806713, 207028, 1483767, 859972, 637214, 1740873, 542211, 1483775, 1483779, 2662632, 1165401, 1806714, 1806728, 636638, 859965, 105069, 2658172, 1806726, 7243, 859961, 1806731, 1806734, 859964, 1806709, 567824, 1483745, 1165402, 1806703, 1806723, 368791, 863845, 1740871, 542214, 1806704, 1483744, 863847, 633137, 2662176, 1806698, 1806727, 859956, 859957, 372990, 384662, 863852, 863552, 1156982, 1806719, 1806737, 1806717, 866331, 637216, 637213, 1806733, 1806725, 542213, 863554, 863857, 2654875, 1483774, 1806732, 1806711, 1806699, 1806697, 2659716, 2661368, 863549, 1806730, 1740874, 863854, 1483746, 636637, 1806708, 636639, 542212, 1156983, 859971, 863846, 1483780, 1806710, 1740870, 863855, 451736, 1806700, 859969, 863856, 859958, 1806735, 2647542, 1806712, 1156981, 1483773, 406373, 1806705, 542207, 1551468, 1551466, 1187405, 1806706, 1551470, 1175293, 1551469, 1551472, 1551473, 1551464, 1173093, 1551465, 1173094, 859959, 1178315, 1178314, 1175292, 1551467, 1551475, 1551471, 1806702, 1551476, 636556, 1551474, 152092, 2660303, 2649250, 2654526, 2660229, 1551468, 2660303, 1551466, 1551470, 2649250, 1551464, 1551465, 1551467, 1551475, 2654526, 1551471, 1551476, 2660229, 1551474, 1551472, 1551473, 199731, 439133, 197624, 315842, 197623, 315841, 210628, 151358, 367893, 371940, 1159603, 439132, 1159602, 1176831, 210716, 571129, 428970, 332482, 428969, 249768, 334604, 1176832, 571203, 3554 |

| **Type 2 diabetes** |
|---|

| Code System | Concept Code |
|---|---|
| ICD10CM | E11, O24.1, E11.3521, E11.35, O24.13, E11.3411, O24.112, E11.610, E11.44, E11.3492, E11.37X2, E11.61, E11.3291, E11.3511, E11.3413, E11.37, E11.3591, E11.3211, E11.3522, E11.3599, E11.8, E11.39, E11.354, E11.3213, E11.9, O24.111, E11.3493, E11.49, E11.01, E11.638, E11.3533, E11.62, E11.341, E11.37X9, E11.3219, E11.2, E11.40, E11.34, E11.3531, E11.321, E11.3499, E11.3212, E11.3311, E11.3391, E11.3549, E11.331, E11.3542, E11.31, E11.3523, E11.349, E11.3553, E11.5, E11.3593, E11.42, E11.3512, E11.641, E11.41, O24.119, E11.3491, E11.3529, E11.640, E11.64, E11.649, E11.22, E11.359, E11.32, E11.351, E11.33, E11.59, E11.3539, E11.3312, E11.3, E11.3393, E11.311, E11.352, E11.628, E11.3292, E11.36, E11.620, E11.618, E11.630, E11.353, O24.12, E11.3513, E11.43, E11.11, E11.621, E11.0, E11.3313, E11.339, E11.622, E11.3392, E11.21, E11.51, E11.3412, E11.3293, E11.3543, E11.29, E11.3592, E11.3551, E11.00, E11.3399, E11.37X1, E11.3559, E11.69, E11.3541, E11.319, E11.3519, O24.11, E11.65, E11.329, E11.1, E11.4, E11.37X3, E11.3419, E11.10, E11.3532, E11.3299, E11.3319, E11.3552, E11.52, O24.113, E11.63, E11.355, E11.6 |
| ICD9CM | 250.30, 250.32, 250.52, 250.62, 250.12, 250.72, 250.50, 250.40, 250.00, 250.10, 250.70, 250.60, 250.80, 250.90, 250.20, 250.02, 250.92, 250.42, 250.22, 250.82 |
| SNOMED CT | 368101000119109, 420279001, 421779007, 422166005, 428007007, 609567009, 104961000119108, 82551000119103, 190389009, 422099009, 719216001, 314902007, 71441000119104, 313436004, 422034002, 199230006, 712883005, 138941000119105, 97331000119101, 82541000119100, 314903002, 359642000, 190331003, 138921000119104, 1501000119109, 713703005, 237599002, 368581000119106, 87921000119104, 9859006, 81531005, 713706002, 138911000119106, 60951000119105, 28331000119107, 41911000119107, 427027005, 1511000119107, 44054006, 420436000, 421847006, 1551000119108, 1481000119100, 421326000, 110181000119105, 237604008, 237627000, 1196922005, 420715001, 771000119108, 140121000119100, 368591000119109, 751000119104, 140101000119109, 71421000119105, 420756003, 97341000119105, 90791000119104, 711000119100, 741000119101, 90781000119102, 157141000119108, 731000119105, 721000119107, 140131000119102, 781000119106, 769220000, 703138006, 421986006, 140111000119107, 1491000119102, 127991000119101, 16748141000119100, 16745131000119100, 16746341000119103, 16745531000119108, 16745931000119102, 16745651000119101, 16747141000119104, 16746131000119105, 16746901000119101, 16747661000119109, 16747901000119104, 1003605004, 16745051000119107, 16749661000119102, 16747021000119109, 16746261000119108, 1217044000 |
| **AUD (broad/base definition)** | |
| ICD10CM | F10.1, F10, F10.2, F10.180, F10.92, F10.24, F10.920, F10.239, F10.14, F10.96, F10.10, F10.95, F10.21, F10.29, F10.150, F10.129, F10.20, F10.230, F10.282, F10.281, F10.121, F10.988, F10.229, F10.12, F10.19, F10.28, F10.959, F10.982, F10.221, F10.232, F10.25, F10.27, F10.980, F10.280, F10.11, F10.159, F10.259, F10.23, F10.951, F10.99, F10.22, F10.981, F10.26, F10.15, F10.18, F10.181, F10.288, F10.251, F10.9, F10.97, F10.182, F10.220, F10.231, F10.98, F10.921, F10.151, F10.120, F10.250, F10.188, F10.94, F10.929, F10.950, F10.93, F10.131, F10.931, F10.932, F10.139, F10.13, F10.130, F10.939, F10.132, F10.930, F10.90, F10.91 |
| ICD9CM | 305.0, 303, 305.00, 303.0, 303.00, 303.91, 303.90, 303.01, 303.02, 305.02, 303.92, 303.93, 305.01, 305.03, 303.03, 303.9 |
| SNOMED CT | 66590003, 15167005, 7200002, 284591009, 191883007, 191813001, 191882002, 87810006, 268645007, 713583005, 288031000119105, 714829008, 713862009, 10755041000119100, 97571000119109, 10741871000119101, 191811004, 231467000, 191884001, 191812006 |
| **AUD (narrow definition)** | |

| Code System | Concept Code |
|---|---|
| ICD10CM | F10.1, F10.2, F10.180, F10.24, F10.239, F10.14, F10.10, F10.21, F10.29, F10.150, F10.129, F10.20, F10.230, F10.282, F10.281, F10.121, F10.229, F10.12, F10.19, F10.28, F10.221, F10.232, F10.25, F10.27, F10.280, F10.11, F10.159, F10.259, F10.23, F10.22, F10.26, F10.15, F10.18, F10.181, F10.288, F10.251, F10.182, F10.220, F10.231, F10.151, F10.120, F10.250, F10.188, F10.131, F10.139, F10.13, F10.130, F10.132 |
| ICD9CM | 303.0, 303.00, 303.91, 303.90, 303.01, 303.02, 305.02, 303.92, 303.93, 305.01, 305.03, 303.03, 303.9, 305.00, 305.0, 303 |
| SNOMED CT | 284591009, 191883007, 713583005, 288031000119105, 714829008, 15167005, 66590003, 7200002, 191813001, 191882002, 87810006, 268645007, 191884001, 191812006, 713862009, 10755041000119100, 97571000119109, 10741871000119101, 191811004, 231467000 |
| **Alcohol-related hospitalization diagnosis** | |
| ICD10CM | F10.90, F10.91, K29.2, F10.239, F10.1, F10.230, F10.232, T51, F10.2, F10.9, Z71.4, F10.231, F10.180, F10.93, F10.92, T51.3X1S, T51.94, T51.3X1A, T51.9, F10.131, F10.24, F10.920, T51.0, T51.1X2, T51.1X1D, F10.14, T51.3X3A, T51.0X2, F10.96, F10.10, T51.2X2D, F10.95, T51.8X2, F10.931, T51.2X3, T51.2X1, T51.2X4S, F10.21, F10.932, T51.2X3D, T51.3X, T51.2X4D, F10.29, T51.8X4, T51.1X, T51.8X3D, T51.0X2A, F10.150, T51.3, F10.129, K29.20, T51.2X1D, F10.20, T51.1X4D, F10.139, F10.282, K29.21, T51.1X2S, F10.281, T51.1X3S, Z71.41, F10.121, T51.8X3S, F10.988, F10.229, T51.1X2D, F10.12, T51.3X1D, T51.0X4, F10.19, T51.0X3, T51.1X2A, T51.92XD, T51.3X2A, T51.3X2S, F10.28, F10.959, F10.982, T51.2X1A, T51.1X1S, F10.221, T51.2X3S, T51.3X3D, F10.25, F10.27, T51.8X1D, T51.3X4S, T51.1, F10.13, T51.8, T51.2X2, T51.91XS, T51.91XA, T51.1X1A, T51.1X4A, F10.980, T51.2X4A, T51.0X2D, F10.280, F10.11, T51.0X4S, T51.8X2A, T51.2, F10.159, T51.93XA, T51.1X3A, T51.8X2D, T51.94XD, T51.8X2S, T51.94XS, F10.130, T51.8X3A, F10.939, F10.259, F10.23, T51.1X4S, T51.3X3, F10.132, T51.93XD, T51.91XD, F10.951, T51.92, T51.2X1S, T51.2X, T51.8X1, F10.99, T51.3X4A, F10.22, T51.8X, T51.8X4S, T51.93, T51.0X4A, T51.3X4, T51.2X4, T51.0X, T51.2X3A, T51.2X2S, F10.981, T51.0X1A, T51.1X1, T51.0X1S, F10.26, T51.0X3A, T51.94XA, T51.8X1S, T51.1X3, F10.15, T51.3X4D, T51.0X1, F10.930, F10.18, T51.8X4D, T51.3X2, F10.181, T51.8X4A, Z71.42, T51.3X3S, F10.288, T51.91, T51.0X3S, F10.251, T51.1X3D, F10.97, T51.8X1A, T51.92XA, F10.182, F10.220, T51.0X2S, F10.98, T51.3X1, T51.0X1D, F10.921, T51.1X4, T51.3X2D, T51.8X3, T51.93XS, F10.151, T51.2X2A, T51.0X4D, F10.120, F10.250, T51.0X3D, F10.188, F10.94, F10.929, T51.92XS, F10.950 |
| ICD9CM | 303.0, 305.0, 980, 291.9, 535.3, 291.5, 291.8, 291.3, 303, 303.00, 303.91, 980.3, 303.93, 303.90, 303.01, 291.89, 303.02, 305.02, 303.92, 535.31, 980.8, 291.81, 305.03, 980.2, 305.01, 980.9, 980.0, 535.30, 303.03, 303.9, 291.82, 980.1, 305.00 |
| SNOMED CT | 66590003, 191476005, 723926008, 191480000, 288041000119101, 8635005, 308742005, 85561006, 15167005, 7200002, 713583005, 191884001, 713862009, 10755041000119100, 97571000119109, 10741871000119101, 191812006, 191811004, 231467000, 2043009, 67426006, 4953006, 6749002, 291274005, 296270002, 291211008, 291224008, 291272009, 7916009, 291269002, 216636002, 291276007, 291221000, 296340005, 296949004, 82782008, 287166006, 291220004, 291226005, 291216003, 296259009, 291270001, 296337005, 296258001, 191804003, 291273004, 291922003, 296272005, 291214000, 89507002, 212809004, 296260004, 291271002, 296950004, 296342002, 59686008, 291225009, 57346004, 291213006, 291268005, 291223002, 269765000, 216651006, 216640006, 296338000, 296263002, 296264008, 291219005, 216633005, 43695003, 191802004, 191806001, 291215004, 296951000, 291275006, 296262007, 291277003, 216648004, 291218002, 699208000, 296341009, 87460008, 426692001, 296271003, 25702006, 291222007, 291923008, 212807002, 20881008, 296336001, 18653004, 291212001, 212813006, 216635003, 191805002, 216645001, 291217007, 291924002, 52703008, 1149333003, 47237001, 461171000124100, 1145073006, 191813001, 284591009, 191883007, 191882002, 87810006, 268645007, 288031000119105, 714829008 |

| Code System | Concept Code |
|---|---|
| **Alcohol-related tests** | |
| LOINC | 55349-5, 74683-4, 93706-0, 74758-4, 5643-2, 14719-9, 58423-5, 58377-3, 104105-2, 5640-8, 45324-1, 58375-7, 58376-5, 60676-4, 58378-1, 74859-0, 58425-0 |
| **Cirrhosis** | |
| ICD10CM | K74.60, K74.3, K71.7, K70.30, K74.6, K70.3, K74.4, K74.5, K74.69, K74, K70.31, K74.02, K74.01, K74.00, K74.0, K74.1, K74.2 |
| ICD9CM | 571.6, 571.2, 571.5 |
| SNOMED CT | 266470007, 15999000, 123717006, 123716002, 266471006, 425413006, 197303009, 86454000, 197305002, 103611000119102, 725939009, 78208005, 74669004, 699189004, 27156006, 536002, 197299004, 271440004, 21861000, 197301006, 197291001, 371139006, 725940006, 197310003, 725938001, 76301009, 16070004, 266468003, 735733008, 89580002, 235896001, 197294009, 235897005, 33144001, 235895002, 45256007, 6183001, 716203000, 12368000, 43904005, 419728003, 123605001, 123604002, 197293003, 109819003, 266469006, 19943007, 420054005, 715401008, 197296006, 831000119103, 1761006, 123606000, 37688005, 31712002, 871619002, 725416005, 715864007, 1010616001, 197279005, 235901004, 235902006, 235899008 |
| **Asthma** | |
| ICD10CM | J45.21, J45.998, J45.50, J45.991, J45.3, J45.30, J45.4, J45.990, J45.20, J45.909, J45.5, J45.51, J45.42, J45.9, J45, J45.41, J45.22, J45.901, J45.40, J45.99, J45.32, J45.2, J45.90, J45.902, J45.52, J45.31 |
| SNOMED CT | 10674711000119105, 707981009, 125001000119103, 707446004, 10692721000119102, 10675431000119106, 708094006, 10675911000119109, 426656000, 10675991000119100, 10675551000119104, 707512002, 707513007, 707447008, 708095007, 427295004, 707980005, 370221004, 708090002, 707979007, 10676511000119109, 370219009, 10676431000119103, 99031000119107, 1751000119100, 124991000119109, 10675471000119109, 708096008, 10676391000119108, 442025000, 708093000, 782513000, 733858005, 135171000119106, 10676271000119104, 10675751000119107, 734904007, 786836003, 829976001, 10675391000119101 |
| **COPD** | |
| ICD10CM | J44.0, J44.9, J43, J41, J42, J44.1, J43.0, J43.8, J41.1, J43.2, J43.1, J41.0, J41.8, J43.9 |
| SNOMED CT | 285381006, 106001000119101, 70756004, 836477007, 33325001, 195959009, 708030004, 195958001, 47895001, 195957006, 233674008, 57686001, 1177120001, 135836000, 313296004, 195951007, 313299006, 13645005, 313297008, 86680006, 196001008, 87433001, 68328006, 233675009, 10692761000119107, 16846004, 77690003, 1751000119100, 16003001, 840351007, 840350008, 1010333003, 233677001, 196026004, 47938003, 23958009, 1010334009, 60805002, 266355005, 762618008, 4981000, 266356006, 31898008, 66987001 |
| **CKD** | |
| ICD10CM | J44.0, J44.9, J43, J41, J42, J44.1, J43.0, J43.8, J41.1, J43.2, J43.1, J41.0, J41.8, J43.9 |
| SNOMED CT | 23958009, 1010334009, 60805002, 87433001, 33325001, 195959009, 708030004, 195958001, 47895001, 195957006, 233674008, 57686001, 1177120001, 135836000, 313296004, 195951007, 313299006, 13645005, 313297008, 285381006, 86680006, 196001008, 68328006, 233675009, 10692761000119107, 16846004, 77690003, 1751000119100, 16003001, 840351007, 106001000119101, 70756004, 836477007, 840350008, 1010333003, 233677001, 196026004, 47938003, 266355005, 762618008, 4981000, 266356006, 31898008, 66987001 |
| **CLD** | |

| Code System | Concept Code |
|---|---|
| ICD10CM | K76.5, K76.3, K73.1, K74.02, K74.0, K76.6, K74.01, K71.3, K70.41, K73.9, K71.51, K73.8, K72.11, K73.2, K73.0, K74.60, K70.9, K70.2, K71.10, K74.2, K72.10, K76.2, K76.7, K71.50, K74.3, K71.7, K70.30, K70.0, K71.11, K71.4, K71.0, K74.00, K74.4, K70.40, K76.1, K74.5, K74.69, K70.31, K74.1 |
| ICD9CM | 571.9, 572.2, 571.6, 573.8, 573.0, 573.2, 571.40, 571.49, 571.3, 572.1, 571.0, 571.2, 571.5, 573.5, 573.3, 572.4, 571.41, 571.1, 573.9, 572.3, 571.8, 571.42, 573.1, 572.0, 572.8, 573.4 |
| SNOMED CT | 15999000, 768006009, 88518009, 767810006, 123717006, 123716002, 66870002, 197441003, 424340000, 425413006, 128302006, 235898000, 235917005, 60037002, 86454000, 235901004, 235869004, 328383001, 30188007, 768289009, 768126006, 78208005, 768125005, 281388009, 57339008, 536002, 27156006, 271440004, 21861000, 737202006, 1116000, 371139006, 186639003, 89789003, 50167007, 41889008, 767809001, 768288001, 186698009, 76301009, 266468003, 735733008, 197284004, 79607001, 62484002, 61977001, 450880008, 235896001, 235897005, 33144001, 235900003, 235895002, 76783007, 10295004, 768127002, 12368000, 43904005, 123604002, 197442005, 79720007, 235889003, 197293003, 109819003, 19943007, 420054005, 197296006, 38662009, 235899008, 1761006, 123606000, 235880004, 37688005, 31712002, 123607009, 1220580006, 1208726006, 197279005, 266470007, 266471006, 197303009, 1234822009, 197286002, 197305002, 235886005, 74669004, 197299004, 197301006, 73475009, 197291001, 197310003, 235902006, 16070004, 89580002, 197294009, 31155007, 45256007, 34736002, 123605001, 9843006, 197300007, 266469006, 190823004, 43634002, 58282009, 72925005, 74162007, 1155913007, 1010616001, 1155841005, 6183001, 314963000, 703866000, 724278007, 153091000119109, 838380002, 713370005, 723829000, 16098491000119109, 713965007, 103611000119102, 774204006, 725939009, 713966008, 702969000, 699189004, 240792005, 721710005, 10690671000119109, 725940006, 725938001, 435101000124104, 717187000, 722870008, 307757001, 871619002, 771149000, 735451005, 724766009, 863957008, 725416005, 716203000, 419728003, 838377003, 1092801000119102, 432908002, 773737004, 713181003, 715864007, 715401008, 870517000, 831000119103, 838305005, 708198006, 721847002, 1197704005, 1197739005, 1230342001, 16859701000119109, 1186865008, 1197736003, 1197150002, 1186854003, 1237346001 |
| **CVD** | |

| Code System | Concept Code |
| --- | --- |
| ICD10CM | I70.21, I70.26, I70.20, I70.92, I70.4, I70.32, I70.24, I70.31, I70.7, I73.89, I70.22, E13.5, I70.5, I70.34, I70.25, I70.33, I70.6, I70.30, E11.5, I73.9, I70.35, E09.5, I70.23, I70.90, I70.91, E10.5, I70.29, E08.5, I70.299, I70.712, I70.43, I70.723, I70.211, I70.639, I70.761, I70.611, I70.328, I70.649, E09.52, I70.748, I70.292, I70.545, I70.268, I70.608, I70.45, I70.74, E10.52, E13.51, I70.791, I70.798, I70.729, I70.202, I70.432, I70.598, I70.641, I70.709, I70.692, I70.469, I70.541, I70.629, I70.463, I70.728, I70.409, I70.56, I70.434, I70.461, I70.421, I70.468, I70.528, I70.561, I70.663, I70.442, I70.40, I70.303, I70.261, I70.603, I70.742, I70.792, I70.619, I70.44, I70.64, I70.349, I70.519, I70.72, I70.73, I70.321, I70.213, I70.293, I70.701, I70.334, I70.231, I70.249, I70.235, I70.331, I70.531, I70.422, I70.244, I70.76, I70.623, I70.418, I70.429, I70.738, I70.745, E10.51, I70.234, I70.229, I70.435, E08.59, I70.503, I70.523, I70.711, I70.221, I70.69, I70.308, I70.609, I70.662, I70.203, I70.732, I70.411, I70.511, I70.548, I70.403, I70.333, I70.438, I70.431, I70.638, I70.212, I70.309, I70.718, I70.592, I70.413, I70.799, I70.208, I70.70, I70.513, I70.462, I70.269, I70.534, I70.634, I70.699, I70.63, I70.79, I70.342, I70.643, I70.222, I70.532, I70.341, I70.601, I70.768, I70.543, I70.733, I70.749, I70.344, I70.512, I70.492, I70.223, I70.53, I70.668, I70.621, E08.52, I70.529, I70.75, I70.313, I70.518, I70.635, I70.322, I70.46, E09.51, I70.622, I70.698, I70.433, I70.542, I70.591, I70.302, I70.412, I70.661, I70.735, I70.338, I70.739, I70.702, I70.793, I70.499, I70.642, I70.535, I70.419, I70.708, I70.335, I70.62, I70.402, I70.448, E09.59, I70.41, I70.54, I70.445, I70.312, I70.323, I70.301, I70.291, I70.762, E11.59, I70.423, I70.55, I70.59, I70.311, I70.648, I70.444, I70.345, I70.743, I70.298, I70.245, I70.719, I70.219, I70.563, I70.51, I70.65, I70.509, I70.562, I70.703, I70.60, I70.443, I70.501, I70.408, I70.232, I70.722, I70.243, I70.491, I70.631, I70.568, I70.502, I70.533, E11.51, I70.744, I70.522, I70.42, I70.449, I70.329, I70.713, I70.332, I70.521, I70.218, I70.248, I70.209, I70.318, I70.741, I70.428, I70.61, I70.633, I70.441, I70.498, I70.612, I70.763, I70.401, I70.734, I70.493, I70.599, I70.731, I70.618, I70.50, I70.343, E08.51, I70.262, I70.602, I70.263, I70.628, I70.71, I70.508, I70.52, I70.201, I70.49, I70.721, I70.348, E13.52, I70.238, I70.569, I70.613, I70.693, I70.644, E13.59, I70.538, I70.319, I70.544, I70.549, I70.645, I70.539, I70.769, I70.233, I70.593, I70.239, I70.669, I70.66, I70.632, I70.241, I70.242, I70.691, I70.439, I70.339, E11.52, E10.59, I70.228, I69.390, I63.523, I63.332, I63.341, I69.322, I63.89, I63.43, I63.311, I69.361, I63.02, I63.13, I63.349, I63.321, I63.233, I63.342, I63.81, I63.219, I69.359, I63.019, I63.531, I63.429, I69.318, I63.542, I63.10, I63.20, I63.549, I69.393, I63.12, I69.30, I63.513, I63.532, I63.539, I63.312, I63.211, I63.441, I63.0, I69.352, I69.339, I69.333, I63.031, I63.232, I63.42, I63.01, I63.32, I63.323, I63.00, I63.431, I69.363, I63.40, I63.412, I63.543, I69.341, I63.112, I63.139, I69.319, I63.119, I63.329, I63.343, I63.11, I69.365, I63.212, I63.449, I63.111, I63.213, I63.422, I63.133, I69.323, I69.351, I63.113, I63.519, I63.30, I63.011, I63.039, I69.349, I69.398, I69.331, I63.419, I63.313, I63.239, I69.342, I69.311, I63.012, I63.442, I69.332, I63.443, I63.521, I63.39, I63.09, I69.328, I69.364, I69.344, I63.33, I63.132, I63.411, I63.34, I69.353, I69.320, I69.313, I63.231, I63.423, I63.19, I63.29, I69.334, I63.522, I63.03, I63.5, I63.439, I63.032, I63.331, I63.433, I63.511, I63.131, I69.310, I69.314, I63.8, I63.322, I63.541, I63.033, I69.321, I69.343, I63.59, I63.529, I63.339, I69.362, I63.49, I69.35, I63.512, I63.319, I63.44, I69.369, I63.50, I63.413, I69.312, I63.22, I63.9, I63.333, I63.421, I69.392, I63.533, I63.432, I63.013, I69.391, I69.354, I63.53, I63.51, I63.52, I63.54, I21.9, I22.2, I21.02, I21.3, I21.A1, I22.9, I22.1, I21.11, I21.A9, I21.21, I21.29, I21.4, I21.01, I21.19, I21.09, I22.8, I22.0 |
| ICD9CM | 433.21, 434.11, 433.01, 433.11, 434.01, 433.81, 433.91, 433.31, 410.40, 410.00, 410.42, 410.41, 410.31, 410.61, 410.52, 410.51, 410.50, 410.82, 410.70, 410.01, 410.20, 410.02, 410.92, 410.30, 410.12, 410.80, 410.32, 410.91, 410.11, 410.81, 410.62, 410.90, 410.21, 410.72, 410.71, 410.10, 410.22, 410.60 |

| | |
|---|---|
| SNOMED CT | 195232006, 230717002, 10349009, 230287006, 230700009, 90099008, 230518009, 64009001, 14070001, 195233001, 56267009, 195234007, 230699008, 230716006, 195186005, 57658009, 195236009, 230701008, 237701005, 195206000, 70936005, 25772007, 195200006, 195185009, 230602005, 230695002, 195190007, 230702001, 230285003, 230523009, 230698000, 230694003, 95830009, 230769007, 14309005, 230692004, 230693009, 195230003, 195189003, 88032003, 195213000, 230703006, 111298007, 206576006, 230708002, 230707007, 1153543002, 38595071000119104, 517253051000119105, 732330391000119107, 849488701000119104, 1153545009, 329541000119100, 851365731000119106, 939885431000119109, 1153544008, 168747591000119109, 496369931000119104, 759950981000119101, 251770561000119107, 329501000119102, 152148641000119104, 107557061000119108, 384430101000119103, 511452481000119102, 915141931000119109, 859422751000119101, 1153546005, 58173271000119101, 806161651000119106, 957319791000119104, 195772021000119105, 297778421000119102, 563789641000119101, 293671000119109, 1204202004, 1269243009, 1260271009, 1269237007, 1163482004, 488408691000119104, 828066161000119106, 578968971000119100, 1231168008, 1155699009, 723082006, 442024001, 705130002, 425882004, 433931000124109, 434151000124101, 195243003, 428668000, 705128004, 34191000119104, 20059004, 433901000124101, 434991000124105, 88174006, 34181000119102, 293811000119100, 307767006, 149821000119103, 307766002, 37906005, 86003009, 99451000119105, 287731003, 276219001, 140281000119108, 432504007, 371040005, 441960006, 433891000124100, 442733008, 151161000119102, 1155688007, 125081000119106, 434831000124101, 434961000124102, 23671000119107, 441991000, 293831000119105, 433911000124103, 371041009, 291121000119103, 430781000124102, 430841000124101, 16002031000119102, 672531000119106, 329431000119105, 290911000119103, 674131000119105, 16002271000119109, 16002471000119108, 430831000124106, 33331000119103, 690271000119104, 427065003, 672471000119105, 672451000119101, 329461000119102, 690251000119108, 441746000, 330411000119109, 672441000119103, 329561000119101, 9901000119100, 442676003, 12237911000119109, 302902003, 330791000119108, 672571000119109, 329481000119106, 12367511000119101, 427296003, 674101000119103, 430851000124104, 734382000, 442181008, 12237951000119105, 329361000119107, 276706004, 16703821000119101, 290901000119101, 307363008, 290931000119108, 16002111000119106, 432201000124103, 442668000, 290891000119100, 291081000119100, 408665008, 329571000119107, 672491000119106, 329391000119100, 690281000119101, 430959006, 91601000119109, 106241000119108, 690241000119106, 384993003, 674351000119101, 762630002, 672551000119100, 329401000119103, 427432001, 672511000119101, 690231000119102, 106021000119105, 290921000119105, 16002231000119106, 674401000119108, 329371000119101, 771476007, 16218291000119100, 455851000124106, 674121000119107, 329651000119102, 672381000119100, 16002431000119105, 734374000, 15707841000119101, 672461000119104, 16002511000119104, 230706003, 302904002, 672421000119109, 15707881000119106, 433951000124102, 291101000119107, 16002191000119102, 413102000, 703207000, 762651004, 46421000119102, 434881000124100, 291091000119102, 329621000119105, 444657001, 434951000124104, 762652006, 413758000, 329491000119109, 329631000119108, 724994008, 690261000119105, 16002071000119104, 15707921000119104, 102831000119104, 329421000119107, 329451000119104, 16002391000119100, 674391000119106, 329641000119104, 724993002, 690331000119109, 433941000124104, 441894009, 674161000119102, 430947007, 426107000, 703208005, 441526008, 292861000119106, 672371000119103, 733199002, 292851000119109, 455931000124109, 103761000119107, 433961000124100, 762629007, 788881005, 16002351000119105, 425642008, 291111000119105, 703205008, 330421000119102, 361000119103, 425420004, 455841000124109, 145741000119101, 54329005, 10273003, 30277009, 394710008, 42531007, 58612006, 70422006, 70211005, 194856005, 52035003, 314207007, 73795002, 233843008, 233835003, 307140009, 22298006, 194809007, 1755008, 282006, 233841005, 233838001, 233825009, 233828006, 233826005, 233831007, 233836002, 233833005, 233839009, 233842003, 194858006, 233832000, 194857001, 233840006, 233829003, 233834004, 194802003, 233827001, 233830008, 233837006, 15962541000119106, 836295007, 868214006, |

| Code System | Concept Code |
| --- | --- |
| | 836293000, 15712841000119100, 12238151000119107, 836294006, 846683001, 380001000004106, 15713121000119105, 868217004, 15712961000119108, 840609007, 15713161000119100, 868224003, 15713081000119108, 868220007, 840316004, 846668006, 868225002, 12238111000119106, 15713201000119105, 15712921000119103, 285981000119103, 32574007, 15963181000119104, 840309000, 16837681000119104, 868226001, 840680009, 17531000119105, 15712881000119105, 840312002, 23311000119105, 15713041000119103, 1204155000, 44831000087103, 15713001000119100, 1204151009, 44851000087107, 44821000087100, 44811000087108, 1204154001, 1204152002, 1204222000, 44841000087109, 428196007, 15990001, 79009004, 401314000, 62695002, 57054005, 401303003, 428752002, 129574000, 65547006, 304914007, 418044006, 64627002, 70998009, 59063002, 76593002, 703165004, 703251009, 311792005, 703212004, 703253007, 703252002, 703164000, 703209002, 703211006, 311793000, 703360004, 703213009, 311796008, 703210007, 896691006, 879955009, 896689003, 896697005, 896696001, 726499301000119105, 1208872002, 1163440003, 1208873007, 35411000087108, 35421000087100, 35431000087103, 35401000087106, 35451000087107, 35441000087109 |
| **Obstructive sleep apnea** | |
| ICD10CM | G47.3, G47.30, G47.33, G47.39, G47.34, G47.35, G47.36, G47.37, G47.32, G47.31 |
| SNOMED CT | 41975002, 1101000119103, 1091000119108, 230493001, 78275009, 230494007, 111489007, 73430006, 79280005, 105191000119100, 89911000119102, 443760008, 361208003, 774068004, 719976001, 724506009, 724504007, 765751002, 399040002, 789055001, 430390000, 719972004, 441910000, 27405005, 724508005, 724509002, 9741000119101, 426542005, 442164004, 85721000119105, 429456008, 288581000119102, 104831000119109, 16275741000119100, 24825006, 91441000119109, 724507000 |
| **Hypertension** | |
| ICD10CM | I16, I13, I12, I15, I11, I10, I15.2, I13.1, I16.1, I12.9, I11.0, I16.0, I13.2, I13.10, I13.11, I12.0, I16.9, I15.9, I15.0, I15.8, I15.1, I11.9, I13.0 |
| ICD9CM | 402.9, 402.90, 402.91, 403.11, 403.00, 404.13, 404.92, 403.01, 404.03, 403.91, 401.0, 402.1, 403.1, 404.9, 402.01, 404.10, 401.1, 404.90, 404.0, 405.09, 402.00, 404.1, 403.90, 405.99, 401.9, 404.11, 402.10, 405.11, 405.1, 405.01, 404.93, 405.91, 402.11, 404.12, 404.91, 405.0, 403.10, 405.19, 404.02, 404.00, 402.0, 403.9, 403.0, 405.9, 404.01, 405, 403, 402, 401, 404 |

| Code System | Concept Code |
|---|---|
| SNOMED CT | 1201005, 199008003, 443482000, 371125006, 56218007, 461301000124109, 1078301000112109, 429457004, 46481004, 38341003, 704667004, 65518004, 28119000, 71421000119105, 194791005, 194783001, 762463000, 26078007, 39727004, 284991000119104, 284981000119102, 73410007, 428575007, 169465000, 59621000, 78975002, 194788005, 89242004, 123799005, 57684003, 123800009, 71701000119105, 74451002, 14973001, 449759005, 10725009, 48552006, 194785008, 111438007, 31992008, 48146000, 427889009, 59720008, 397748008, 39018007, 1003729000, 1003728008, 1236831007, 1237173008, 9901000, 198983002, 198951008, 79355007, 198968007, 34694006, 8762007, 95343001, 198949009, 193003, 78808002, 198999008, 198944004, 63287004, 23130000, 46113002, 37618003, 41114007, 23786008, 24042004, 49102001, 8218002, 57873008, 238795008, 237282002, 54225002, 198952001, 49171007, 36315003, 73030000, 198997005, 18416000, 83105008, 49220004, 29259002, 198967002, 194780003, 52698002, 6962006, 237281009, 67359005, 66610008, 35303009, 15394000, 50490005, 64715009, 198946002, 199000005, 36221001, 77970009, 198941007, 81363003, 65443008, 81626002, 34273003, 71874008, 20753005, 5148006, 198942000, 72022006, 199007008, 59733002, 194781004, 31407004, 66052004, 199003007, 237279007, 65402008, 198953006, 194779001, 198985009, 198947006, 206596003, 66709007, 86041002, 199006004, 22966008, 198984008, 199002002, 69909000, 198965005, 78544004, 198945003, 199005000, 23717007, 198986005, 84094009, 38481006, 62240004, 19769006, 86234004, 46764007, 31881008, 10562009, 194767001, 1474004, 60899001, 198966006, 70272006, 128001000119105, 285911000119109, 8501000119104, 16229371000119106, 140121000119100, 697929007, 129181000119109, 422001004, 96721000119103, 140101000119109, 429198000, 434431000124103, 96751000119106, 10757401000119104, 288250001, 285041000119107, 345771000119101, 698640000, 765182005, 428163005, 285001000119105, 706882009, 367390009, 40511000119107, 285871000119106, 285831000119108, 286371000119107, 111411000119103, 712832005, 153891000119101, 720568003, 398254007, 285851000119102, 153851000119106, 275516004, 285011000119108, 712487000, 15781000119107, 473392002, 5501000119106, 285861000119100, 284961000119106, 340441000119107, 96741000119109, 766937004, 120261000119101, 96701000119107, 285881000119109, 132721000119104, 285061000119106, 367821000119106, 698591006, 129171000119106, 736992003, 140131000119102, 96731000119100, 129151000119102, 285921000119102, 104931000119100, 10757441000119102, 307632004, 108701000119102, 10757481000119107, 95605009, 40521000119100, 285841000119104, 284971000119100, 871642009, 449631000124102, 541000119105, 334831000119106, 434711000124103, 472749004, 698638005, 117681000119102, 10752641000119102, 697930002, 421731000, 285101000119109, 48194001, 140111000119107, 285081000119102, 129161000119100, 82771000119102, 127991000119101, 10759031000119106, 703310005, 96711000119105, 118781000119108, 1208845005, 1208839002, 1260271009, 1204139007 |
| **Hyperlipidemia** | |
| ICD10CM | E78.49, E78.00, E78.4, E78.5, E78.2, E78.41, E78.1, E78.0, E78.3, E78.01 |
| SNOMED CT | 15771000119109, 398036000, 402474007, 426161002, 397915002, 129590000, 137931000119102, 238084008, 402473001, 34349009, 238085009, 403827000, 402785008, 267435002, 267433009, 238088006, 402725005, 129591001, 34528009, 238040008, 773649005, 302870006, 402727002, 403829002, 403831006, 1571000119104, 402787000, 137941000119106, 773726000, 238080004, 403828005, 238089003, 402786009, 767133009, 267432004, 33513003, 238079002, 238078005, 238082007, 402726006, 238081000, 403830007, 13644009, 238076009, 238083002, 445261005, 55822004, 402475008, 238077000, 190774002, 238087001, 267434003, 701000119103, 114831000119107, 299465007, 129589009, 275598004, 1208738002, 1197489003 |
| **Major depressive disorder** | |
| ICD10CM | F32.2, F33.1, F33.2, F33.0, F32.4, F33.41, F32.0, F33.9, F33.3, F32.9, F32.1, F32.3 |

| Code System | Concept Code |
|---|---|
| SNOMED CT | 2618002, 832007, 42810003, 38694004, 70747007, 46244001, 33736005, 19527009, 40379007, 73867007, 25922000, 66344007, 77911002, 30605009, 69392006, 75084000, 71336009, 76441001, 14183003, 63778009, 63412003, 36923009, 15639000, 87512008, 68019004, 33135002, 79298009, 39809009, 20250007, 15193003, 18818009, 42925002, 36474008, 60099002, 28475009, 33078009, 191604000, 191611001, 191613003, 191610000, 268621008, 10811121000119102, 719592004, 16265301000119106, 720452006, 726772006, 16264821000119108, 16266991000119108, 10811161000119107, 16264901000119109, 16265951000119109, 16266831000119100, 720453001, 720454007, 16264621000119109, 251000119105, 104851000119103, 16265061000119105, 720451004, 281000119103, 720455008, 320751009, 430852001, 450714000, 370143000, 319768000 |
| **Opioid use disorder** | |
| ICD10CM | F11.151, F11.188, F11.19, F11.15, F11.22, F11.1, F11.2, F11.28, F11.221, F11.222, F11.259, F11.182, F11.122, F11.10, F11.229, F11.23, F11.220, F11.121, F11.11, F11.12, F11.24, F11.14, F11.29, F11.129, F11.250, F11.120, F11.282, F11.25, F11.251, F11.288, F11.150, F11.21, F11.281, F11.13, F11.20, F11.159, F11.181, F11.18 |
| ICD9CM | 304.7, 305.5, 305.51, 304.71, 305.50, 304.73, 304.72, 304.70, 305.53, 305.52 |
| SNOMED CT | 5602001, 75544000, 191819002, 1255019005, 191914006, 1255015004, 1255018002, 703845008, 191912005, 145121000119106, 191867007, 231477003, 191868002, 231479000, 1255013006, 191869005, 724653003, 191820008, 191913000, 426001001, 520841000124109, 231478008, 191865004, 231480002, 191821007, 1081000119105, 191909007 |
| **Other substance use disorder** | |
| ICD10CM | F12.188, F13.120, F13.288, F15.250, F15.159, F16.22, F14.159, F13.24, F13.229, F15.280, F15.11, F14.150, F18.180, F15.251, F18.151, F12.150, F15.282, F18.11, F18.288, F16.288, F18.20, F14.12, F13.10, F15.151, F15.181, F16.25, F14.22, F14.280, F13.25, F16.19, F14.229, F14.151, F13.150, F12.28, F14.24, F18.19, F15.122, F18.24, F13.22, F13.280, F13.19, F14.21, F16.15, F12.22, F18.15, F15.29, F18.259, F14.14, F14.121, F15.28, F14.120, F15.22, F15.12, F13.182, F13.12, F18.229, F16.159, F14.281, F16.259, F14.23, F18.27, F12.159, F15.121, F18.29, F13.230, F12.250, F14.251, F12.121, F12.25, F12.122, F18.250, F14.282, F14.182, F12.220, F15.129, F14.11, F15.229, F14.15, F15.150, F13.151, F14.29, F18.220, F15.21, F18.221, F13.259, F12.222, F13.28, F15.120, F14.220, F12.23, F14.188, F16.28, F15.15, F12.229, F15.25, F13.21, F13.220, F12.221, F16.18, F18.25, F18.159, F16.180, F12.21, F12.19, F15.180, F18.188, F15.10, F16.21, F18.14, F13.281, F18.129, F16.24, F18.280, F18.21, F16.121, F14.180, F12.11, F16.14, F15.18, F16.120, F14.25, F14.288, F18.121, F13.232, F12.120, F16.251, F16.221, F13.250, F16.188, F15.220, F12.180, F13.29, F12.259, F15.20, F14.222, F16.151, F14.18, F14.10, F13.11, F15.23, F16.280, F16.150, F13.20, F12.18, F13.121, F13.239, F12.280, F16.20, F13.18, F12.10, F15.188, F12.151, F15.182, F12.251, F16.122, F14.19, F13.282, F14.129, F12.12, F13.26, F15.24, F14.250, F18.17, F15.259, F18.120, F14.20, F18.22, F18.18, F12.129, F13.23, F16.229, F16.29, F15.222, F16.250, F14.181, F13.181, F13.27, F18.10, F16.220, F12.15, F13.129, F16.11, F12.29, F13.14, F15.14, F15.288, F14.259, F16.283, F18.150, F15.221, F14.122, F14.221, F18.12, F13.180, F15.281, F16.10, F12.20, F13.251, F13.15, F13.221, F16.129, F14.28, F13.159, F18.28, F13.231, F15.19, F12.288, F13.188, F16.12, F16.183, F18.251, F13.132, F15.13, F13.131, F13.139, F13.13, F12.13, F13.130, F14.13, F14.2, F12.1, F18.1, F15.1, F13.1, F15.2, F12.2, F14.1, F16.1, F18.2, F13.2, F16.2 |
| ICD9CM | 304.52, 305.23, 305.20, 305.21, 305.30, 305.63, 304.41, 305.43, 304.51, 305.70, 305.71, 305.41, 304.12, 305.22, 305.73, 304.13, 304.31, 305.62, 305.31, 305.72, 305.60, 304.33, 304.21, 304.43, 305.40, 304.50, 304.42, 304.10, 304.22, 305.3, 304.2, 305.7, 305.2, 304.4, 304.1, 305.6, 305.4, 304.3, 304.5, 304.20, 304.32, 305.32, 304.11, 305.61, 304.40, 304.53, 304.30, 305.33, 305.42, 304.23 |

| Code System | Concept Code |
|---|---|
| SNOMED CT | 191838006, 231472009, 78267003, 191832007, 191916008, 191849000, 85005007, 191851001, 231474005, 191918009, 231462006, 37344009, 191900006, 191831000, 191839003, 231473004, 268646008, 191901005, 191899001, 31956009, 231469002, 191893000, 84758004, 191919001, 191837001, 191850000, 21647008, 191833002, 191891003, 58727001, 191920007, 191894006, 231468005, 191895007, 1231331002, 1230088003, 1231333004, 1230072003, 1230082002, 1230087008, 1231336007, 1230266009, 1231319004, 1230089006, 1230071005, 1230086004, 1230085000, 1230083007, 1230084001, 441527004, 7071007, 74851005, 427327003, 64386003, 442406005, 86391000119101, 426095000, 70340006, 38247002, 34111000119108, 425885002, 724715004, 724704009, 762504005, 838527002, 425841004, 724695007, 724694006, 145841000119107, 275471001, 11047881000119101, 724712001, 428219007, 429692000, 145101000119102, 723933008, 724713006, 724688003, 427229002, 817962007, 144981000119109, 724657002, 762505006, 11048011000119103, 699449003, 125851000119106, 426873000, 785277001, 427205009, 724714000, 1231325000, 1231328003, 1231327008 |
